# Process Evaluation for the Delivery of a Water, Sanitation and Hygiene Mobile Health Program: Randomized Controlled Trial of the PICHA7 Mobile Health Program

**DOI:** 10.1101/2025.02.26.25322956

**Authors:** Presence Sanvura, Kelly Endres, Jean-Claude Bisimwa, Jamie Perin, Cirhuza Cikomola, Justin Bengehya, Ghislain Maheshe, Raissa Boroto, Alain Mwishingo, Lucien Bisimwa, Camille Williams, Christine Marie George

## Abstract

In the Democratic Republic of the Congo (DRC) there are over 85 million diarrhea episodes annually. Effective and scalable water, sanitation, and hygiene (WASH) interventions are needed to reduce diarrheal diseases in the DRC. Mobile health (mHealth) reminders have been shown to reduce disease morbidity and increase health-protective behaviors. Therefore, WASH mHealth programs present a promising approach to improve WASH behaviors. The Preventative-Intervention-for-Cholera-for-7-days (PICHA7) program is a targeted WASH intervention combining of mHealth and in-person visits delivered to diarrhea patient households in DRC to reduce diarrheal diseases. During the randomized controlled trial (RCT) of PICHA7, 1196 participants received weekly PICHA7 mHealth program voice, interactive voice response (IVR) quiz, and text messages over 12 months. Outcome indicators included % of unique text, voice, and IVR messages received (fidelity) and % of unique messages fully listened to (dose) assessed using the engageSPARK mobile message platform, and program reach to households assessed through monthly follow-up visits. 84% of households received unique text messages and 90% of unique voice and IVR messages were answered. Households reported receiving a PICHA7 mHealth message in the past two weeks at 72% of surveillance visits (844/1177). 74% (309/418) of participants reported sharing a PICHA7 mHealth message with another person at least once. These findings show high fidelity, dose, and reach of mobile message delivery in the PICHA7 mHealth program. This study demonstrates the feasibility of delivering the PICHA7 mHealth program in eastern DRC and provides important insights for delivering WASH mHealth programing in low– and middle-income countries globally.

**Conflict of interest:** The authors declare no conflicts of interest.

## 1. Introduction

There are an estimated 2.9 million cases of cholera annually resulting in over 95,000 deaths worldwide (1, 2). The Democratic Republic of the Congo (DRC) has one of highest rates of cholera in Africa (3). Previous studies have identified consuming untreated water, stored drinking water being uncovered, using an unimproved water source, lack of handwashing with soap, and consuming food outside one’s home as significant cholera risk factors (4, 5). Household members of cholera patients are at a 100 times higher risk of cholera infections compared to the general population during the 7-day period after the cholera patient is admitted to the healthcare facility (6, 7). Furthermore, this high risk of diarrheal disease transmission to the household members of diarrhea patients spans beyond cholera to other enteric diseases including patients with *Shigella* and enterotoxigenic *E. coli* (8, 9).

Water, sanitation and hygiene (WASH) interventions have the potential to reduce cholera and severe diarrhea globally. A previous study in the DRC found that delivery of a hygiene kit with soap and a handwashing station to suspected cholera patient households resulted in a 56% lower incidence of suspected cholera (defined by diarrhea, vomiting, or healthcare facility visit for diarrhea) (10). Furthermore, our previous randomized controlled trial (RCT) in Bangladesh found that delivery of the targeted Cholera-Hospital-Based-Intervention-for-7-Days (CHoBI7) WASH program, which included a health promoter visit in the healthcare facility and a similar hygiene kit, resulted in a significant reduction in bacterial culture confirmed cholera (11). However, major challenges remain in identifying scalable approaches to promote sustained WASH behavior change over time for high risk populations for cholera.

Mobile health (mHealth) presents a scalable approach to deliver health communication programs at a low cost which has been shown to increase disease prevention behaviors, with increasing evidence in low and middle income countries (12–14). In Bangladesh, we developed the CHoBI7 mHealth program, with weekly automated voice, interactive voice response (IVR), and text messages to diarrhea patient households for 12 months combined with a single in-person healthcare facility visit during the time of treatment for the diarrhea patient (15). The RCT of the CHoBI7 mHealth program found this WASH intervention significantly increased handwashing with soap, improved stored drinking water quality, and reduced diarrhea prevalence and stunting in diarrhea patient households (16). In India, a WASH mHealth program for new mothers conducted also observed increases in handwashing with soap behaviors (17). Similarly, in Kenya, a mHealth program which sent weekly messages four times a week for three months on diarrhea symptoms, causes, and prevention measures to mothers found that participants had higher rates of handwashing with soap and water treatment behaviors compared to a control group, though these measures were self-reported (18). These findings suggest that mHealth is a promising approach to increase WASH behaviors and reduce diarrheal diseases. However, to date, no RCTs of a WASH mHealth program have been conducted in sub-Saharan Africa.

To address this gap and reduce diarrheal diseases in the DRC, our research group developed the Preventative-Intervention-for-Cholera-for-7-days (PICHA7) mHealth program. This program delivers automated weekly voice, IVR, and text messages promoting handwashing with soap, water treatment, and safe water storage from a doctor at a local cholera treatment center. These messages are sent over a 12-month period using the web-based engageSPARK platform. Our recent RCT of 2334 participants of the PICHA7 program found that the program significantly increased handwashing with soap and water treatment relative to free chlorine, improved stored drinking water quality relative to *E. coli*, and reduced both diarrhea and stunting in young children (George et al. 2025, in press). The objective of this present study was to conduct a process evaluation of the PICHA7 mHealth program to assess the fidelity, dose, and reach of this program during our recently completed RCT.

## Methods

### 2.1 Study design

The RCT of the PICHA7 mHealth program enrolled 2334 participants in Bukavu, South Kivu province, DRC from December 2021 to December 2023 in a two-arm cluster RCT (where a cluster is defined as a diarrhea patient household). Bukavu is an urban city with a population of over one million in eastern DRC (19).

The target population for this study was diarrhea patients and their household members. Daily diarrhea patient surveillance was conducted at 115 public and private health facilities from December 2021 to December 2022. Eligible diarrhea patients met the following criteria: 1) diarrhea patient admitted to a health facility with three or more loose stools over a 24-hour period; 2) provided a blood and a stool sample within 24-hours of enrollment; 3) had a child under five years old living in the household; 4) planned to reside in Bukavu for at least 12 months; 5) had resided at home for the three nights before hospitalization; 6) no functioning tap to collect water inside home; and 7) had at least one working mobile phone in home. After enrollment, diarrhea patients were randomly assigned to a study arm and their household members were enrolled. Randomization (1:1) of diarrhea patients to study arms was conducted using a random number generator, with assignments carried out by the study biostatistician (JP) using R (version 3.3.0). Household members were eligible if they planned to reside in the same household as the index diarrhea patient for the next 12 months, shared meals from the same cooking pot, and had resided in the same home with the diarrhea patient for the past three days. Additional details on the study design and recruitment is reported elsewhere (George et al. 2025, in press).

The RCT compared the standard discharge recommendations for diarrhea patients in the DRC on oral rehydration solution (ORS) use for rehydration (standard message arm) to the PICHA7 mHealth program (PICHA7 arm). The PICHA7 mHealth program was developed through community-centered formative research, including semi-structured interviews and a pilot study of 518 participants (19). Delivery of the PICHA7 mHealth program to diarrhea patient households included an initial in-person healthcare facility visit by a health promoter, two home visits during the 7-day period following diarrhea patient healthcare facility admission, quarterly home visits for 12 months, and weekly voice and IVR calls for the 12-month program period. Diarrhea patient households received a summary text message following each voice and IVR message. This study focuses specifically on the mHealth component of the PICHA7 program.

mHealth messages were developed and tailored during formative research. Two characters deliver the PICHA7 mHealth messages: “Dr. Picha” and “Mwanza”. Dr Picha is a doctor at a hospital who calls and texts participants to share information and reminders on handwashing with soap and water treatment behaviors. Mwanza is a woman who brought her young child to the healthcare facility for diarrhea treatment and learned proper handwashing with soap and water treatment behaviors from Dr. Picha. Dr. Picha is played by RB, a physician on our research team who provides care to diarrhea patients. RB reviewed the content of messages and recorded the automated messages sent to participants.

Subscription to the PICHA7 mHealth program was performed by a health promoter who entered a shortcode into the phone of the diarrhea patient and their household members at the healthcare facility. To train diarrhea patients and their household members on receiving and responding to PICHA7 mHealth messages, the health promoter sent a practice voice, IVR, and text message to the mobile phones of the household members of patients present in the healthcare facility. They also provided support with technical challenges encountered when answering practice calls, responding to practice IVR messages, and opening text practice messages. A total of 63 unique voice, 65 unique IVR, and 128 unique summary text messages were sent during the 12-month program period. Example IVR, voice, and text messages are provided in Supplemental File 1. The web-based engageSPARK platform was used to send PICHA7 mHealth program messages. If the participant did not receive a call from the PICHA7 mHealth program (missed or call failed) a subsequent call was sent after 15 minutes, 30 minutes, 1 hour, and 24 hours. Details of study recruitment and enrollment procedures are published elsewhere (George et al., 2025 in press). Messages were sent to all mobile phones in the household, and phone owners were encouraged to share messages with other household members.

Study process evaluation indicators are presented using the Medical Research Council framework for process evaluation (20). Program fidelity was defined as at least 80% of program households receiving program voice, IVR, and text messages. Program dose was defined as the percentage of voice, IVR, and text messages answered (voice and IVR received) and fully listened to (voice and IVR) by program households. Program reach was defined by the percentage of program households that reported receiving a PICHA7 mHealth message in the past two weeks.

### 2.2 EngageSPARK platform process evaluations indicators

The process evaluation indicators assessed using the engageSPARK platform were the following: 1) the percentage of PICHA7 voice, IVR, and text messages received by program households; 2) the percentage of PICHA7 voice and IVR messages fully listened to by program households; and 3) the percentage of program households replying correctly to PICHA7 IVR quiz responses. The percentage of PICHA7 unique text messages received by program households was calculated by dividing the total number of unique text messages received by the total number of unique text messages sent. The percentage of unique voice and IVR messages answered by program households was calculated by dividing the total number of unique messages answered by the total unique number of messages sent to the households. The engageSPARK platform was also used to determine how long (call duration) a recorded voice and IVR message was listened to before the call end. If a voice or IVR message was answered, it was classified as “fully listened” if at least 80% of the message was completed (based on the length of the full recorded message), otherwise this was classified as “partially listened”. The percentage of PICHA7 unique voice and IVR messages fully listened to by PICHA7 mHealth program households was calculated by dividing the number of unique messages fully listened to by the total number of unique messages answered.

For IVR messages, responses options for a question asked were pressing either 1 or 2 based on which statement the participant thought was correct. When the respondent selected a response, the answer was classified as “replied” and given the status of “correct” or “incorrect” (pressed 1 or 2), “, or “invalid” (not 1 or 2 (e.g. 3)). The percentage of IVR messages replied correctly was calculated by dividing the total number of IVR messages replied “correct” by the total number of IVR messages replied to.

### 2.3 Participant reports

Participants 12 years of age or older were administered monthly in-person questionnaires to assess their interactions with the mHealth program. Program reach was assessed using the percentage of households and participants that reported receiving or sharing a PICHA7 mHealth message in the past two weeks. Information was also collected on the percentage of individuals in program households that reported challenges with receiving PICHA7 mHealth messages during the program period. The percentage of households and participants that reported interacting with a PICHA7 mHealth message in the past two weeks was calculated by dividing the number of households and participants (>12 years) that reported receiving a mHealth message in the past two weeks by the total number of household and participants (>12 years) with monthly surveillance data available at each time point.

### 2.4 Data collection for diarrhea, handwashing and water E. coli outcomes

During the study period we conducted monthly clinical surveillance of diarrhea (3 or more loose stools in a 24-hour period in the past two weeks) among all diarrhea patient household members (all age groups). Unannounced spot check visits were conducted at Day 7 and 1, 3, 6, 9, and 12 months after enrollment in a randomly selected subset of 100 households per study arm for water sample collection (storage water and water source) for the *E. coli* analysis. Five-hour structured observation was conducted on Day 7 and 1, 3, 6, 9, and 12 months after enrollment in a randomly selected subset of 50 households per study arm to observe handwashing with soap behavior at key events. Handwashing with soap was defined as washing both hands with a cleansing agent (bar soap, detergent powder, liquid soap, ash or soapy water). Key events for handwashing recorded during structured observation include 1) before food preparation (food event), 2) before eating (food event), 3) before feeding someone (food event), 4) before serving food (food event), 5) after going to the toilet (stool event), 6) after disposing of stool (stool event), and 7) after washing the anus of children (stool event).

### 2.5. Statistical analysis

Descriptive statistics were used to define the study population. We used logistic regression to assess the impact of IVR quiz responses on diarrhea, handwashing with soap, and household stored water *E. coli* concentration at the subsequent month. Predictors were: 1) correct answer for IVR message response vs. did not answer, and 2) answered IVR message vs. did not answer. Diarrhea, handwashing with soap, and household stored water *E. coli* concentration at the subsequent month (next month) were the outcomes and were collected between 20-40 days after the IVR message was sent. All regressions were performed using generalized estimating equations to account for clustering at the household and participant levels. Analyses were completed using SAS version 9.4.

### 2.6 Ethical approval

The PICHA7 research program was approved and validated by the ethics committee of the Johns Hopkins University (JHU), Bloomberg School of Public Health under the ethical number 9848 and the Catholic University of Bukavu (UCB) under the ethical number 7107. Informed consent was received from all participants or guardians.

## 2. Results

Overall, 1196 participants were enrolled in the PICHA7 mHealth program (Table 1). There were 475 participants over 12 years of age, of whom 71% (335/475) were over 18 years of age. The mean age for PICHA7 participants was 27 years and 68% (321/475) of participants were female. Five percent (22/462) of participants reported refrigerator ownership, and 99% (458/462) of participants had at least one household member who could read and write. The average number of mobile phones per household enrolled in the PICHA7 program at baseline was 1 ± 0.7 (standard deviation) (range: 1-5), and the average household size was 8.0 ± 2.7 (1–15).

**Table 1.** Baseline mHealth participant characteristics.

|  | % | n | N |
| --- | --- | --- | --- |
| Households |  |  | 180 |
| Participants |  |  | 1196 |
| Participants $\geq 12$ years of age | | | 475 |
| Participants $< 5$ years of age | | | 406 |
| Primary caregivers |  |  | 251 |
| Mean ± SD (min-max) | 1.5 ± 0.9 (1-7) | | |
| Household size |  |  | 170 |
| Mean ± SD (min-max) | 8.0 ± 2.7 (1-15) | | |
| <sup>§</sup>Age (years) |  |  |  |
| Mean ± SD (min-max) | 27.1 ± 12.7 (12-79) | | |
| 12-18 | 29% | 140 | 475 |
| $> 18$ | 71% | 335 | 475 |
| <sup>§</sup>Gender |  |  |  |
| Female | 68% | 321 | 475 |
| Phones per household |  |  |  |
| Mean ± SD (min-max) | 1.0 ± 0.7 (1-5) | | |
| <sup>§</sup>Household Roof type |  |  |  |
| Concrete | 5% | 24 | 462 |
| Other | 95% | 438 | 462 |
| <sup>§</sup>Household income |  |  |  |
| Household income | 96% | 441 | 457 |
| No household income | 4% | 16 | 457 |
| <sup>§</sup>Refrigerator ownership | 5% | 22 | 462 |
| <sup>§</sup>At least one household member who can read and write | 99% | 458 | 462 |

SD = standard deviation.
<sup>§</sup> Among participants $\geq 12$ years of age. Participants $\geq 12$ years of age assessed for interaction with mHealth program and represent primary group of interest for analysis.
All primary caregivers $\geq 12$ years of age.
Household characteristics (household roof type, household income, refrigerator ownership, reading and writing information) reported at the participant level.

Eighty-four percent of unique text messages sent to PICHA7 mHealth program households (15132/18042) during the 12-month program period were received (Table 2). Ninety percent (8122/9046) of unique voice calls were answered by program households, and 86% (7021/8122) of these voice calls were listened to fully. Ninety percent (8394/9298) of unique IVR calls were answered by program households and 78% (6542/8394) of these IVR calls were listened to fully. All households that received an IVR message replied to at least one IVR message during the study period. Overall, 60% (2372/3985) of unique replies were correct, and 22% (884/3985) of unique replies were invalid. A full summary of household responses to unique IVR quiz questions is found in Supplemental File 2.

**Table 2.** Summary of PICHA7 pilot mHealth program mobile messages sent to households (N=180)

| Message type | Indicator | All messages |  | Unique messages |  |
| --- | --- | --- | --- | --- | --- |
|  |  | n | % | n | % |
| Text | Sent | 38467 | - | 18042 | - |
|  | Not delivered | 11390 | 30% | 2910 | 16% |
|  | Delivered | 27077 | 70% | 15132 | 84% |
| Voice | Sent | 66480 | - | 9046 | - |
|  | Not delivered | 926 | 1% | 33 | <1% |
|  | No answer | 50379 | 76% | 891 | 10% |
|  | Answered | 15175 | 23% | 8122 | 90% |
|  | Partially listened | 3463 | 23% | 1101 | 14% |
|  | Fully listened<sup>†</sup> | 11712 | 77% | 7021 | 86% |
| Interactive voice response | Sent | 64048 | - | 9298 | - |
|  | Not delivered | 926 | 1% | 32 | <1% |
|  | No answer | 48074 | 75% | 872 | 9% |
|  | Answered | 15048 | 23% | 8394 | 90% |
|  | Partially listened | 4865 | 32% | 1852 | 22% |
|  | Fully listened<sup>†</sup> | 10183 | 68% | 6542 | 78% |

Unique messages indicate the number of unique messages sent to the household, and if at least one of each unique message was delivered/answered/listened to.
Fully listened indicates at least 80% of the message was completed before the call was ended.

PICHA7 household members reported receiving a program mHealth message in the past two weeks at 72% (844/1177) of surveillance visits (Figure 1). Seventy-three (309/422) percent of participants reported sharing a PICHA7 mHealth message with another person at least once during the study period (Table 3). Twenty-three percent (59/259) of participants reported that distracting or noisy background sounds made it hard to hear voice calls during the program period (Table 4), 22% (58/258) of participants reported that they were unclear on how to reply to a IVR message, 21% (56/270) reported that their phone was damaged, and 20% (53/261) reported the poor mobile phone reception made a voice/IVR call hard to hear. No significant association was found between replying to an IVR message and diarrhea prevalence, handwashing with soap, or household stored water *E. coli* concentration (Table 5).

**Figure 1.**
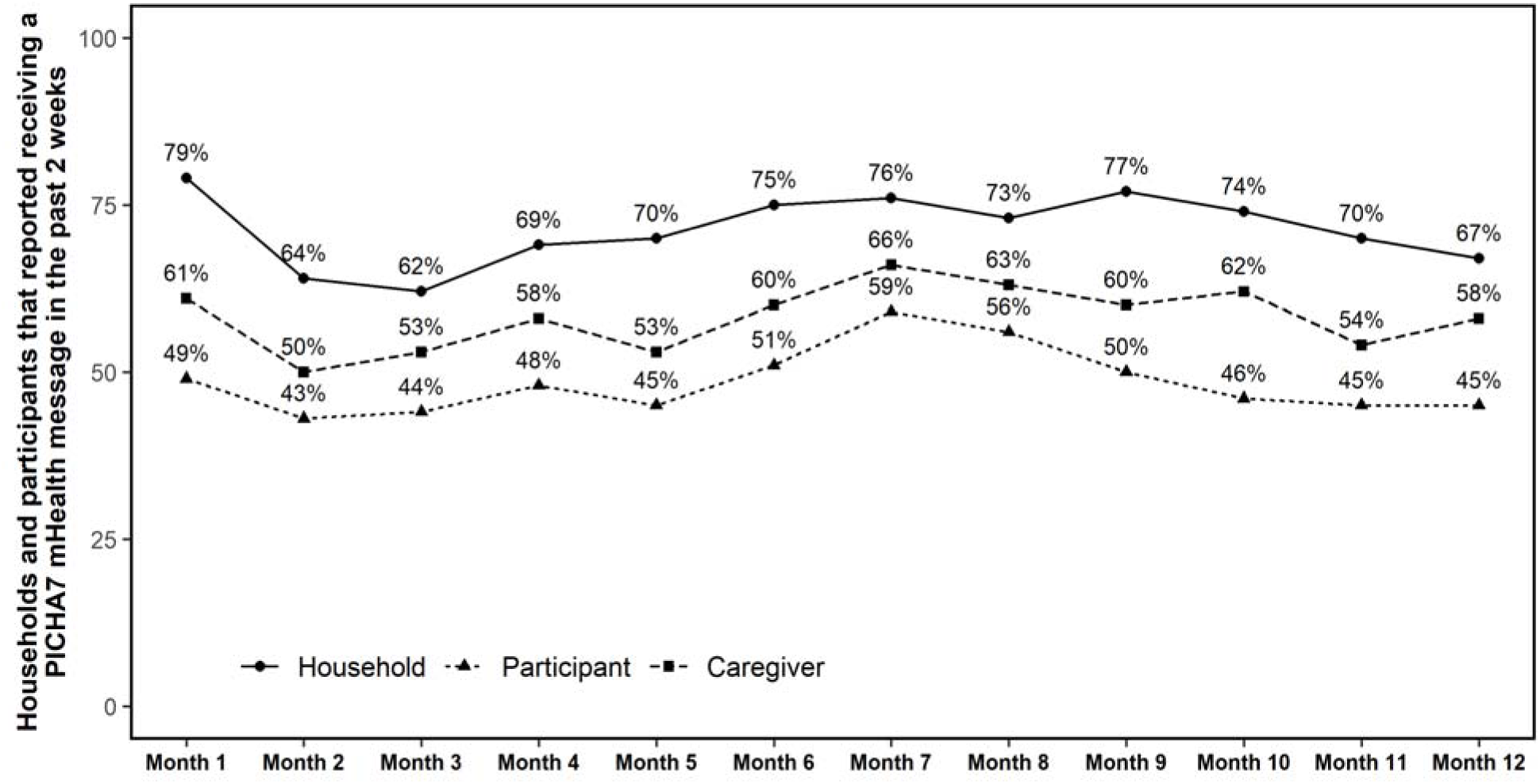
PICHA7 Participant reporting receiving an mHealth message in the previous 2 weeks. Only includes participants age 12 or over.

**Table 3.** Participants reporting sharing a PICHA7 mHealth message with others and those reporting someone shared a message with them during the study period (N= 422)

|  | You shared with<br>% (n) | Someone shared with<br>you % (n) |
| --- | --- | --- |
| Any sharing | 73% (309) | 64% (270) |
| Spouse | 27% (113) | 38% (159) |
| Neighbor | 5% (20) | 44% (187) |
| Children | 20% (84) | 62% (261) |
| Parents | 31% (131) | 32% (136) |
| Grandparents | 3% (12) | 9% (39) |
| Sibling | 16% (68) | 43% (180) |
| In-law | 3% (11) | 7% (31) |
| Friends | 6% (25) | 46% (193) |
| Co-worker | 1% (6) | 11% (48) |
| Other | 10% (41) | 5% (19) |

More than 1 response possible per participant at each timepoint.
Sharing messages in the past 2 weeks (month 1 visit and later).
N=total number of responses over study period.
Only includes participants age 12 or over.

**Table 4.** Reported challenges with the PICHA7 mHealth program over the study period (N = 416)

| Challenge | % |
| --- | --- |
| Distracting/noisy background made it hard to hear the voice call | 23% |
| Unclear on how to reply to quiz (question where we asked you to press 1 or 2) questions | 23% |
| Phone is damaged | 21% |
| Poor cell phone reception made voice call hard to hear | 20% |
| Not enough messages received—need more guidance | 17% |
| Accidentally hung up on voice call before message finished | 15% |
| Message arrived at time inconvenient for reading the text or listening to the voice call | 14% |
| When you answer the call it is cut by the sender after a few seconds | 14% |
| Phone is Lost | 14% |
| Too busy to receive messages | 13% |
| Did not have time to listen to full voice call | 12% |
| Accidentally deleted SMS before reading it | 12% |
| Too many messages received—burdensome | 11% |
| No one available to read the SMS | 10% |
| Message not shared by phone owner | 9% |
| Afraid to reply to quizzes (question where we asked you to press 1 or 2) because might be charged | 9% |
| Full inbox blocked incoming messages | 9% |
| You are concerned that if you press the wrong answer something bad will happen | 8% |
| You cannot read the SMS Message | 5% |
| Instructions in the messages are confusing | 4% |
| Message content not interesting or pertinent to needs | 3% |
| Other | 26% |

More than 1 response possible per participant.
Challenges in the past 2 weeks assessed during monthly visits.
N=total number of overall participants
Only includes participants age 12 or over.

**Table 5.** Association between a response to a interactive voice response quiz message, diarrhea in the previous 2 weeks, and WASH behaviors at household visit the subsequent month.

| Group | Correct answer for IVR message response<br>(vs. did not answer) |  |  |  |  | Answered IVR message<br>(vs. did not answer) |  |  |  |  |
| --- | --- | --- | --- | --- | --- | --- | --- | --- | --- | --- |
|  | Correct answer |  | Did not answer |  | OR (95% CI) | Answered |  | Did not answer |  | OR (95% CI) |
|  | % | N | % | N |  | % | N | % | N |  |
|  | % diarrhea in past 2 weeks |  |  |  |  | % diarrhea in past 2 weeks |  |  |  |  |
| Diarrhea |  |  |  |  |  |  |  |  |  |  |
| All ages | 7% | 3101 | 7% | 1572 | 0.92 (0.7, 1.22) | 7% | 4282 | 7% | 1572 | 0.86 (0.66, 1.12) |
| Children < 5 years | 12% | 1198 | 13% | 602 | 1.02 (0.74, 1.4) | 12% | 194 | 13% | 602 | 0.96 (0.71, 1.3) |
| Children < 2 years | 16% | 569 | 19% | 309 | 0.91 (0.64, 1.3) | 16% | 779 | 19% | 309 | 0.88 (0.63, 1.22) |
| Handwashing with soap |  |  |  |  |  |  |  |  |  |  |
| Any event | 49% | 618 | 49% | 293 | 0.85 (0.59, 1.23) | 49% | 851 | 49% | 293 | 0.9 (0.64, 1.26) |
| Food event | 45% | 552 | 47% | 256 | 0.89 (0.57, 1.38) | 44% | 731 | 47% | 256 | 0.93 (0.62, 1.39) |
| Stool event | 45% | 303 | 40% | 139 | 0.95 (0.61, 1.48) | 47% | 407 | 40% | 139 | 0.99 (0.67, 1.45) |
| Household stored water *E.coli* concentration |  |  |  |  |  |  |  |  |  |  |
| < 1/100 mL CFU | 90% | 227 | 89% | 104 | 1.22 (0.57, 2.58) | 89% | 303 | 89% | 104 | 1.04 (0.54, 2.01) |
| < 100/100 mL CFU | 98% | 227 | 96% | 104 | 1.8 (0.47, 6.94) | 89% | 303 | 96% | 104 | 1.7 (0.48, 6.02) |
| < 1000/100 mL CFU | 99% | 227 | 100% | 104 | - | 99% | 303 | 100% | 104 | - |

Handwashing for participants age 2 or greater
OR = Odds ratio
CI = Confidence interval

## 3. Discussion

The process evaluation of the PICHA7 mHealth program demonstrated high fidelity, dose, and reach of program calls and text messages to households. Ninety percent of unique voice and IVR calls were answered and over 80% of text messages were received by program households, with the majority of unique voice calls (86%) and IVR calls (78%) being fully listened to. Furthermore, program households reported receiving a PICHA7 mHealth message in the past two weeks at the majority of surveillance visits (72%). A high proportion of message sharing was also observed among those receiving program mobile messages (74%). These results demonstrate that the PICHA7 mHealth program presents a promising approach to deliver voice and IVR calls and text messages to reinforce the WASH behaviors promoted during in-person visits for program delivery. These findings complement our RCT of the PICHA7 mHealth program which showed that when combined quarterly in-person visits, this program was effective in significantly increasing handwashing with soap and water treatment behaviors and lowering diarrhea prevalence and stunting during the 12-month program (George et al. 2025 in press).

The success of the PICHA7 mHealth program in terms of fidelity, dose, and reach is likely attributed to the community-centered formative research conducted over an 18-month period prior to the implementation of the program. This formative research allowed us to tailor the PICHA7 mHealth program to address identified barriers to effective implementation. For example, during the pilot study conducted as part of this formative research, we identified that participants had difficulty understanding some voice and IVR messages. In response, we simplified the language in these messages to accommodate those with limited or no formal education before implementing the RCT program (19). During the pilot, participants also requested that voice and IVR calls be sent in the evenings between 7 and 9 PM instead of between 4 and 6 PM, to ensure most household members were home and could listen to messages together. In addition, the pilot highlighted the importance of an initial orientation at the healthcare facility, where a practice voice call, IVR call, and summary text were sent to each phone of diarrhea patient household members as part of hands on training. This training allowed for major technical challenges with message delivery to be addressed at the start of the program. Future WASH mHealth programs should similarly engage community members in intervention development and delivery and conduct pilot studies prior to large-scale program implementation.

Our findings are consistent with our previous CHoBI7 WASH mHealth program in Bangladesh, which also demonstrated high fidelity, dose, and reach in program delivery to diarrhea patient households (21). In the CHoBI7 mHealth program, 83% of voice calls and 86% of IVR calls were fully listened to and 92% of text messages were received by program households. At the 12-month timepoint, 78% of program households reported receiving a CHoBI7 mHealth message in the past two weeks during surveillance visits. This program was also developed through community-centered formative research (15). Consistent with PICHA7, the CHoBI7 mHealth program significantly increased handwashing with soap and water treatment behaviors while also reducing diarrhea prevalence and stunting. These findings suggest that WASH mHealth programs that combine mHealth with in-person visits can be effectively delivered to reduce diarrhea and improve child growth in diarrhea patient households in distinct contexts such as the DRC and Bangladesh.

Despite its success, the delivery of the PICHA7 program presented several challenges. These included background noise making calls hard to hear, participants being unclear on how to reply to IVR calls, damaged phones within households, and poor cell phone reception. Similar challenges have been reported in other mHealth studies conducted in Africa, where participants also faced difficulties related to changing phone numbers, limited time for intervention interactions, and technical issues (ex. only receiving part of messages or receiving messages in two parts) (22, 23). Low average message response rates and difficulties with IVR message replies have also been reported in other studies in sub-Saharan African (22, 24, 25). However, despite these challenges, mHealth programs in Africa have generally been found to be acceptable and capable of achieving desired outcomes. For example, one study in South Africa on a preconception health trial found higher self-reported SMS text intervention adherence and better hemoglobin levels among those that received a 2-way text intervention compared to those that did not, however there was only a 11% message response rate (22).

The PICHA7 mHealth program has some limitations. First, the PICHA7 mHealth program was conducted in an urban area in South Kivu and therefore findings cannot be generalized to rural areas. Future studies should pilot the PICHA7 program in rural areas of DRC. Second, we focused on households which reported having at least one functioning mobile phone. Future studies should include households that have shared access to phones. Additionally, voice, IVR, and text messages were only provided in Swahili, the primary language in our study setting. Future programs should include multiple language options at sign up to reach a broader audience.

This study has strengths. The first strength is the monthly surveillance of the mHealth process evaluation indicators which allowed us to investigate program reach. Second, the use of the process evaluation indicators from the engageSPARK platform (fidelity and dose) to assess call and message status (received, answered, full listened and key pressed for quiz messages). Finally, the investigation of the role of program household IVR responses on WASH and health outcomes.

## 4. Conclusion

The PICHA7 mHealth program had high fidelity, dose, and reach for voice and IVR calls and text messages sent to program households. These findings demonstrate the feasibility of delivering the PICHA7 mHealth program in our study setting in eastern DRC and provides important insights for developing WASH mHealth programing in low– and middle-income countries globally. The PICHA7 mHealth program is a promising approach to increase WASH behaviors and reduce diarrhea and stunting in DRC.

## Data Availability

All data produced in the present study are available upon reasonable request to the authors.

## Acknowledgements

We thank the study participants, and the dedicated field research officers including Pacifique Kitumaini, Alves Namunesha, Willy Felicien, Freddy Endelea, Feza Rugusha, Blessing Muderhwa Banywesize, Raissa Boroto, Gisèle Nsimire, Claude Lunyelunye, Emmanuel Buhendwa, Pascal Kitumaini Bujiriri, Jessy Tumusifu, Brigitte Munyerenkana. We would also like to thank the following individuals for their support with this study: Avner Mizrahi and Md. Sazzadul Islam Bhuyian. We thank also the engage SPARK team, including Christine Joy Hortinela and Temitope Olamolu. This work was made possible with funding from UK aid from the Foreign and Commonwealth Development Office funding grant number 215674Z19Z provided to Christine Marie George at Johns Hopkins School of Public Health. The views expressed do not necessarily reflect FCDO’s official policies or views. The funding agencies had no involvement in study design, data collection, data analysis, and data interpretation.

