## Supplemental File 1 for "Process Evaluation for the Delivery of a Water, Sanitation and Hygiene Mobile Health Program: Randomized Controlled Trial of the PICHA7 Mobile Health Program"

### Preventative-Intervention-for-Cholera-for-7-days (PICHA7) Mobile Health Message Bank

This file contents a comprehensive list of SMS, voice, and interactive voice response (IVR) messages sent as part of the PICHA7 Program randomized controlled trial completed in 2023. This program delivers automated weekly voice, IVR, and text messages promoting handwashing with soap, water treatment, and safe water storage from a doctor at a local cholera treatment center. These messages are sent over a 12-month period using the web-based engageSPARK platform. The recent RCT of 2334 participants of the PICHA7 program found that the program significantly increased handwashing with soap and water treatment relative to free chlorine, improved stored drinking water quality relative to *E. coli*, and reduced health facility visits for diarrhea and stunting in young children. Messages were adapted for PICHA7 from the larger WASHmobile program, of which PICHA7 is the Democratic Republic of the Congo-specific adaptation.

WASHmobile
a water, sanitation, and hygiene mobile health program

PICHA7
a WASHmobile program

| ID | Time | Type | Topic | WASHmobile mHealth messages adapted for PICHA7 | Summary SMS |
|---|---|---|---|---|---|
| **1** | **Day 1** | **Voice** | Introduction Message on 7-Day High Risk Period and Hardware Set-up | Hello! My name is Dr. Picha from Provincial General Referral Hospital in Bukavu. My team has visited your family recently to share with you the information about cholera. In the next twelve months, I will be calling you twice a week to remind you how to protect your family from cholera and other severe diarrheal diseases. Our calls and messages are free. Right now, and for the next 7 days, your family is at very high risk for getting severe diarrhea again. Please properly use the handwashing station and safe water storage bucket that we provided to help prevent this. Have you set up the blue safe water storage bucket on the blue stool? Please also add the chlorine tablet and definitely cover it with the lid! Also make sure the handwashing station is sitting on the red stool provided, with the soapy water bottle or bar soap and a clean cloth next to it. Be sure to share this message with your family! I'll talk to you again. Take care! | For the next 7 days, your family is at very high risk for getting severe diarrhea again. Put your handwashing station on the stool provided and add chlorine tablets to the safe water storage container. Share the message. -Dr. Picha |
| **2** | **Day 1** | **IVR** | Importance of WASH materials for reducing cholera and severe diarrhea | Hello again, this is Dr. Picha from the Provincial General Referral Hospital in Bukavu. As you know, right now and for the 7 next days, you and your family are at high risk for getting cholera or severe diarrhea. It is always important to practice the teachings that my team has taught you. I would like to ask you a question. You can reply back to me by pressing 1 or 2 on the mobile phone. After you press 1 or 2 then you will receive the correct answer. Your reply is free.<br><br>Using materials such as the soapy water bottle, hand washing station, drinking water storage bucket, chlorine tablets, and stools we provided will help to prevent cholera and severe diarrhea. Press 1 if you think there is no way to prevent cholera and severe diarrhea. Press 2 if using materials such as the soapy water bottle, hand washing station, drinking water storage bucket, chlorine tablets, and stools we provided will help to prevent cholera and severe diarrhea. You will not be charged for your reply.<br><br>**If 1 was pressed:** Thanks for trying. Using the soapy water bottle, hand washing station, drinking water storage bucket, chlorine tablets, and stools we provided can prevent cholera and severe diarrhea! I will call you again. Take care!<br><br>**If 2 was pressed:** Excellent. Using the soapy water bottle, hand washing station, drinking water storage bucket, chlorine tablets, and stools we provided can prevent cholera and severe diarrhea! Please make sure the handwashing station is sitting on the red stool provided, with the soapy water bottle or bar soap and a clean cloth next to it. I will call you again. Take care!<br><br>**If no button was pressed:** We did not hear a response from you. The correct response is using the soapy water bottle, hand washing station, drinking water storage bucket, chlorine tablets, and stools we provided can prevent cholera and severe diarrhea! Please make sure the handwashing station is sitting on the red stool provided, with the soapy water bottle or bar soap and a clean cloth next to it. I will call you again. Take care! | Using all the materials (soapy water bottle, hand washing station, drinking water storage bucket, chlorine tablets, stools to place the buckets and others) that we provide will help you and your family to prevent the contamination of cholera or severe diarrhea. -Dr. Picha |

| | | | | | |
|---|---|---|---|---|---|
| **3** | **Day 2** | **Voice** | Water treatment during the 7-day high risk period | Hello again! This is Dr. Picha from the Provincial General Referral Hospital in Bukavu. I am here again with Mwanza, the mother of a young boy that came to the hospital with cholera. We are discussing the importance of water treatment during the 7-day high-risk period. Cholera germs can be present in our drinking water. These cholera germs cannot be seen with a naked eye, and many germs do not have a smell. Even water that looks clean could have cholera germs.<br>**Dr. Picha:** Mwanza, can you tell us how to treat drinking water?<br>**Mwanza:** Yes Dr. Picha. I fill up my blue bucket you gave me with water to the top, add one chlorine tablet, and put the lid back on the bucket. I then wait 30 minutes. This about the time it takes to boil cassava.<br>**Dr. Picha:** Excellent Mwanza. I know we all have busy lives with all of our little ones running around. How do you find time to treat your drinking water?<br>**Mwanza:** I treat my water early in the morning when I wake up to make sure this water is ready for my family to drink.<br>**Dr. Picha:** Wonderful Mwanza. You are also a champion like Mwanza. Share this message! Be well and stay safe. | To protect your family from cholera and severe diarrhea during this 7 day high risk period, make sure you treat all of your drinking water with a chlorine tablet. Share this message! -Dr. Picha |
| **4** | **Day 3** | **IVR** | How to correctly treat water with chlorine tablets | Hello, Dr. Picha speaking. Have you been using the chlorine tablets? (pause) You might have noticed there is a taste and odor when you drink the water; this is normal and means the water is safe to drink.<br><br>I would like to ask you a question. You can reply back to me by pressing 1 or 2 on the mobile phone. After you press 1 or 2, you will receive the correct answer. Your reply is free. How long after adding the chlorine tablet should you wait to drink? Please press 1 on your phone if you think 15 minutes. Please press 2 on your phone if you think 30 minutes. Again, responding to this question is free.<br><br>**If 1 was pressed:** Thanks for trying. 15 minutes is not long enough for the chlorine to work. Be sure to wait 30 minutes before drinking the water. Remember that it is about the same amount of time as cooking rice. Please drink the safe chlorine-treated water and ask your family to do the same! I will call you again. Take care!<br><br>**If 2 was pressed:** 100% correct! 30 minutes is long enough for your water to be safe to drink. Remember that it is about the same amount of time as cooking rice. Please drink the safe chlorine-treated water and ask your family to do the same! I will call you again. Take care!<br><br>**If no button was pressed:** We did not hear a response from you. Be sure to wait 30 minutes before drinking the water. Remember that it is about the same amount of time as cooking rice. Please drink the safe chlorine-treated water and ask your family to do the same! I will call you again. Take care! | You may notice a taste or odor when you drink chlorinated water. This is normal and means the water is safe to drink. Wait 30 minutes after adding chlorine to drink. Share the message. -Dr. Picha |

| | | | | | |
|---|---|---|---|---|---|
| **5** | **Day 5** | **IVR** | Key times for handwashing with soap | Hello again, this is Dr. Picha from the Provincial General Referral Hospital in Bukavu. To protect yourself from cholera during the 7-day high-risk period for you and your family, you and your household members must always wash your hands with soapy water or soap during the key moments you were taught.<br><br>I would like to ask you a question. How many key times are there for washing hands with soap? You can answer by pressing button 1 or button 2 on your phone. Your answer is free. Press button 1 on your phone if you think there are 4 key moments. Press button 2 on your phone if you think there are 5 key moments. After you press 1 or 2, you will receive the correct answer. Again, your answer is free.<br><br>**If 1 was pressed:** Nice try. Actually, there are 5 key moments for handwashing with soap or soapy water. These 5 key moments are: before preparing food, before eating, before feeding the child, after using the toilet, and after wiping or cleaning a child's stool. Please wash your hands during these 5 key moments to keep your family healthy and happy. Share this message with your family. Stay healthy.<br><br>**If 2 was pressed:** Excellent. Great job. There are 5 key moments for handwashing with soap or soapy water. These 5 key moments are: before preparing food, before eating, before feeding the child, after using the toilet, and after wiping or cleaning a child's stool. Please wash your hands during these 5 key moments to keep your family healthy and happy. Share this message with your family. Stay healthy.<br><br>**If no button was pressed:** We did not hear a response from you. There are 5 key moments for handwashing with soap or soapy water. These 5 key moments are: before preparing food, before eating, before feeding the child, after using the toilet, and after wiping or cleaning a child's stool. Please wash your hands during these 5 key moments to keep your family healthy and happy. Share this message with your family. Stay healthy. | Protect against severe diarrhea by washing your hands with soap before preparing food, before eating, and before feeding a child, and after using the toilet or touching child feces. Share the message. -Dr. Picha |
| **6** | **Week 1** | **Voice** | Handwashing with soap and boiling water | Hello again! This is Dr. Picha from the Provincial General Referral Hospital in Bukavu. Congratulations! You and your family have made it through the riskiest 7 days for contracting cholera! The chlorine tablets we gave you may have run out by now, but that is okay. Now you can heat your water until it reaches a rolling boil. Continue boiling your water to make it safe for drinking, and store it in your blue bucket with the lid on.<br><br>Even though the risky 7 days are over, keeping your family safe is a lifelong task – keep up with handwashing with soapy water at all five key times and using the handwashing station and safe water buckets to help you do this. Keep the red bowl under the red bucket and use them! Share this message with your family! Until next time, stay well. | Congratulations for completing the riskiest 7 days. Keeping your family safe is a lifelong task. Continue to wash hands with soapy water at the five key times using the handwashing station. The chlorine tablets may have run out by now, but that is okay. Boil your water and store it in your blue bucket with the lid on. -Dr. Picha |

| | | | | | |
|---|---|---|---|---|---|
| **7** | **Week 1** | **IVR** | How to correctly boil water | Hello, this is Dr. Picha on the phone from the Provincial General Referral Hospital in Bukavu, congratulations for completing the 7-day high-risk period. It is important to remember that your household members are still vulnerable to cholera and diarrheal diseases. It is important that you always treat your drinking water. The chlorine tablets may have run out by now, but that is okay. Boil your drinking water.<br><br>I am here with Mwanza, the mother of a patient that came to the CTC for treatment. She has forgotten the correct way to boil her water and has asked for your help. Please help her. You can answer by pressing the 1 or 2 button on your phone. Your answer will be free. What is the correct way to boil your drinking water? Press the 1 button on your phone if you need to boil your water for 1 hour for it to be safe. Press the 2 button on your phone if you need to boil water until large bubbles rise up. You will receive the correct answer after you press 1 or 2. The answer to this is free.<br><br>**If 1 was pressed:** Nice try. Boiling water safely is easy. You only have to boil water until large bubbles rise up, a rolling boil. After boiling, please put your water in the blue drinking water vessel we provided with the lid on. Only use the tap to dispense water. Share this message. We will talk again soon.<br><br>**If 2 was pressed:** Correct. Boiling water safely is easy. You only have to boil water until large bubbles rise up, a rolling boil. After boiling, please put your water in the blue drinking water vessel we provided with the lid on. Only use the tap to dispense water. Share this message. We will talk again soon.<br><br>**If no button was pressed:** We did not hear a response from you. Boiling water safely is easy. You only have to boil water until large bubbles rise up, a rolling boil. After boiling, please put your water in the blue drinking water vessel we provided with the lid on. Only use the tap to dispense water. Share this message. We will talk again soon. | Keeping healthy is a lifelong task. Your chlorine may be gone, but thats okay. Boil your water. It’s easy. Heat water until large bubbles rise up then put it in the blue bucket with the lid on. -Dr. Picha |
| **8** | **Week 2** | **Voice** | How to prepare a soapy water bottle | Hello again! This is Dr. Picha from the Provincial General Referral Hospital in Bukavu. I have Mwanza, the mother of a young child that came to the hospital with cholera, here with me again today. She has a question about handwashing with soap and safe water for me to keep her family safe from cholera and severe diarrhea.<br>**Mwanza:** Dr. Picha, my soapy water bottle is empty. What kind of soap can I use to make more?<br>**Dr. Picha:** Detergent is great for making soapy water, just use 7 capfuls in any half-liter plastic bottle. This soapy water helps you to make your hands germs free. Make more soapy water bottles to keep in the kitchen and bathroom area. Wash your hands at every key time, everyday - not for one or two days, then you, your child and your all family members will be healthy and well! Take care. | Powder detergent is great for making soapy water. Just use 7 capfuls in any half-liter plastic bottle. It works as well as liquid or bar soap for getting rid of germs. Share this message. -Dr. Picha |

| | | | | | |
|---|---|---|---|---|---|
| **9** | **Week 2** | **IVR** | Water treatment | Hello, this is Dr. Picha from the Provincial General Referral Hospital in Bukavu. I hope you and your family are well. I am here with Mwanza again today, the mother of a patient that came to the Provincial General Referral Hospital in Bukavu for diarrhea. She wants to tell her story.<br>**Mwanza:** We collect our drinking water from REGIDESO. But my son just got sick again with severe diarrhea. What happened?<br>**Dr. Picha:** For those listening, I have a question for you. You can answer by pressing button 1 or button 2 on the phone. After pressing button 1 or button 2, you will be given the correct answer. Your answer is free. Do you need to treat drinking water collected from REGIDESO? Press the 1 button for No. Press the 2 for Yes. After you press 1 or 2, you will receive the correct answer. Again, your answer is free.<br><br>**If 1 was pressed:** Nice try. In many parts of Bukavu, the pipes are old. Many of these pipes have small holes that allow germs and worms to enter that make us sick. Even water that looks clean and clear can have these bacteria, since they are invisible to the naked eye. Mwanza, did you know REGIDESO water could be contaminated with germs and worms that can cause severe diarrhea?<br>**Mwanza:** No! I thought REGIDESO was safe, and my family would stay healthy.<br>**Dr. Picha:** For those listening, always boil or treat your water collected from REGIDESO for drinking. Stay healthy!<br><br>**If 2 was pressed:** Correct. In many parts of Bukavu, the pipes are old. Many of these pipes have small holes that allow germs and worms to enter that make us sick. Even water that looks clean and clear can have these bacteria, since they are invisible to the naked eye. Did you know Mwanza REGIDESO water could be contaminated with germs and worms that can cause severe diarrhea?<br>**Mwanza:** No! I thought REGIDESO was safe, and my family would stay healthy.<br>**Dr. Picha:** For those listening, always boil or treat your water collected from REGIDESO for drinking. Stay healthy!<br><br>**If no button was pressed:** We did not hear a response from you. In many parts of Bukavu, the pipes are old. Many of these pipes have small holes that allow germs and worms to enter that make us sick. Even water that looks clean and clear can have these bacteria, since they are invisible to the naked eye. Did you know Mwanza REGIDESO water could be contaminated with germs and worms that can cause severe diarrhea?<br>Mwanza: No! I thought REGIDESO was safe, and my family would stay healthy.<br>Dr. Picha: For those listening, always boil or treat your water collected from REGIDESO for drinking. Stay healthy! | Did you know REGIDESO water can be contaminated with germs that can cause diarrhea? Many of these pipes are old and have small holes that allow germs to enter. Boil your water to keep your family safe. -Dr. Picha |
| **10** | **Week 3** | **Voice** | Handwashing with soap | Hello again! This is Dr. Picha from the Provincial General Referral Hospital in Bukavu. We will remember that to protect ourselves from cholera, it is very important to wash your hands with soap. I'm here with Mwanza. She is having some trouble remembering when to wash her hands. Can we help her? Mwanza, how many key times are there for hand washing with soap?<br>**Mwanza:** hmm…. I think 3 times…..after using toilet…after cleaning child's anus…and when…..<br>**Dr. Picha:** Hmm, that is a good start! But there are 5 key times for handwashing with soap. The 5 key times are: After defecation, after cleaning a child's feces or anus, before preparing food, before eating, and before feeding children.<br>**Mwanza:** Oh, thank you Dr. Picha! I Actually, my phone was not active the last few days and my husband also forgot to share your phone messages with us. But I think it is important for those with phones in the household to share the messages from Dr. Picha.<br>**Dr. Picha:** Thank you! That's fantastic, everyone! Please share your voice and SMS messages with all household members, especially your wives. | Make sure your whole family washes their hands with soap at the 5 key times to stay healthy: After defecation and cleaning a child's feces, and before preparing food, eating, and feeding children. Share this message! -Dr. Picha |

| | | | | | |
|---|---|---|---|---|---|
| **11** | **Week 3** | **IVR** | Chlorine tablets are available at pharmacies in your area | Hello again, this is Dr. Picha from the Provincial General Referral Hospital in Bukavu. I am here again with Mwanza, the mother of a patient that came to the Provincial General Referral Hospital in Bukavu for treatment. She has run out of chlorine tablets. She is scared her baby will get sick again.<br><br>For those listening, we need your help. I have a question to ask you. You can press button 1 or button 2 on your phone to respond. By pressing button 1 or button 2 on your phone, you will receive the correct answer. Your message is free. Can Mwanza buy chlorine tablets for her drinking water in the market areas in Bukavu? Press 1 for No. Press 2 for Yes. By pressing button 1 or button 2 on your phone, you will receive the correct answer. Your message is free.<br><br>**If 1 was pressed:** Nice try. Chlorine tablets are available at pharmacies in Bukavu. We tested AquaTabs, Oasis, and Pure in the laboratory, and they were found to be effective in killing germs. Add one chlorine tablet to the blue 20-liter bucket we gave you, and then wait 30 minutes before drinking. If you don’t have money for chlorine tablets, you can boil your water until it has big bubbles. Store drinking water in your blue bucket with the lid on. We will talk again soon!<br><br>**If 2 was pressed:** Congratulations, that's right. Chlorine tablets are available at pharmacies in Bukavu. We tested AquaTabs, Oasis, and Pure in the laboratory, and they were found to be effective in killing germs. Add one chlorine tablet to the blue 20-liter bucket we gave you, and then wait 30 minutes before drinking. If you don’t have money for chlorine tablets, you can boil your water until it has big bubbles. Store drinking water in your blue bucket with the lid on. We will talk again soon!<br><br>**If no button was pressed:** We did not hear a response from you. Chlorine tablets are available at pharmacies in Bukavu. We tested AquaTabs, Oasis, and Pure in the laboratory, and they were found to be effective in killing germs. Add one chlorine tablet to the blue 20-liter bucket we gave you, and then wait 30 minutes before drinking. If you don’t have money for chlorine tablets, you can boil your water until it has big bubbles. Store drinking water in your blue bucket with the lid on. We will talk again soon! | Chlorine is available at pharmacies in Bukavu. We tested Aquatabs, Oasis, and Pure in our laboratory. They were all effective at killing germs. Share the message. -Dr. Picha |
| **12** | **Week 4** | **Voice** | Handwashing with soap and water treatment | Hello again! This is Dr. Picha from the Provincial General Referral Hospital in Bukavu. We continue to remind you that cholera and severe diarrheal disease can affect everyone, rich or poor, man, woman or child, who does not respect the rules of hygiene and does not treat their drinking water. Let's wash our hands every time with soapy water and boil or treat our water with chlorine to protect ourselves and our families from cholera and severe diarrhea. Please share this message with your family to keep them healthy and happy. Be well and stay safe. | Severe diarrhea can affect everyone, rich or poor, man, woman or child who does not respect hygiene rules by washing hands with soap or ash and treating water. Share the message. -Dr. Picha |

| | | | | | |
|---|---|---|---|---|---|
| **13** | **Week 4** | **IVR** | Clean looking hands can have germs and handwashing with soap | Hello! This is Dr. Picha from the Provincial General Referral Hospital in Bukavu, I hope you and your family are well. I am here with Mwanza again today, the mother of a child that came to the CTC with diarrhea.<br>**Mwanza:** My baby got sick again. Ewww. What a nightmare! I had to bring him back to the CTC. He is only breastfeeding, so no water. I don't know how this happened! I wash my hands with soap.<br>**Dr. Picha:** When do you wash your hands with soap?<br>**Mwanza:** Whenever they look dirty!<br><br>For those listening, Mwanza needs your help. I would like to ask you a question. You can answer by pressing the 1 or 2 button on your phone. Your answer will be free. Should we wash our hands only when they look dirty? Press the 1 button on your phone if you think only dirty hands can have germs and microbes. Press 2 if you think that hands that look like clean can still have invisible germs and microbes. After you press 1 or 2, you will receive the correct answer. Again, your answer is free.<br><br>**If 1 was pressed:** Thanks for trying. Our hands can look clean but still have deadly invisible germs that can make our children and family very sick. It is important that we always wash our hands with soap before eating, preparing food, feeding a child, and after toileting or wiping or cleaning a child's feces. We will talk again soon.<br><br>**If 2 was pressed:** Excellent. Our hands can look clean but still have deadly invisible germs that can make our children and family very sick. It is important that we always wash our hands with soap before eating, preparing food, feeding a child, and after toileting or wiping or cleaning a child's feces. We will talk again soon.<br><br>**If no button was pressed:** We did not hear a response from you. Our hands can look clean but still have deadly invisible germs that can make our children and family very sick. It is important that we always wash our hands with soap before eating, preparing food, feeding a child, and after toileting or wiping or cleaning a child's feces. We will talk again soon. | Hands can look clean even though they contain diarrhea germs. We must always wash our hands using soapy water before eating and after touching feces. -Dr. Picha |
| **14** | **Week 5** | **Voice** | Safe water storage | Hello again! This is Dr. Picha from the Provincial General Referral Hospital in Bukavu. Did you know that once you treat your water it can become contaminated again? We still want to let you know that if our drinking water remains uncovered with a lid, dirt, germs or microbes and insects can fall into it. This is why we must cover our blue bucket used for storing drinking water every time. I would like to talk to you next week. Please share this message with your family. Stay well! | Treated drinking water can be contaminated again. If our drinking water remains uncovered without a lid, germs can fall into it. Keep your family healthy by covering your blue bucket with a lid. Share the message! -Dr. Picha |

| | | | | | |
|---|---|---|---|---|---|
| **15** | **Week 5** | **IVR** | How to prepare soapy water with detergent powder | Hello again, this is Dr. Picha from the Provincial General Referral Hospital in Bukavu. I am here with Mwanza again, the mother of a patient that came to the Provincial General Referral Hospital in Bukavu for treatment. She is running low on soapy water again and needs your help.<br>**Mwanza:** Can you help me make soapy water?<br><br>For those listening, I would like to ask you a question. You can answer by pressing the 1 or 2 button on your phone. Your answer will be free. How do you make soapy water for hand washing? Please press the 1 button on your phone if you put a full packet of detergent powder into a half-liter (500mL) bottle of water. Please press button 2 on your phone if you put 7 caps full of detergent powder into a half-liter (500mL) bottle of water.<br><br>**If 1 was pressed:** Thank you for trying. You must put 7 caps full of detergent powder into a 500 mL bottle of water. Remember, mix this well by shaking the bottle. To make a new bottle, make a small hole in the center of the lid with a nail, then start using the soapy water from this small hole to wash your hands. Share the message. We will talk again soon.<br><br>**If 2 was pressed:** Congratulations, this is correct. You must put 7 caps full of detergent powder into a 500 mL bottle of water. Remember, mix this well by shaking the bottle. To make a new bottle, make a small hole in the center of the lid with a nail, then start using the soapy water from this small hole to wash your hands. Share the message. We will talk again soon.<br><br>**If no button was pressed:** We did not hear a response from you. You must put 7 caps full of detergent powder into a 500 mL bottle of water. Remember, mix this well by shaking the bottle. To make a new bottle, make a small hole in the center of the lid with a nail, then start using the soapy water from this small hole to wash your hands. Share the message. We will talk again soon. | Powder detergent is great for making soapy water—just use 7 capfuls in any plastic bottle. It works as well as liquid or bar soap for getting rid of germs. Share the message. -Dr. Picha |
| **16** | **Week 6** | **Voice** | Encouraging male household members to wash hands with soap | Hello again! This is Dr. Picha from the Provincial General Referral Hospital in Bukavu. The male members of our family usually work very hard outside of the home. Thus, they may forget about washing hands with soapy water. Brothers, you should not forget this. Today, I have with me Mwanza's husband who is the father of the young child that came to the hospital with cholera.<br>**Dr. Picha:** Brother, how are you brother? How is Mwanza and your children?<br>**Mwanza's Husband:** Yes, good! My children had diarrhea often. Now, they remain healthy.<br>**Dr. Picha:** Why do you think they remain healthy now?<br>**Mwanza's Husband:** By drinking chlorinated water and washing hands with soapy water at key times as you instructed. These key times are before eating, before feeding our children, before preparing food, after cleaning a child's feces or anus, and after toileting. You called and texted us and I have shared these messages with my family and followed all the instructions.<br>**Dr. Picha:** Thank you Brother, thank you for listen today. Share the message. | It is important for every person, both men and women, to wash their hands with soap or ashes to protect themselves and their family members from severe diarrhea. Share the message! -Dr. Picha |

| | | | | | |
|---|---|---|---|---|---|
| **17** | **Week 6** | **IVR** | Handwashing with Ash | Hello! This is Dr. Picha from the Provincial General Referral Hospital in Bukavu. I hope you and your family are well. I am here with Mwanza again today, the mother of a child that came to the Provincial General Referral Hospital in Bukavu with diarrhea. Her family is facing a hard time and she wants to tell her story.<br>**Mwanza:** My husband lost his job, and now we barely have enough money to provide food for my children. We ran out of soapy water and don't have money for more. I am scared we will all get severe diarrhea again.<br>**Dr. Picha:** I am very sorry to hear this. Let's see how we can help you.<br><br>For those listening, I have a question; you can answer by pressing button 1 or button 2 on the phone. After pressing button 1 or button 2, you will be offered a correct answer. Your answer is free. Can ash be used if you do not have money to buy detergent powder or soap? Press 1 if you think ash can be used to replace detergent powder or soap for hand washing. Press 2 if you think ash cannot replace detergent powder or soap for handwashing. After you press 1 or 2, you will receive the correct answer. Again, your answer is free.<br><br>**If 1 was pressed:** Correct. Mwanza says thank you! Ash is just as effective as soap and soapy water at removing germs and dirt from our hands. Share the message. We will talk again soon.<br><br>**If 2 was pressed:** Thanks for trying. Ash is just as effective as soap and soapy water at removing germs and dirt from our hands. Share the message. We will talk again soon.<br><br>**If no button was pressed:** We did not hear a response from you. Ash is just as effective as soap and soapy water at removing germs and dirt from our hands. Share the message. We will talk again soon. | If you don't have soap, your family can wash their hands with ashes. Ash has the same effect of removing germs from hands as soap or soapy water. Share the message. -Dr. Picha |
| **18** | **Week 7** | **Voice** | Handwashing with soap | Hello again! This is Dr. Picha from the Provincial General Referral Hospital in Bukavu. We hope the whole family is healthy. We remind you that we will protect ourselves and our family members from severe diarrhea if we wash our hands at the following key times: before preparing food, before eating, before feeding the child, after using the toilet, and after wiping the child's stool. Be well and stay safe. | Protect your family by handwashing with soap: before preparing food, eating, and feeding children, and after toileting and wiping or cleaning a child's stool. Share the message. -Dr. Picha |

| | | | | | |
|---|---|---|---|---|---|
| **19** | **Week 7** | **IVR** | Everyone in your household should washing their hands with soap | Hello, this is Dr. Picha on the phone from the Provincial General Referral Hospital in Bukavu. It is important to remember that your household members are still vulnerable to cholera and diarrheal diseases. I am here with Mwanza, the mother of a patient that came to the CTC.<br>**Mwanza:** Oh no Dr. Picha! I always wash my hands with soap and boil our drinking water, but my husband and small son got severe diarrhea. How!<br>**Dr. Picha:** Did your whole family wash their hands with soap?<br>**Mwanza:** My husband is busy and is often outside the home. He doesn't have time to wash his hands with soap. He is using only water.<br><br>Please help Mwanza. You can answer by pressing the 1 or 2 button on your phone. Your answer will be free. Do those that are always outside the home during the day need to wash their hands with soap? Press 1 for Yes. Press 2 for No.<br><br>**If 1 was pressed:** Correct. All household members must always wash their hands with soap, even husbands that are outside the home most of the day for work. Mwanza's husband likely got severe diarrhea from not washing his hands with soap and spread this to the son. Share this message. We will talk again soon.<br><br>**If 2 was pressed:** Thanks for trying. All household members must always wash their hands with soap, even husbands that are outside the home most of the day for work. Mwanza's husband likely got severe diarrhea from not washing his hands with soap and spread this to the son. Share this message. We will talk again soon.<br><br>**If no button was pressed:** We did not hear a response from you. All household members must always wash their hands with soap, even husbands that are outside the home most of the day for work. Mwanza's husband likely got severe diarrhea from not washing his hands with soap and spread this to the son. Share this message. We will talk again soon. | Protect the health of your beloved children. All household members must always wash their hands with soap, even husbands who are outside the home most of the day. Share this message. -Dr. Picha |
| **20** | **Week 8** | **Voice** | Clean looking hands can have germs and handwashing with soap | Hello again! This is Dr. Picha from the Provincial General Referral Hospital in Bukavu. I hope you and your family are well. Did you know that severe diarrhea like cholera is spread through dirty hands? (Pause) Hands can look clean even though they contain diarrhea germs/microbes. That is why we must wash our hands at all times using soapy water to protect ourselves and our families from severe diarrhea. Remember that it is everyone's responsibility to keep the family healthy! | Hands can look clean even though they contain diarrhea germs. We must ash our hands using soapy water to protect our families from diarrhea. Share the message. -Dr. Picha |

| | | | | | |
|---|---|---|---|---|---|
| **21** | **Week 8** | **IVR** | Children with diarrhea should not sleep in the same bed with other children in the household | Hello, this is Dr. Picha again from the Provincial General Referral Hospital in Bukavu. I am here with Mwanza again today, the mother of a patient that came to the CTC. She wants to share her story.<br>**Mwanza:** All of my young children sleep together on a bed in the living room. When my youngest son first got diarrhea, he soiled the bed with feces in the middle of the night. I heard him crying and came to see what was happening. Feces was everywhere and got on my other children. After I brought my son to the hospital, my 4-year-old daughter also got severe diarrhea and a day later came to the hospital. Dr. Picha explained to me that I must not keep my child with diarrhea in the bed with my other children, since this can spread diarrhea quickly. Ewww. I didn't know and my babies got sick!<br><br>**Dr. Picha:** For those listening, I have a question for you. You can answer by pressing the 1 or 2 button on your phone. Your answer will be free.<br>What should you do if one of your children has diarrhea while they are in the bed with your other children? Press 1 on your phone if you think it is okay to keep the child with diarrhea in bed with your other children. Press 2 on your phone if you should move this child to the floor on a blanket.<br><br>**If 1 was pressed:** Thanks for trying. When a child has diarrhea, he or she should not sleep in the same bed with siblings. Place a blanket on the floor for the child to sleep if you do not have an extra bed. Wash the feces-soiled clothing, sheets, and blankets with soap. Share this message. We will talk again soon.<br><br>**If 2 was pressed:** Excellent. Correct. When a child has diarrhea, he or she should not sleep in the same bed with siblings. Place a blanket on the floor for the child to sleep if you do not have an extra bed. Wash the feces-soiled clothing, sheets, and blankets with soap. Share this message. We will talk again soon.<br><br>**If no button was pressed:** We did not hear a response from you. When a child has diarrhea, he or she should not sleep in the same bed with siblings. Place a blanket on the floor for the child to sleep if you do not have an extra bed. Wash the feces-soiled clothing, sheets, and blankets with soap. Share this message. We will talk again soon. | A child suffering from diarrhea can spread diarrhea to other children that sleep in the same bed. Keep children with diarrhea out of the bed with other children. Share the message. -Dr. Picha |
| **22** | **Week 9** | **Voice** | Children with diarrhea should not sleep in the same bed with other children in the household | Hello again! This is Dr. Picha from the Provincial General Referral Hospital in Bukavu. Do you remember Mwanza, the mother of a patient that came with her sick child to the cholera treatment center for severe diarrhea? She has a question about children in the same household sharing the same bed when one of them has diarrhea.<br>**Mwanza:** Dr. Picha! My child under five years old suffers from diarrhea. Is there a risk if he continues to share a bed with my other children?<br>**Dr. Picha:** Letting a child who suffers from cholera sleep with others children in the same bed exposes these other children to contracting severe diarrhea like cholera. When a child suffers from diarrhea, he should not sleep in the same bed with others children or with his parents. Share this message with other members of your family. Protect the health of your family. Stay healthy! | A child suffering from diarrhea can spread diarrhea to other children that sleep in the same bed. Keep children with diarrhea out of the bed with other children. Share the message. -Dr. Picha |

| | | | | | |
|---|---|---|---|---|---|
| **23** | **Week 9** | **IVR** | How to prepare a new handwashing station | Hello, this is Dr. Picha again from the Provincial General Referral Hospital in Bukavu. I am here with Mwanza again today, the mother of a patient that came to the CTC.<br>**Mwanza:** My children were playing and knocked over the handwashing station and the tap broke! Eww! What should I do?<br><br>**Dr. Picha:** To those listening, can you help Mwanza? I have a question for you. You can answer by pressing the 1 or 2 button on your phone. Your answer will be free. Can Mwanza buy materials in the market to fix her handwashing station? Press 1 on your phone if you think materials are available in the market in Bukavu to fix the broken handwashing station. Press 2 if you think materials cannot be found in the market in Bukavu to fix the broken handwashing station.<br><br>**If 1 was pressed:** Excellent, correct. If the tap on your handwashing station or drinking water vessel breaks, you can buy a new one for 6000 CF, and a 20-liter bucket for 7500 CF in most local markets. Using the knife, cut a very small hole near the bottom of the bucket. Make it bigger until you can screw in the tap. These materials are important to protect the health of you and your family. Fix these materials if they break. Share the message. We will talk again soon.<br><br>**If 2 was pressed:** Thanks for trying. If the tap on your handwashing station or drinking water vessel breaks, you can buy a new one for 6000 CF, and a 20-liter bucket for 7500 CF in most local markets. Using the knife, cut a very small hole near the bottom of the bucket. Make it bigger until you can screw in the tap. These materials are important to protect the health of you and your family. Fix these materials if they break. Share the message. We will talk again soon.<br><br>**If no button was pressed:** We did not hear a response from you. If the tap on your handwashing station or drinking water vessel breaks, you can buy a new one for 6000 CF, and a 20-liter bucket for 7500 CF in most local markets. Using the knife, cut a very small hole near the bottom of the bucket. Make it bigger until you can screw in the tap. These materials are important to protect the health of you and your family. Fix these materials if they break. Share the message. We will talk again soon. | If the tap on the bucket containing the hand washing water or the bucket of drinking water breaks, you can buy a new tap for 6000 CF and the 20 liter bucket for 7500 CF in various markets locally. Share the message. -Dr. Picha |
| **24** | **Week 10** | **Voice** | How to correctly handwashing with soap | Hello. It is Dr. Picha from the Provincial General Referral Hospital in Bukavu. I hope that your family is feeling well. I am with Mwanza, the mother of a young child that came to the hospital with cholera. An upsetting thing is that cholera and severe diarrhea come from germs in feces, and spreads through hands and water. To protect your family from these germs, make sure your whole family washes their hands with soap at 5 key times: after defecation, after cleaning a child's feces and anus, before preparing food, before eating, and before feeding children. Use soapy water each time! Help small children wash their hands with soap.<br>**Dr. Picha:** Mwanza, can you tell me how to properly wash my hands?<br>**Mwanza:** Yes! First, I must wet my hands and rub them together with soapy water. Then I rub my hands, palm and surface, between my fingers and under the nails! I keep this up for 20 seconds. When I'm done, I just rinse my hands with clean water and dry them with a clean dry towel. I also help small children do the same.<br>**Dr. Picha:** Thank you, Mwanza! That's absolutely right! You and Mwanza are starting to become experts on these things. Share this message. Remember, the health of your family is in your hands! | Make sure your whole family washes their hands with soap to protect them from diarrhea: Help small children who cannot reach the hand washing station themselves. Share the message. -Dr. Picha |

| | | | | | |
|---|---|---|---|---|---|
| **25** | **Week 10** | **IVR** | Importance of both handwashing with soap and water treatment | Hello again! This is Dr. Picha from the Provincial General Referral Hospital in Bukavu. Currently, is anyone in your household suffering from diarrhea? (Pause)<br>If someone in your household has diarrhea, now and for the next 7 days your family is still at high risk of suffering from severe diarrhea.<br><br>I would like to ask you a question: You can answer me by pressing button 1 or 2 on your phone. Pressing 1 or 2 will give you the correct answer. Your answer is free. How do we avoid severe diarrhea like cholera during the 7-day high-risk period after someone in your household has diarrhea? Please press 1 on your phone if you think handwashing with soap only is enough. Please press 2 on your phone if you think treating drinking water and washing hands with soap is required. Again, responding to this question is free.<br><br>**If 1 was pressed:** Thanks for trying. You must wash your hands with soap and treat your drinking water by boiling or using chlorine tablets to avoid severe diarrhea. If you follow Dr. Picha's advice, your family will be happy and healthy! We will talk again soon.<br><br>**If 2 was pressed:** Correct. You must wash your hands with soap and treat your drinking water by boiling or using chlorine tablets to avoid severe diarrhea. If you follow Dr. Picha's advice, your family will be happy and healthy! We will talk again soon.<br><br>**If no button was pressed**: We did not hear a response from you. You must wash your hands with soap and treat your drinking water by boiling or using chlorine tablets to avoid severe diarrhea. If you follow Dr. Picha's advice, your family will be happy and healthy! We will talk again soon. | Wash your hands with soap and treat drinking water by boiling it or using the chlorine tablet to prevent severe diarrhea. Stay happy and healthy! Share the message. -Dr. Picha |
| **26** | **Week 11** | **Voice** | Handwashing with soap helps prevent Ebola and COVID-19 | Hello, this is Dr. Picha again from the Provincial General Referral Hospital in Bukavu. I hope your family is well. I wanted to share a very important message. As we know we have been facing Ebola and coronavirus here in DRC. These diseases are deadly just like cholera and severe diarrhea. By washing our hands with soap before eating our food, feeding our children, and preparing our meals as well as after cleaning or wiping our child’s feces or toileting, we can protect our families and the children we love from Ebola and coronavirus. Always use your handwashing station and refill your soapy water bottle with 7 caps full of detergent powder and water. Keep your family healthy and happy. Share this important message with your family. Stay well. We will talk again soon. | Through washing our hands with soap before touching food and after touching feces, we can help protect our families and children from Ebola and coronavirus. Share this message. -Dr. Picha |

| | | | | | |
|---|---|---|---|---|---|
| **27** | **Week 11** | **IVR** | How to correctly boil water | Hello again! This is Dr. Picha from the Provincial General Referral Hospital in Bukavu. For today, I am here with Mwanza again, the mother of a patient that came to the CTC.<br>**Mwanza:** Dr. Picha, yesterday my young child was at my neighbor's house and he came home with severe diarrhea after drinking untreated water. What should I do?<br>**Dr. Picha:** I am so sorry to hear that your child became sick with severe diarrhea because your neighbors offered them untreated water. Our neighbors will appreciate us more if we give guests that come to our home chlorine-treated or boiled water.<br>**Mwanza:** I will tell my neighbors how to treat their water. I always use chlorine tablets to treat my water but some of my neighbors can't afford this. Can you remind me how to correctly boil water?<br><br>**Dr. Picha:** For those of you who are following us, can you help Mwanza? You can answer by pressing 1 or 2 on the phone. There is no charge for your answer. Press 1 on your phone if you think that to boil water correctly you should always boil for 30 minutes. Press 2 on your phone if you think you should wait until large bubbles rise up, a rolling boil.<br><br>**If 1 was pressed:** Thank you for trying! To treat your water by boiling you must always heat water until large bubbles rise up, a rolling boil. Share the message. Take care!<br><br>**If 2 was pressed:** That's right! To treat your water by boiling you must always heat water until large bubbles rise up, a rolling boil. Share the message. Take care!<br><br>**If no button was pressed**: We did not hear a response from you. To treat your water by boiling you must always heat water until large bubbles rise up, a rolling boil. Share the message. Take care! | Heat your water thoroughly so that it boils until big bubbles form. Visitors will be happy if we give them treated water. Please share this message. -Dr. Picha |
| **28** | **Week 12** | **Voice** | Safe water storage | Hello! This is Dr. Picha from the Provincial General Referral Hospital in Bukavu. We have some bad news to share today that affected another family like yours in this program. After 3 months in this program, they stopped using their red handwashing station and blue bucket for storing their drinking water. Their child was admitted to the hospital with diarrhea again this week and sadly died. Don't let this be your story - protect yourself and your family. Always fill your blue bucket with water and treat it with chlorine tablets or heat water until large bubbles form for boiling. Always keep the lid on your bucket. Also fill your red handwashing station and use bar or liquid soap, soapy water or ash to wash your hands to protect your family and children from severe diarrhea. Stay healthy! | After 3 months in this program, a family like yours stopped using their red handwashing station and blue bucket for storing their drinking water. Their child was admitted to the hospital with diarrhea again this week and died. Don't let this be your story! Protect yourself and your family from severe diarrhea. Always fill your blue bucket with water and use chlorine tablets or heat water until large bubbles form for boiling and fill your red handwashing station and use soap or ash. Share the message. -Dr. Picha |

| | | | | | |
|---|---|---|---|---|---|
| **29** | **Week 12** | **IVR** | Cleaning your drinking water vessel | Hello again! This is Dr. Picha from the Provincial General Referral Hospital in Bukavu. Today, I am here with Mwanza again, the mother of a patient that came to the Provincial General Referral Hospital in Bukavu for treatment.<br>**Mwanza:** Dr. Picha, can you remind me how often I need to wash the blue bucket for my drinking water?<br><br>**Dr. Picha:** For those who are listening, do you have any ideas to help Mwanza? You can answer by pressing 1 or 2 on your phone, and your answer will not cost you anything. You can press 1 on your phone if you think that you should wash your blue bucket with detergent or soap weekly. Or you can press 2 on your phone if you think that every time you get water for drinking you should wash your blue bucket with detergent or soap.<br><br>**If 1 was pressed:** You've given it a good try! Every time you get water for drinking, you should wash your blue bucket with detergent or soap. Don't forget to share this message with other members of your family. Let's all remember that our health is in our hands! I will call you again.<br><br>**If 2 was pressed:** 100%. Correct! Every time you get water for drinking, you should wash your blue bucket with detergent or soap. Don't forget to share this message with other members of your family. Let's all remember that our health is in our hands! I will call you again.<br><br>**If no button was pressed:** We did not hear a response from you. Every time you get water for drinking, you should wash your blue bucket with detergent or soap. Don't forget to share this message with other members of your family. Let's all remember that our health is in our hands! I will call you again. | Every time you get water for drinking you should wash your blue bucket with detergent or soap. Share this message. Let's all remember that our health is in our hands! -Dr. Picha |
| **30** | **Week 13** | **Voice** | Water treatment | Hello! I am Dr. Picha from the Provincial General Referral Hospital in Bukavu. We know that sometimes it is difficult to get water in the two buckets we gave you because it was difficult to find water at your tap for your drinking water. Go and draw water frequently, after pouring water from this jerrycan into the blue bucket add a single chlorine tablet and wait 30 minutes. Share the message. | We know that sometimes it is difficult to fill your blue bucket. Go and draw water frequently, after pouring water from this jerrycan into the blue bucket add a single chlorine tablet and wait 30 minutes. Share the message. -Dr. Picha |

| | | | | | |
|---|---|---|---|---|---|
| **31** | **Week 13** | **IVR** | Volume of water needed for water treatment using chlorine tablets | Hello! I am Dr. Picha from the Provincial General Referral Hospital in Bukavu. As we all know, it is difficult to always have water in the drinking water storage bucket and in the hand washing station because sometimes there is no water in the tap where you get your drinking water. I wanted to ask you a question, and you can answer by pressing 1 or 2 on your phone. Your answer will not cost you anything.<br><br>How much water do you need in your blue drinking water vessel before you add an chlorine tablet? If you think you can prepare chlorine treated water if you have 10 liters of water, press 1. If you think you can prepare chlorine treated water if you have 20 liters of water, press 2.<br><br>**If 1 was pressed:** Thank you very much for trying. You need at least 20 liters of water to treat your drinking water using the chlorine tablets we gave you. We know that sometimes it is difficult to find enough water to fill your blue drinking water storage container. Use your 20-liter jerry can to store extra water when it is available so you can use this when there is no water coming out of your tap. Stay healthy and happy. Share the message.<br><br>**If 2 was pressed:** Thank you very much! Great job. You need at least 20 liters of water to treat your drinking water using the chlorine tablets we gave you. We know that sometimes it is difficult to find enough water to fill your blue drinking water storage container. Use your 20-liter jerry can to store extra water when it is available so you can use this when there is no water coming out of your tap. Stay healthy and happy. Share the message.<br><br>**If no button was pressed:** Unfortunately we did not receive your answer, the correct answer is: You need at least 20 liters of water to treat your drinking water using the chlorine tablets we gave you. We know that sometimes it is difficult to find enough water to fill your blue drinking water storage container. Use your 20-liter jerry can to store extra water when it is available so you can use this when there is no water coming out of your tap. Stay healthy and happy. Share the message. | We know that sometimes it is difficult find enough water to fill your blue drinking water storage container. Use your 20 liter jerrycan to store extra water when it is avaliable so you can use this when there is no water coming out of your tap. Stay healthy. Share the message. -Dr. Picha |
| **32** | **Week 14** | **Voice** | Handwashing with ash is free and effective | **Dr. Picha:** Hello once again! I am Dr. Picha from the Provincial General Referral Hospital in Bukavu. We are here with Mwanza, the mother of a little child who came to the hospital because of severe diarrhea. We are discussing the importance of washing our hands during key moments. Mwanza has a question for me about how to keep her hands and those of her children clean when she doesn't have the money to buy soap.<br>**Mwanza:** Dr. Picha, for the past two days, my children and I have been washing our hands with water only because it was difficult to find money to buy soap or detergent. I wanted to know if there is anything else we can do to wash our hands when we are without soap and detergent?<br>**Dr. Picha:** When we don't have soap available in our house, we can use ash to make our hands clean because ash doesn't cost us anything and plays the same role as soap in making our hands clean. yYou and your family will be healthy if you do so. Use ash to stay safe, healthy, and happy. Share the message. | When we do not have soap at home, we should use ash to wash our hands because ash does not require any cost and plays the same role as soap to clean our hands properly. Share this message. -Dr. Picha |

| | | | | | |
|---|---|---|---|---|---|
| **33** | **Week 14** | **IVR** | Ash is free and effective for handwashing | Hello again! I am Dr. Picha from the Provincial General Referral Hospital in Bukavu. We are here with Mwanza, the mother of a young child who came to the hospital with severe diarrhea. We are discussing the importance of hand washing with soap during key moments. Mwanza has a question for me about how to clean her hands and those of her children when she has no money to buy soap.<br>**Mwanza:** Dr. Picha, for the past two days, my children and I have been washing our hands with water only because it has been difficult to get money to buy soap or detergent. I would like to know if there is another way to wash our hands when we do not have soap or detergent.<br><br>**Dr. Picha:** For the one who is listening! Can you help Mwanza? You can press 1 or 2 on your phone to answer. Your answer will not cost you anything. If you think ash can be used as a free, low-cost effective alternative to soap, press 1. If you think water is the best option if you don't have money for soap, press 2.<br><br>**If 1 was pressed:** Congratulations! When we lack soap in our house, we can use ash to clean our hands because ash does not incur any cost and plays the same role in cleaning our hands properly. You and your family members will be healthy if you do this. Be healthy. Share the message.<br><br>**If 2 was pressed:** Thanks for trying! When we lack soap in our house, we can use ash to clean our hands because ash does not incur any cost and plays the same role in cleaning our hands properly. You and your family members will be healthy if you do this. Be healthy. Share the message.<br><br>**If no button was pressed:** By bad luck, we did not have your answer! When we lack soap in our house, we can use ash to clean our hands because ash does not incur any cost and plays the same role in cleaning our hands properly. You and your family members will be healthy if you do this. Be healthy. Share the message. | It happens sometimes that it can be hard to find even 200 CF to buy soap or detergent to wash your hands at home. You can use ash to wash your hands because ash does not cost money and plays the same role as soap in cleaning our hands properly. Share this message. -Dr. Picha |
| **34** | **Week 15** | **Voice** | Correct placement of WASH photo cue cards on handwashing with soap, water treatment, and how to prepare a handwashing station | Hello! I am Dr. Picha from the Provincial General Referral Hospital in Bukavu. I am here with Mwanza, the mother of a little child who came to the hospital because of severe diarrhea.<br>**Mwanza:** Dr. Picha, your team came to visit me at home and brought me materials such as the drinking water storage container, the hand washing station, soapy water, a jerrycan to draw drinking water, and so on. I would like to know why they also brought us different pictures.<br>**Dr. Picha:** Aaah! These photos are very important, especially for visitors and other members of the household who were absent during the family teachings. They help to show how to treat drinking water, the key moments of washing hands with soap, and they show how to place the tap on the blue and red buckets.<br>**Mwanza:** Thank you very much Dr. Picha for making me understand<br>**Dr. Picha:** Put each of them in the right place as shown by my team to always remind you on how to treat water and the key moments to wash your hands with soap. Your family's health is in your hands. Share the message. | The photos we brought you are very important, especially for visitors and other household members who were absent during the family teachings. They help to show how to treat drinking water and the key moments of washing hands with soap. Make sure these are visible in your home to your family and guests. Be careful. -Dr. Picha |

| | | | | | |
|---|---|---|---|---|---|
| **35** | **Week 15** | **IVR** | Please use keep cue cards for handwashing and water treatment on the walls in your home | Hello again! This is Dr. Picha from the Provincial General Referral Hospital in Bukavu. I am here with Mwanza, the mother of a small sick child, who came to the hospital for treatment of severe diarrhea. She wanted to know something.<br>**Mwanza:** Dr. Picha, your team visited my family and brought a drinking water storage container, the hand washing station, the soapy water, the blue bucket for storing drinking water, and photos of handwashing with soap and water treatment. We would like to understand why these photos are so important for our health.<br><br>Your family also received these photos like Mama Mwanza's. Can you help Mama Mwanza by explaining to her the importance of these photos that my team gave her? You can answer by pressing 1 or 2 on your phone. Your answer is free. If you think that these photos show you how to create a new handwashing station or drinking water vessel if your current one is broken, press 1. If you think these photos show you how to feed your children healthy food, press 2.<br><br>**If 1 was pressed:** Congratulations! These photos are very important to keep your family members and visitors healthy. These photos remind you and your family to treat drinking water with chlorine tablets, the key moments of hand washing with soap, and how to put a tap on the blue bucket and the red bucket. Attach these photos to your wall to protect that health of you and your family. Share the message. I will call you again!<br><br>**If 2 was pressed:** Thanks for trying. These photos are very important to keep your family members and visitors healthy. These photos remind you and your family to treat drinking water with chlorine tablets, the key moments of hand washing with soap, and how to put a tap on the blue bucket and the red bucket. Attach these photos to your wall to protect that health of you and your family. Share the message. I will call you again!<br><br>**If no button was pressed:** Too bad you haven't given your answer yet! The correct answer is: These photos are very important to keep your family members and visitors healthy. These photos remind you and your family to treat drinking water with chlorine tablets, the key moments of hand washing with soap, and how to put a tap on the blue bucket and the red bucket. Attach these photos to your wall to protect that health of you and your family. Share the message. I will call you again! | These photos are very important to keep your family members and visitors healthy. These photos remind you and your family to treat drinking water with chlorine tablets, the key moments of hand washing with soap, and how to put a tap on the blue bucket and the red bucket. Attach these photos to your wall to protect that health of you and your family. Share the message. I will call you again! -Dr. Picha |
| **36** | **Week 16** | **Voice** | Chlorine tablets are available at pharmacies in your area | Hello! Dr. Picha from Provincial General Referral Hospital in Bukavu. We want to remind you that not treating our drinking water puts us and our young children at risk for severe diarrheal diseases. If it is not possible to find chlorine tablets at pharmacies in our neighborhood, we can get them in pharmacies in the city center for 200 CF per one tablet. Keep your family healthy and happy. Share the message. Thank you! | We would like to remind you that not continuing to treat drinking water exposes us and our young children to severe diarrheal diseases. Therefore, if chlorine tablets are not available in your area, you can buy these in the city center for only 200 CF per one tablet. Keep your family healthy. Share the message. -Dr. Picha |

| | | | | | |
|---|---|---|---|---|---|
| **37** | **Week 16** | **IVR** | Many diarrhea patient households in your area have returned to the health facility | Hello! This is Dr. Picha from the Provincial General Referral Hospital in Bukavu. I would like to remind you of one thing, but before I do, I have a question. You can answer by pressing 1 or 2 from your phone. There is no cost for responding. Not treating drinking water with chlorine tablets or not boiling water can put you and your children at risk of returning to the health facility with diarrhea. If you think to stay free from diarrhea it is okay for you and your children to sometimes drink untreated water as long as most of your water is treated, press 1. If you think to stay free from diarrhea you and your children must always drink treated water, press 2.<br><br>**If 1 was pressed:** Thanks for trying! Many diarrhea patient households in your area have returned to the health facility because their child had diarrhea from drinking untreated water. Protect your family and children by always drinking treated water using chlorine tablets or by boiling water. It is not okay to sometimes drink untreated water. Even a little untreated water can make you and your child sick and force you to return to the health facility. This would be very costly for your family. You can protect yourself, your family, and your children from severe diarrheal diseases by always putting water in the blue bucket, using it only for drinking, and not using it for any other purpose in the house. Remember that your health and that of your family and children is in your hands. Share the message!<br><br>**If 2 was pressed:** Thank you! Many diarrhea patient households in your area have returned to the health facility because their child had diarrhea from drinking untreated water. Protect your family and children by always drinking treated water using chlorine tablets or by boiling water. It is not okay to sometimes drink untreated water. Even a little untreated water can make you and your child sick and force you to return to the health facility. This would be very costly for your family. You can protect yourself, your family, and your children from severe diarrheal diseases by always putting water in the blue bucket, using it only for drinking and not using it for any other purpose in the house. Remember that your health and that of your family and childre is in your hands. Share the message!<br><br>**If no button was pressed:** So far, we haven't received your call! Many diarrhea patient households in your area have returned to the health facility because their child had diarrhea from drinking untreated water. Protect your family and children by always drinking treated water using chlorine tablets or by boiling water. It is not okay to sometimes drink untreated water. Even a little untreated water can make you and your child sick and force you to return to the health facility. This would be very costly for your family. You can protect yourself, your family, and your children from severe diarrheal diseases by always putting water in the blue bucket, using it only for drinking, and not using it for any other purpose in the house. Remember that your health and that of your family and children is in your hands. Share the message! | Many diarrhea patient households in your area have returned to the health facility because their child got diarrhea from drinking untreated water. This would be very costly for your family. Protect your family and children by always treating drinking water using chlorine tablets or by boiling water and always storing treated water in your blue bucket. Remember that your health and that of your children and family is in your hands. Share the message! -Dr. Picha |
| **38** | **Week 17** | **Voice** | Correctly using WASH package materials | Hello! Dr. Picha from the Provincial General Referral Hospital in Bukavul here. Some households like yours have broken their stools for their handwashing stations and water vessels because they used them as chairs or put heavy items on them. These stools are not strong enough for sitting. You need to use your hardware to stay healthy so please always use the stool we gave you only for your handwashing station and drinking water vessel. Keep your family healthy and happy. Share the message. | The stools we gave you are not strong enough to sit on. You need to use your hardware to stay healthy, so please always use the stools we gave you only for your handwashing station and drinking water vessel, not for sitting. Keep your family healthy and happy. Share this message. -Dr. Picha |

| | | | | | |
|---|---|---|---|---|---|
| **39** | **Week 17** | **IVR** | Please maintain well your handwashing station | Hello again! This is Dr. Picha from the Provincial General Referral Hospital in Bukavu. I am here with Mwanza, the mother of a small sick child, who came to the hospital for treatment of severe diarrhea. She has a question!<br>**Mwanza:** Dr. Picha! My children were sitting on the stools you gave us and broke them! I feel so bad now that I did not respect the message you sent about not using the stool you provided for sitting. How can I continue to use my handwashing station?<br><br>For those who are listening! Can you help Mwanza? You can answer by pressing 1 or 2 on your phone and your answer will not be charged. If you think Mwanza can put the hand washing station directly on the floor, press 1. If you think Mwanza can use small stool (kasonga) to put under the hand washing station, press 2.<br><br>**If 1 was pressed:** Thanks for trying! Your stools are fragile and are meant to only hold the handwashing station and water vessel and are not meant for sitting. If your children break your stool, you can use a small stool (kasonga). It is important you continue to use your handwashing station to washing your hands with soap or ash to keep your family healthy and happy. Share the message.<br><br>**If 2 was pressed:** Congratulations! Your stools are fragile and are meant to only hold the handwashing station and water vessel, and are not meant for sitting. If your children break your stool, you can use a small stool (kasonga). It is important you continue to use your handwashing station to wash your hands with soap or ash to keep your family healthy and happy. Share the message.<br><br>**If no button was pressed:** We didn't receive your reply. Your stools are fragile and are meant to only hold the handwashing station and water vessel, and are not meant for sitting. If your children break your stool, you can use a small stool (kasonga). It is important you continue to use your handwashing station to wash your hands with soap or ash to keep your family healthy and happy. Share the message. | Your stools are fragile and are meant only to hold the handwashing station and water vessel and are not meant for sitting. If your children break your stool you can use a small stool (kasonga) instead. It is important you continue to use your handwashing station to wash your hands with soap or ash to keep your family healthy and happy. Share the message. -Dr. Picha |
| **40** | **Week 18** | **Voice** | Maintenance for WASH package materials | Hello once again! I am Dr. Picha from the Provincial General Referral Hospital in Bukavu. I would like to remind you that it is important to make good use of the materials that our team gave you to continue to keep your family healthy by drinking water treated with chlorine tablets every time and by washing your hands during key moments. Take care to not break these materials and clean them frequently. Remember that your family's health is a priority. If you do break them, you can prepare new ones using the instructions on the picture we gave you. These materials can be found at the market for a low price. Stay healthy. Share the message. | Remember that your family's health is a priority. Take care to not break the materials we gave you and clean them frequently. If you do break them, you can prepare new ones using the instructions on the picture we gave you. These materials can be found at the market for a low price. Share this message with your family members. -Dr. Picha |

| | | | | | |
|---|---|---|---|---|---|
| **41** | **Week 18** | **IVR** | How to properly use your handwashing station | Hello! This is Dr. Picha from the Provincial General Referral Hospital in Bukavu. I am here with Mwanza, the mother of a small sick child, who came to the hospital for treatment of severe diarrhea.<br>**Mwanza:** Dr. Picha, my child came back to the health facility yet again.<br>**Dr. Picha:** I am so sorry your child is sick again. What happened? Did you keep using your handwashing station with the basin?<br>**Mwanza:** You know we have a problem with space in our household. Our place is small, and I have 8 children. I could not find a place for the basin, so I now keep it in the kitchen for dishes.<br>**Dr. Picha:** Mwanza, using your handwashing station and basin is important to make sure you and everyone in your household wash their hands regularly.<br>**Mwanza:** What should I do?<br>For you on the call, please help Mwanza. You can answer by pressing 1 or 2 from your phone. There is no charge to respond. If you think Mwanza is out of luck and can do nothing, press 1. If you think Mwanza can put the basin on top the handwashing station when no one is using it to save space, press 2.<br><br>**If 1 was pressed:** Thanks for trying! It is important to use the materials that my team gave you correctly. Always keep your basin under or on top of your handwashing station, and always keep water inside for handwashing, and soap or ash to keep your children from coming back to the hospital with diarrhea. Always wash your hands with soap or ash. Stay healthy and happy. Share the message.<br><br>**If 2 was pressed:** Thank you! That's a good answer! It is important to use the materials that my team gave you correctly. Always keep your basin under or on top of your handwashing station, and always keep water inside for handwashing, and soap or ash to keep your children from coming back to the hospital with diarrhea. Always wash your hands with soap or ash. Stay healthy and happy. Share the message.<br><br>**If no button was pressed:** We didn't hear a response from you. It is important to use the materials that my team gave you correctly. Always keep your basin under or on top of your handwashing station, and always keep water inside for handwashing, and soap or ash to keep your children from coming back to the hospital with diarrhea. Always wash your hands with soap or ash. Stay healthy and happy. Share the message. | It is important to use the materials that my team gave you correctly. Always keep your basin under or on top of your handwashing station, and always keep water inside for handwashing and soap or ash nearby. Keep your children from coming back to the hospital with diarrhea by always washing your hands with soap or ash. Stay healthy and happy. Share the message. -Dr. Picha |
| **42** | **Week 19** | **Voice** | Importance of sustained handwashing with soap and water treatment | Hello! I am Dr. Picha from the Provincial General Referral Hospital in Bukavu. I am here with Mwanza, the mother of a little child who came to the hospital because of severe diarrhea. She has a question!<br>**Mwanza:** Dr. Picha! My child was sick, and after getting teachings from your team, we treated drinking water and washed our hands for the 7 high risk days. After these seven days, my child had again fallen ill with severe diarrhea and returned to the hospital. What do you think happened?<br>**Dr. Picha:** I am sorry to hear this Mother Mwanza! It appears that after the seven days of risk, your family did not continue to treat their drinking water with chlorine tablets or wash their hands with soap after using the toilet, after cleaning up the child's feces, before eating, before feeding the child, and before preparing the food.<br>**Mwanza:** That's right, Dr. Picha! After those seven days of risk, we thought there was no more risk. We drank untreated water and did not wash our hands with soap during key moments.<br>**Dr. Picha:** Other families like yours where we visited, continue with the practice of treating their drinking water and washing their hands with soap even after the seven-day high-risk period, they are healthy and their children are growing normally and have not had severe diarrhea. Therefore, continue to treat your drinking water and wash your hands with soap during key moments so that your family can continue to live free from severe diarrhea. I'll call you later. Be careful. Share the message. | If we and our families and don't continue to wash our hands with soap, treat our drinking water with chlorine tablets, and safely store our water after the 7 day high risk period, we and our young children can become sick like Mwanza. Stay healthy. Share the message. -Dr. Picha |

| | | | | | |
|---|---|---|---|---|---|
| **43** | **Week 19** | **IVR** | Boiling is just as effective as chlorine tablets in reducing microbes | Hello! This is Dr. Picha from the Provincial General Referral Hospital in Bukavu. I am here with Mwanza, the mother of a small sick child, who came to the hospital for treatment of severe diarrhea.<br>**Mwanza:** Dr. Picha! My money is really tight right now because my husband lost his job, and we have 8 children. We no longer buy chlorine tablets to treat our drinking water. What should we do? I want to keep my children and family healthy. We don't want to go to the health facility again.<br>**Dr. Picha:** Please help Mwanza. You can answer by pressing 1 or 2 on your phone. Your answer is free of charge. Do you think boiling can be as effective as using a chlorine tablet in reducing microbes? If you think boiling can be as effective as chlorine tablets in reducing microbes, press 1. If you think boiling is not as effective as chlorine tablets in reducing microbes, press 2.<br><br>**If 1 was pressed:** Congratulations! Yes, boiling is just as effective as chlorine tablets in reducing microbes. So, if you don't have money for more chlorine tablets, boil your water using the leftover wood or coal from cooking. Boil water until large bubbles form, then cover the water and let it cool. Once cool, pour this into your bucket with the lid. Practice this to keep your family healthy and happy. Share the message!<br><br>**If 2 was pressed:** Thanks for trying! Boiling is just as effective as chlorine tablets in reducing microbes. So, if you don't have money for more chlorine tablets, boil your water using the leftover wood or coal from cooking. Boil water until large bubbles form, then cover the water and let it cool. Once cool, pour this into your bucket with the lid. Practice this to keep your family healthy and happy. Share the message!<br><br>**If no button was pressed:** We didn't hear a response. Boiling is just as effective as chlorine tablets in reducing microbes. So, if you don't have money for more chlorine tablets, boil your water using the leftover wood or coal from cooking. Boil water until large bubbles form, then cover the water and let it cool. Once cool, pour this into your bucket with the lid. Practice this to keep your family healthy and happy. Share the message! | Boiling is just as effective as chlorine tablets in reducing microbes. If you don't have money for more chlorine tablets, boil your water using the leftover wood or coal from cooking. Boil water until large bubbles form, then cover the water and let it cool. Once cool pour this into your bucket with the lid. Practice this to keep your family health and happy. Share the message! -Dr. Picha |
| **44** | **Week 20** | **Voice** | Importance of using your handwashing station | Hello! I am Dr. Picha from the Provincial General Referral Hospital in Bukavu. I am here with Mwanza, the mother of a little child who came to the hospital because of severe diarrhea. She had a question!<br>**Mwanza:** Dr. Picha, why is it so important to use the handwashing station you gave us vs. just using a basin?<br>**Dr. Picha:** Mother Mwanza! There are two main benefits of the handwashing station. It is easy to dispense water to wash hands, and we don't dip our hands in the water others used to clean their hands which can make ourselves and our children sick. Use the hand washing station every time during key moments to always stay healthy. Share the message. Stay safe. | Use the handwashing station to make it easy for family members to wash their hands with soap. When you are using it, it is easy to pour water to wash your hands, the basin prevents water from being poured on the floor, and it is easy for children to use. Keep your family healthy every day. Share the message. -Dr. Picha |

| | | | | | |
|---|---|---|---|---|---|
| **45** | **Week 20** | **IVR** | Importance of properly using your handwashing station | Hello! This is Dr. Picha from the Provincial General Referral Hospital in Bukavu. I am calling all patient households to let them know that we have an increasing number of children returning to the hospital yet again for diarrhea in your area. This is very alarming. Parents are having to pay high fees at the health facility for their care. I would like to ask you a question. You can answer by pressing 1 or 2 from your phone. There will be no charge for your answer. Do you currently have water in your red bucket? If you have water in the red bucket we gave you press 1. If you do not have water in your red bucket, press 2.<br><br>**If 1 was pressed:** Congratulations! Please always keep water in your red bucket and the basin underneath. Many of the households that returned to the health facility because their child had diarrhea were not using our handwashing station. Some even had beans or dishes inside. Oh no! To keep your family and children healthy, everyone, including children, must wash their hands with soap before eating and preparing food and after toileting or cleaning a child's feces. If you don't have soap, ash works just as well. Not washing your hands with soap or ash during key moments can put you and your children at risk of getting sick from severe diarrhea again. Be healthy and happy. Share the message.<br><br>**If 2 was pressed:** Thank you! Please always keep water in your red bucket and the basin underneath. Many of the households that returned to the health facility because their child had diarrhea were not using our handwashing station. Some even had beans or dishes inside. Oh no! To keep your family and children healthy, everyone, including children, must wash their hands with soap before eating and preparing food and after toileting or cleaning a child's feces. If you don't have soap, ash works just as well. Not washing your hands with soap or ash during key moments can put you and your children at risk of getting sick from severe diarrhea again. Be healthy and happy. Share the message.<br><br>**If no button was pressed:** So far, we have not received your answer! Please always keep water in your red bucket and the basin underneath. Many of the households that returned to the health facility because their child had diarrhea were not using our handwashing station. Some even had beans or dishes inside. Oh no! To keep your family and children healthy, everyone, including children, must wash their hands with soap before eating and preparing food and after toileting or cleaning a child's feces. If you don't have soap, ash works just as well. Not washing your hands with soap or ash during key moments can put you and your children at risk of getting sick from severe diarrhea again. Be healthy and happy. Share the message. | Many households in your area have returned to the health facility because their child had severe diarrhea because they were not using our handwashing station. Some even had beans or dishes inside the hand washing station. Oh no! To keep your family and children healthy, everyone including children must what their hands with soap before eating and preparing food and after toileting or cleaning a child's feces. If you don't have soap, ash works just as well. Not washing your hands with soap or ash during key moments can put you and your children at risk of getting sick from severe diarrhea again. Be healthy and happy. Share the message. -Dr. Picha |
| **46** | **Week 21** | **Voice** | Importance of children handwashing with soap | Hello! Dr. Picha from the Provincial General Referral Hospital in Bukavu here. I wanted to remind you that just as you should wash your hands with soap before serving your child food. Children should also wash their hands before taking food to eat to protect themselves from severe diarrhea. This also includes when you give your child small foods such as bread or donuts and not just when you give him or her food such as fufu or other foods. Be careful. Share this message with other members of your family. | We should always wash our hands with soap before serving our children food. Children should also wash their hands before taking that food to eat to protect themselves from severe diarrhea. This also includes when we give our children small foods such as bread or donuts and not just when you give him or her food such as fufu. Stay healthy, share the message. -Dr. Picha |

| | | | | | |
|---|---|---|---|---|---|
| **47** | **Week 21** | **IVR** | There have been many diarrhea patients admitted to health facilities in your area, mostly children | Hello again! This is Dr. Picha from the Provincial General Referral Hospital in Bukavu. We are reaching out to you with a grave warning. There have been many diarrhea patients admitted to health facilities in your area, mostly children. Even households receiving this very intervention are returning because they stopped practicing. Listen closely to learn how to protect your family. I will start with a question for you.<br><br>What can we do to avoid going back to the health facility because our children contract severe diarrhea? If you think using chlorine tablets only to treat your drinking water is enough to protect your family from severe diarrhea, press 1. If you think all household members must wash their hands with soap or ash, in addition to using chlorine tablets to treat your drinking water, to protect your family from severe diarrhea press 2.<br><br>**If 1 was pressed:** Thanks for trying! There is another major severe diarrhea outbreak in your area. Protect yourself and your children by making sure everyone in your household is washing their hands with soap or ash before eating and after toileting, as well as treating your water using chlorine tablets or boiling. Keep your children and family healthy and happy, and don't forget to share the message!<br><br>**If 2 was pressed:** Congratulations! There is another major severe diarrhea outbreak in your area. Protect yourself and your children by making sure everyone in your household is washing their hands with soap or ash before eating and after toileting, as well as treating your water using chlorine tablets or boiling. Keep your children and family healthy and happy, and don't forget to share the message!<br><br>**If no button was pressed:** We didn't hear a response from you. There is another major severe diarrhea outbreak in your area. Protect yourself and your children by making sure everyone in your household is washing their hands with soap or ash before eating and after toileting, as well as treating your water using chlorine tablets or boiling. Keep your children and family healthy and happy, and don't forget to share the message! | There is another major severe diarrhea outbreak in your area. Protect yourself and your children by making sure everyone in your household is washing their hands with soap or ash before eating and after toileting, as well as treating your water using chlorine tablets or boiling. Keep your children and family health and happy. Share the message! -Dr. Picha |
| **48** | **Week 22** | **Voice** | Fathers, brothers, uncles, and grandpas always wash their hands with soap before eating and after using the toilet | Hello! Dr. Picha from the Provincial General Referral Hospital in Bukavu here. It is important that fathers, brothers, uncles, and grandpas always wash their hands with soap before eating and after using the toilet otherwise they can make their precious children sick. We know our Congolese men are strong but they can have an infection without symptoms and make their family sick including their little children. Brothers listen to my words and keep your family healthy and happy. Share the message. | It is important that fathers, brothers, uncles, and grandpas always wash their hands with soap before eating and after using the toilet otherwise they can make their precious children sick. We know our Congolese men are strong but they can have an infection without symptoms and make their family sick including their little children. Stay healthy, share the message. -Dr. Picha |

| | | | | | |
|---|---|---|---|---|---|
| **49** | **Week 22** | **IVR** | Importance of children washing their with soap | Hello! This is Dr. Picha from the Provincial General Referral Hospital in Bukavu. I hope you and your family are well. I have a question! You can press 1 or 2 from your phone. We are seeing many children with severe diarrhea at the health facilities in your area. Do you think it is still important for your children to wash their hands with soap or ash before eating a donut to prevent severe diarrhea? If you think handwashing with soap or ash is not necessary before eating a donut, press 1. If you think handwashing with soap or ash is necessary before eating a donut, press 2.<br><br>**If 1 was pressed:** Thanks for trying! Many mothers of children with diarrhea in the health facilities thought that it was not necessary to have their children wash their hands with soap or ash before eating a donut. Their young children got sick with severe diarrhea, and their household had to pay the high hospital bills. You and your children should always wash your hands with soap or ash before eating, even when it’s a donut! Share the message!<br><br>**If 2 was pressed:** Congratulations! Many mothers of children with diarrhea in the health facilities thought that it was not necessary to have their children wash their hands with soap or ash before eating a donut. Their young children got sick with severe diarrhea, and their household had to pay the high hospital bills. You and your children should always wash your hands with soap or ash before eating, even when it’s a donut! Share the message!<br><br>**If no button was pressed:** We did not hear an answer from you! Many mothers of children with diarrhea in the health facilities thought that it was not necessary to have their children wash their hands with soap or ash before eating a donut. Their young children got sick with severe diarrhea, and their household had to pay the high hospital bills. You and your children should always wash your hands with soap or ash before eating, even when it’s a donut! Share the message! | Many mothers of children with diarrhea in the health facilities thought that it was not needed to have their children wash their hands with soap or ash before eating a donut. Their young children got sick with severe diarrhea and their household had to pay the high hospital bills. You and your children should always wash your hands with soap or ash before eating even when its a donut! Share the message! -Dr. Picha |
| **50** | **Week 23** | **Voice** | Water treatment and Handwashing | Hello! Dr. Picha from the Provincial General Referral Hospital in Bukavu here. We all know how our children love to play. They get into everything. Sometimes they even play with the handwashing station and take the soapy water bottle as a toy. To prevent this, keep the soapy water bottle on the string we gave you. Children also sometimes play with the water in the basin. To stop this, empty the handwashing water from the basin frequently. Follow my words to keep your family health and happy. Share the message. | Children love to play, even with the soapy water bottle. To prevent this, keep the soapy water bottle on the string we gave you. Also empty your basin frequently to keep children from playing with this water. Stay healthy, share the message. -Dr. Picha |

| | | | | | |
|---|---|---|---|---|---|
| **51** | **Week 23** | **IVR** | Importance of handwashing with soap and water treatment even after the 7-day high risk period | Hello again! This is Dr. Picha from the Provincial General Referral Hospital in Bukavu. I wanted to ask you a question! You can answer by pressing 1 or 2 on your phone. Your answer is free! After leaving the hospital, can we get sick with severe diarrhea again if we don't wash our hands with soap and if we don't treat our drinking water with chlorine tablets or boil it? If you think that washing your hands with soap and treating drinking water with chlorine tablets or boiling water should be a practice for only the 7-day high-risk period, press 1. If you think that washing your hands and treating drinking water with chlorine tablets or boiling water should be a practice for every day of your life, press 2.<br><br>**If 1 was pressed:** Thanks for trying! Washing hands with soap and treating drinking water with chlorine tablets or boiling water should be our daily practice to make sure our family lives free from severe diarrhea. Let's not be like Mama Mwanza who had a child return to the hospital because of severe diarrhea because her family stopped practicing. One of my team members came to the hospital and to her home to teach her. It was observed that Mwanza and her family had continued to practice the teachings, but after 7 days they were no longer washing their hands with soap and treating their drinking water. Unfortunately, after two weeks, her child Jean-Claude was again admitted to the hospital with severe diarrhea. Keep your family healthy every time so that you don't have the same problem as Mama Mwanza's family. Share the message!<br><br>**If 2 was pressed:** Good answer! Thank you very much! Washing hands with soap and treating drinking water with chlorine tablets or boiling water should be our daily practice to make sure our family lives free from severe diarrhea. Let's not be like Mama Mwanza who had a child return to the hospital because of severe diarrhea because her family stopped practicing. One of my team members came to the hospital and to her home to teach her. It was observed that Mwanza and her family had continued to practice the teachings, but after 7 days they were no longer washing their hands with soap and treating their drinking water. Unfortunately, after two weeks, her child Jean-Claude was again admitted to the hospital with severe diarrhea. Keep your family healthy every time so that you don't have the same problem as Mama Mwanza's family. Share the message!<br><br>**If no button was pressed:** We did not receive your reply. Washing hands with soap and treating drinking water with chlorine tablets or boiling water should be our daily practice to make sure our family lives free from severe diarrhea. Let's not be like Mama Mwanza who had a child return to the hospital because of severe diarrhea because her family stopped practicing. One of my team members came to the hospital and to her home to teach her. It was observed that Mwanza and her family had continued to practice the teachings, but after 7 days they were no longer washing their hands with soap and treating their drinking water. Unfortunately, after two weeks, her child Jean-Claude was again admitted to the hospital with severe diarrhea. Keep your family healthy every time so that you don't have the same problem as Mama Mwanza's family. Share the message! | Washing hands with soap and treating drinking water with chlorine tablets or boiling water should be our daily practice to make sure our family lives free from severe diarrhea. Always keep your family healthy so that you don't have the same problem as Mama Mwanza's family. Share the message! -Dr. Picha |
| **52** | **Week 24** | **Voice** | Handwashing with soap | Hello! Dr. Picha from the Provincial General Referral Hospital in Bukavu here. Remember our handwashing song [audio of handwashing song]. Sing this to your children to encourage them to wash their hands with soap. Share the message to keep your family health and happy. | Handwashing with soap before eating and after toileting keeps our families healthy and happy. Share the message. -Dr. Picha |

| | | | | | |
|---|---|---|---|---|---|
| **53** | **Week 24** | **IVR** | How to boil and safely store drinking water | Hello again! This is Dr. Picha from the Provincial General Referral Hospital in Bukavu. Congratulations for reaching 3 months in our intervention! You are becoming an expert. I would like to remind you of one thing through a question! You can press 1 or 2 on your phone. Your answer is free of charge. How should you boil your water? If you think you should boil your water for 30 minutes, press 1. If you think you should boil until the bubbles in the water get big, press 2.<br><br>**If 1 was pressed:** Thanks for trying! Boil your water until big bubbles form then cover the pot and let it cool. When the water is cool, pour this into your blue bucket and put the lid on. When we drink water that was not treated with chlorine tablets or was not boiled, it can cause severe diarrhea and even death for our young children. Therefore, you and your children must always drink treated water to protect your family from diarrhea. You never know when a big diarrhea outbreak may occur in your area. Don't be the one with their child in the hospital. Share the message to keep your child and your family healthy and happy.<br><br>**If 2 was pressed:** Congratulations! Correct. Boil your water until big bubbles form then cover the pot and let it cool. When the water is cool, pour this into your blue bucket and put the lid on. When we drink water that was not treated with chlorine tablets or was not boiled, it can cause severe diarrhea and even death for our young children. Therefore, you and your children must always drink treated water to protect your family from diarrhea. You never know when a big diarrhea outbreak may occur in your area. Don't be the one with their child in the hospital. Share the message to keep your child and your family healthy and happy.<br><br>**If no button was pressed:** We didn't hear any answer from you! Boil your water until big bubbles form then cover the pot and let it cool. When the water is cool, pour this into your blue bucket and put the lid on. When we drink water that was not treated with chlorine tablets or was not boiled, it can cause severe diarrhea and even death for our young children. Therefore, you and your children must always drink treated water to protect your family from diarrhea. You never know when a big diarrhea outbreak may occur in your area. Don't be the one with their child in the hospital. Share the message to keep your child and your family healthy and happy. | Boil your water until big bubbles form and then cover the pot and let it cool. When the water is cool, pour this into your blue bucket and put the lid on. When we drink water that is not treated with chlorine tablets or is not boiled, it can cause severe diarrhea and even death for our young children. Therefore, you and your children must always drink treated water to protect yourselves and your family from diarrhea. You never know when a big diarrhea outbreak may occur in your area. Don't be the one with their child in the hospital. Share the message to keep your child and your family healthy and happy. -Dr. Picha |
| **54** | **Week 25** | **Voice** | Importance of handwashing with soap for fathers | Hello! This is Dr. Picha from the Provincial General Referral Hospital in Bukavu. We know that most of the time fathers don't stay at home. And we all know that severe diarrhea spares no one, whether you are a dad or mom, rich or poor. It is a disease that attacks everyone. But when the father comes home at night, it is very important that he also washes his hands with soap before eating and after toileting to protect himself and his family from severe diarrhea. Fathers can spread severe diarrhea to their young children without even being sick. And as the head of the family, the father is also responsible for educating his family members to wash their hands with soap or ash before eating and toileting to protect themselves from severe diarrhea. Keep your family safe from severe diarrhea. | Severe diarrhea spares no one, whether you are a dad or mom, rich or poor. It is a disease that attacks everyone. But when the father comes home at night, it is very important that he also washes his hands with soap before eating and after toileting to protect himself and his family from severe diarrhea. Fathers can spread severe diarrhea to their young children without being sick themselves. Keep your family safe from severe diarrhea. -Dr. Picha |

| | | | | | |
|---|---|---|---|---|---|
| **55** | **Week 25** | **IVR** | Importance of fathers and brothers washing their hands with soap | Hello! This is Dr. Picha from the Provincial General Referral Hospital in Bukavu. This message is for my fathers and brothers. I have a question! Is handwashing with soap or ash before eating and after toileting as important for fathers and brothers as for our mothers and sisters? You can answer by pressing 1 or 2 on your phone. There is no charge for answering this question. If you think handwashing with soap or ash before eating and after toileting is more important for mothers and sisters, press 1. If you think handwashing with soap or ash before eating and after toileting is equally important for all, press 2.<br><br>**If 1 was pressed:** Thanks for trying! Handwashing with soap or ash before eating and after toileting is equally important for all. We know that most of the time, fathers and brothers don't stay at home during the day. But it is important that they wash their hands with soap or ash before eating and after toileting both in and outside their home. Fathers and brothers can carry severe diarrhea germs even when they are not sick and spread this to their children and family, so protect yourself and your family by always washing your hands with soap or ash before eating and after toileting. Keep your family healthy. Share the message!<br><br>**If 2 was pressed:** Thank you very much. Good answer! Handwashing with soap or ash before eating and after toileting is equally important for all. We know that most of the time, fathers and brothers don't stay at home during the day. But it is important that they wash their hands with soap or ash before eating and after toileting both in and outside their home. Fathers and brothers can carry severe diarrhea germs even when they are not sick and spread this to their children and family, so protect yourself and your family by always washing your hands with soap or ash before eating and after toileting. Keep your family healthy. Share the message!<br><br>**If no button was pressed:** By bad luck we had not yet received your answer! Handwashing with soap or ash before eating and after toileting is equally important for all. We know that most of the time, fathers and brothers don't stay at home during the day. But it is important that they wash their hands with soap or ash before eating and after toileting both in and outside their home. Fathers and brothers can carry severe diarrhea germs even when they are not sick and spread this to their children and family, so protect yourself and your family by always washing your hands with soap or ash before eating and after toileting. Keep your family healthy. Share the message! | We know that most of the time fathers and brothers don't stay at home during the day. But it is important that they wash their hands with soap or ash before eating and after toileting both in and outside their home. Fathers and brothers can carry severe diarrhea germs even when they are not sick and spread this to their children and family, so protect yourself and your family by always washing your hands with soap or ash before eating and after toileting. Share the message! -Dr. Picha |
| **56** | **Week 26** | **Voice** | Handwashing with soap or ash | Hello! This is Dr. Picha from the Provincial General Referral Hospital in Bukavu. We are here with Mwanza and she has a question.<br>**Mwanza:** Dr. Picha! Yesterday my children had diarrhea. I would like to know what happened again. I always treat my water by boiling or chlorine tablets.<br>**Dr. Picha:** Aah! Mama Mwanza! It seems that your food may have diarrhea germs. Have you been consistently washing your hands with soap or ash before preparing your food?<br>**Mwanza:** Oh Dr. Picha! I try but many times I forget. I am just so busy that sometimes I forget to wash my hands with soap or ash before preparing food for my family. I need to remember this so my little ones don't get sick again.<br>**Dr. Picha:** Yes. It is important to wash your hands with soap or ash when preparing food to prevent diarrhea germs from getting into the food. Share the message with your family members. | It is important to wash your hands with soap or ash before preparing food to prevent diarrhea germs from getting into your food which can make your children and family sick with severe diarrhea. Share the message with your family members. -Dr. Picha |

| | | | | | |
|---|---|---|---|---|---|
| **57** | **Week 26** | **IVR** | Handwashing with soap before food preparation to prevent severe diarrhea | Hello! This is Dr. Picha from the Provincial General Referral Hospital in Bukavu. I hope you and your family are well. Now I would like to ask you a question. You can answer by pressing 1 or 2 on your phone. There is no charge for your answer! Who in your household should wash their hands with soap or ash before preparing food? If you think that only the mother has the obligation to wash her hands with soap or ash before preparing food, press 1. If you think that washing hands with soap or ash is an obligation of everyone involved in the preparation of food for your family, press 2.<br><br>**If 1 was pressed:** Thank you very much for trying! It is very important that mothers and everyone involved in the preparation of food for the family wash their hands with soap or ash before preparing food to prevent the spread of severe diarrhea germs to your children and other family members. Even men are responsible for reminding household members to wash their hands with soap or ash when preparing food. This is important for the health of your family. Share the message!<br><br>**If 2 was pressed:** Thank you! Good answer! It is very important that mothers and everyone involved in the preparation of food for the family wash their hands with soap or ash before preparing food to prevent the spread of severe diarrhea germs to your children and other family members. Even men are responsible for reminding household members to wash their hands with soap or ash when preparing food. This is important for the health of your family. Share the message!<br><br>**If no button was pressed:** By bad luck, we had not yet received your answer! It is very important that mothers and everyone involved in the preparation of food for the family wash their hands with soap or ash before preparing food to prevent the spread of severe diarrhea germs to your children and other family members. Even men are responsible for reminding household members to wash their hands with soap or ash when preparing food. This is important for the health of your family. Share the message! | It is very important that mothers and everyone involved in the preparation of food for the family wash their hands with soap or ash before preparing food to prevent the spread of severe diarrhea germs to children and other family members. Even men are responsible for reminding household members to wash their hands with soap or ash when preparing food. This is important for the health of your family. Share the message! -Dr. Picha |
| **58** | **Week 27** | **Voice** | Child feces contains diarrhea germs | Hello again! This is Dr. Picha from the Provincial General Referral Hospital in Bukavu. I would like to remind you again today that washing your hands is a good way to keep your children and family happy and healthy. Did you know that the stool of a child with diarrhea contains germs just like the stool of an adult? Therefore, make every effort to wash your hands with soap or ash after cleaning a child's stool, because not washing your hands with soap or ash can spread severe diarrhea to other members of your family. Be careful, and remember that the health of your family is in your hands. | Stool of a child with diarrhea contains germs like the stool of an adult. Therefore, make every effort to wash your hands with soap or ash after cleaning the child's stool, because not washing your hands with soap or ash can spread severe diarrhea to other members of your family. Be careful and remember that the health of your family is in your hands. -Dr. Picha |

| | | | | | |
|---|---|---|---|---|---|
| **59** | **Week 27** | **IVR** | The feces of young children contain diarrhea germs | Hello! This is Dr. Picha from the Provincial General Referral Hospital in Bukavu. I am here with Mwanza, the mother of a little child who had came to the hospital for severe diarrhea. She has a question!<br>**Mwanza:** Dr. Picha! Your team came to visit me this week and told me that it is important to wash hands with soap or ash after cleaning my child's stool. This wasn't clear to me since I thought children's stool did not contain microbes. Can you help me understand this?<br><br>For those who are listening! You can help Mwanza. Does the stool of young children contain microbes? To answer, you can press 1 or 2 on your phone. There is no cost for your answer. If you think the stool of young children does not contain microbes, press 1. Or If you think the stool of young children does contain microbes, press 2.<br><br>**If 1 was pressed:** Thank you very much for trying! The stool of a small child also has germs like those of an adult person. That is why it is very important to always wash your hands with soap or ash after cleaning the child's stool, even if your child is not sick. Children without diarrhea can still have many germs in their stool which can be spread to other children and family members, causing severe diarrhea. Share the message!<br><br>**If 2 was pressed:** Congratulations! Good answer! The stool of a small child also has germs like those of an adult person. That is why it is very important to always wash your hands with soap or ash after cleaning the child's stool, even if your child is not sick. Children without diarrhea can still have many germs in their stool which can be spread to other children and family members, causing severe diarrhea. Share the message!<br><br>**If no button was pressed:** So far we don't have your answer! The correct answer is: the stool of a small child also has germs like those of an adult person. That is why it is very important to always wash your hands with soap or ash after cleaning the child's stool, even if your child is not sick. Children without diarrhea can still have many germs in their stool which can be spread to other children and family members, causing severe diarrhea. Share the message! | The stool of a small child also has germs like those of an adult person. That is why it is very important to always wash your hands with soap or ash after cleaning the child's stool even your child is not sick. Children without diarrhea can still have many germs in their stool which can be spread to other children and family members which can cause severe diarrhea. Share the message! -Dr. Picha |
| **60** | **Week 28** | **Voice** | Handwashing with soap after cleaning child's anus | Dr. Picha: Hello! This is Dr. Picha from the Provincial General Referral Hospital in Bukavu. I am here with Mwanza, the mother of this little child who had come to the hospital for severe diarrhea. She has a question!<br>**Mwanza:** Dr. Picha! When your team visited the hospital and my household, the health promoter told me that I should wash my hands with soap or ash after cleaning my child's feces. But I did not see the importance of washing my hands with soap or ash if I already rinsed my hands and my child's anus with water.<br>**Dr. Picha:** Ooh, Mama Mwanza! Even if you wash your child's anus and your hands with water, severe diarrhea germs can remain on your hands which can make you and your other children and family members sick. Again, you will remember that the germs of severe diarrhea are not visible to the naked eye. Always wash your hands with soap after cleaning your child's anus. Share the message! | Even if you wash your child's anus and your hands with water, severe diarrhea germs can remain on your hands which can make you and your other children and family members sick. Again, you will remember that the germs of severe diarrhea are not visible to the naked eye. Always wash your hands with soap or ash after cleaning your child's anus. Share the message! -Dr. Picha |

| | | | | | |
|---|---|---|---|---|---|
| **61** | **Week 28** | **IVR** | Handwashing with soap after child feces disposal | Hello again! This is Dr. Picha from the Provincial General Referral Hospital in Bukavu. I hope you and your family are well. This is an important message since there has been an increase in diarrhea patients in your area. Is it okay to wash my hands with water only after cleaning my child's anus? To answer, you can press 1 or 2 on your phone. If you think that it is okay to use water only to wash your hands after cleaning a child's anus, press 1. If you think you need to always use soap or ash to wash your hands after cleaning a child's anus, press 2.<br><br>**If 1 was pressed:** Thanks for trying! Washing your hands with soap or ash after cleaning your child's anus or removing your child's stool is the only way to protect against severe diarrhea. Drinking treated water only is not enough. You must be careful now since there are many severe diarrhea cases in your area. Keep your family safe from severe diarrhea by washing your hands with soap or ash after cleaning your child's anus and disposing of your child's stool, even if you used a pot or a spade. Be careful! Share the message!<br><br>**If 2 was pressed:** Thank you very much! It's true! Washing your hands with soap or ash after cleaning your child's anus or removing your child's stool is the only way to protect against severe diarrhea. Drinking treated water only is not enough. You must be careful now since there are many severe diarrhea cases in your area. Keep your family safe from severe diarrhea by washing your hands with soap or ash after cleaning your child's anus and disposing of your child's stool, even if you used a pot or a spade. Be careful! Share the message!<br><br>**If no button was pressed:** We have not yet received your answer! The correct answer is: Washing your hands with soap or ash after cleaning your child's anus or removing your child's stool is the only way to protect against severe diarrhea. Drinking treated water only is not enough. You must be careful now since there are many severe diarrhea cases in your area. Keep your family safe from severe diarrhea by washing your hands with soap or ash after cleaning your child's anus and disposing of your child's stool, even if you used a pot or a spade. Be careful! Share the message! | Washing your hands with soap or ash after cleaning your child's anus or removing your child's stool is the only way to protect against severe diarrhea. Water only is not enough. You must be careful now since there are many severe diarrhea cases in your area. Keep your family safe from severe diarrhea by washing your hands with soap or ash after cleaning your child's anus and disposing of your child's stool, even if you used a pot or a spade. Be careful! Share the message! -Dr. Picha |
| **62** | **Week 29** | **Voice** | Handwashing with soap | Hello again! This is Dr. Picha from the Provincial General Referral Hospital in Bukavu. The message today is for our young children. My team and I have noticed that many of the children in our program like those in your family are not following my recommendation to wash hands with ash or soap before eating. They rush to eat because they are so hungry. I visited a household last week with children like this. They never washed their hands with soap or ash before eating. Two of these children had to come back to the Provincial General Referral Hospital in Bukavu with severe diarrhea. This was a very bad situation. It turns out that they were playing with other children in the neighborhood who also had come to the hospital with severe diarrhea. Please to keep your children healthy and happy tell them and watch over them to make sure they always wash their hands with soap or ash before eating. Share this message! | Last week, our team had a household with children that never washed their hands with soap or ash before eating. Two of these children had to come back to the Provincial General Referral Hospital in Bukavu with severe diarrhea. It turns out that they were playing with other children in the neighborhood that also had come to the hospital with severe diarrhea. Keep your children healthy and happy by telling them to always wash their hands with soap or ash before eating. Share this message! -Dr. Picha |

| | | | | | |
|---|---|---|---|---|---|
| **63** | **Week 29** | **IVR** | Don't keep your soap in the bedroom away from children keep it available for handwashing | Hello again! This is Dr. Picha from the Provincial General Referral Hospital in Bukavu. I am here again with Mwanza, the mother of this little child who had come to the hospital for severe diarrhea. She has a question!<br><br>**Mwanza:** Dr. Picha! My husband and I are always giving our family treated water and washing our hands with soap before eating and feeding our children and after using the toilet but my young son just got sick again from diarrhea. What is happening!!!<br>**Dr. Picha:** Are your children washing their hands with soap after coming from the toilet?<br>**Mwanza:** I try but they don't listen! What should I do?<br>You can help Mwanza! You can press 1 or 2 on your phone to answer the question. Your answer will not cost anything. Do you think Mwanza should make sure ash or soap is always available for her children to use for handwashing? If you think Mwanza should always put ash or soap next to her handwashing station for her children to wash their hands, press 1. If you think that Mwanza should always keep the soap in her bedroom to keep her children from playing with it, press 2.<br><br>**If 1 was pressed:** Congratulations! You must always keep ash or soap next to your handwashing station and keep water in your handwashing station so your children can wash their hands and stay healthy and happy! Many mothers keep their soap in the bedroom so their children don't waste it. If your soap is low, keep ash next to your handwashing station. Encourage your children to wash their hands with ash before eating and after using the toilet. Don't be stuck bringing your child to the hospital for diarrhea. Share the message!<br><br>**If 2 was pressed:** Thanks for trying! You must always keep ash or soap next to your handwashing station and keep water in your handwashing station so your children can wash their hands and stay healthy and happy! Many mothers keep their soap in the bedroom so their children don't waste it. If your soap is low, keep ash next to your handwashing station. Encourage your children to wash their hands with ash before eating and after using the toilet. Don't be stuck bringing your child to the hospital for diarrhea. Share the message!<br><br>**If no button was pressed:** Now we didn't get your answer! You must always keep ash or soap next to your handwashing station and keep water in your handwashing station so your children can wash their hands and stay healthy and happy! Many mothers keep their soap in the bedroom so their children don't waste it. If your soap is low, keep ash next to your handwashing station. Encourage your children to wash their hands with ash before eating and after using the toilet. Don't be stuck bringing your child to the hospital for diarrhea. Share the message! | You must always keep ash or soap next to your handwashing station and keep water in your handwashing station so your children can wash their hands and stay healthy and happy! Many mothers keep their soap in the bedroom so their children don't waste it. If your soap is low, keep ash next to your handwashing station. Encourage your children to wash their hands with ash before eating and after toilet. Don't be stuck bringing your child to the hospital for diarrhea. Share the message! -Dr. Picha |
| **64** | **Week 30** | **Voice** | Handwashing with soap after urinating | Hello again! This is Dr. Picha from the Provincial General Referral Hospital in Bukavu. As we said in the past days, the microbes of severe diarrhea are not visible to the naked eye and have no smell. I would like to remind you that the toilet is a place where we find thousands of germs. Therefore, we should always wash our hands with soap or ash after using the toilet, even if it's just to urinate. Just as we should wash our hands during defecation, it would be important to wash our hands with soap or ash after urinating. Share this message! | The toilet is a place where we find thousands of germs. Therefore, we should always wash our hands with soap or ash after using the toilet, even if it's just to urinate. Just as we should wash our hands during defecation, it would be important to wash our hands with soap or ash after urinating. Share this message! -Dr. Picha |

| | | | | | |
|---|---|---|---|---|---|
| **65** | **Week 30** | **IVR** | Importance of handwashing with soap after urination | Hello! This is Dr. Picha from the Provincial General Referral Hospital in Bukavu. Fathers and mothers, please listen to my words. Many don't remember that the toilet is a place where we have thousands of germs. I have a question! You can answer by pressing 1 or 2 on your phone, and your answer will not be charged any fee. Do I need to wash my hands with ash or soap if I only urinate? If you think there is no need to wash your hands with ash or soap after urinating, press 1. If you think you must wash your hands with ash or soap after urinating, press 2.<br><br>**If 1 was pressed:** Thanks for trying! To protect yourself, your children, and your family from severe diarrhea, it is important to wash your hands with soap or ash after using the toilet, even when you only urinate. This behavior should be practiced after urinating every time in every day. Be vigilant and remember to tell your children and family members to do this every time. Share the message.<br><br>**If 2 was pressed:** Congratulations! To protect yourself, your children, and your family from severe diarrhea, it is important to wash your hands with soap or ash after using the toilet, even when you only urinate. This behavior should be practiced after urinating every time in every day. Be vigilant and remember to tell your children and family members to do this every time. Share the message.<br><br>**If no button was pressed:** By bad luck we didn't get your answer! To protect yourself, your children, and your family from severe diarrhea, it is important to wash your hands with soap or ash after using the toilet, even when you only urinate. This behavior should be practiced after urinating every time in every day. Be vigilant and remember to tell your children and family members to do this every time. Share the message. | To protect yourself, your children, and your family from severe diarrhea, it is important to wash your hands with soap or ash after using the toilet, even when you only urinate. This behavior should be every time in every day. Be vigilant and remember to tell your children and family members to do this every time you use the toilet. Share the message. -Dr. Picha |
| **66** | **Week 31** | **Voice** | Keep your chlorine tablets for your own household members | Hello! This is Dr. Picha from the Provincial General Referral Hospital in Bukavu. Drinking water that is free of germs is very important for your health and the health of your family. It is very important to treat your family's drinking water with chlorine tablets. These tablets should only be used in your household and should not be shared with family members or even neighbors. If your neighbor or family member asks for chlorine tablets, you can tell them that these tablets are available in pharmacies and are not expensive. Stay healthy. | The chlorine tablets that we gave you are important for your health. These tablets should only be used in your household and should not be shared with family members or even neighbors. If your neighbor or family member asks for chlorine tablets, you can tell them that these tablets are available in pharmacies and are not expensive. Stay healthy. -Dr. Picha |

| | | | | | |
|---|---|---|---|---|---|
| 67 | **Week 31** | **IVR** | Please not share the chlorine tablets we provided with your neighbors | Hello again! This is Dr. Picha from the Provincial General Referral Hospital in Bukavu. I am here with Mwanza, the mother of a little child who came to the hospital for severe diarrhea. She has a question!<br>**Mwanza:** Dr. Picha! A neighbor arrived at my house and asked me to share with him the chlorine tablets that your team gave me recently. He told me that he also needs to treat his family's drinking water as I still do. I don't know what to do! Can I share these tablets with him?<br><br>You can help Mwanza! You can press 1 or 2 on your phone to answer the question. Your answer will not cost anything. Do you think Mwanza should share the chlorine tablets with other family members or neighbors? If you think Mwanza should share the chlorine tablets with her neighbor so that he can also treat his drinking water, press 1. If you think that Mwanza shouldn't share the chlorine tablets we gave her with her neighbor, press 2.<br><br>**If 1 was pressed:** Thanks for trying! Please keep your chlorine tablets for the treatment of your family's drinking water so your children and family can stay safe from diarrhea. This means that you should not share the chlorine tablets we provided with your neighbors. Instead, if your neighbor also needs chlorine tablets for the treatment of his family's drinking water, you can explain to him that these tablets are available in pharmacies in your neighborhood and are sold at a low cost. Stay healthy! Share the message.<br><br>**If 2 was pressed:** Thank you! Good answer! Please keep your chlorine tablets for the treatment of your family's drinking water so your children and family can stay safe from diarrhea. This means that you should not share the chlorine tablets we provided with your neighbors. Instead, if your neighbor also needs chlorine tablets for the treatment of his family's drinking water, you can explain to him that these tablets are available in pharmacies in your neighborhood and are sold at a low cost. Stay healthy! Share the message.<br><br>**If no button was pressed:** Now we didn't get your answer! Please keep your chlorine tablets for the treatment of your family's drinking water so your children and family can stay safe from diarrhea. This means that you should not share the chlorine tablets we provided with your neighbors. Instead, if your neighbor also needs chlorine tablets for the treatment of his family's drinking water, you can explain to him that these tablets are available in pharmacies in your neighborhood and are sold at a low cost. Stay healthy! Share the message. | Please not share the chlorine tablets we provided with your neighbors. Please keep your chlorine tablets for the treatment of your family's drinking water so your children and family can stay safe from diarrhea. Instead if your neighbor also needs chlorine tablets for the treatment of his family's drinking water, you can explain to him that these tablets are available in pharmacies in your neighborhood and are sold at a low cost. Stay healthy! Share the message. - Dr. Picha |
| 68 | **Week 32** | **Voice** | Conserve water for handwashing | Hello again! This is Dr. Picha from the Provincial General Referral Hospital in Bukavu. We hope your family is doing well! We know that currently some families have difficulty finding water that they can use for hand washing with ash or soap or even for drinking water. Remember to use your extra jerrycan for water storage when water is available. Conserve water for handwashing by turning off your tap after you wet your hands and are rub them together. Be vigilant! Share the message to stay healthy and happy. | We know that currently some families have difficulty finding water that they can use for hand washing with ash or soap or even for drinking water. Remember to use your extra jerrycan for water storage when water is avaliable. Conserve water for handwashing by turning off your tap after you wet your hands and are rubbing them together. Be vigilant! Share the message to stay healthy and happy. - Dr. Picha |

| | | | | | |
|---|---|---|---|---|---|
| **69** | **Week 32** | **IVR** | Always refill your handwashing station and drinking water vessel | Hello again! This is Dr. Picha from the Provincial General Referral Hospital in Bukavu. I hope you and all your family are well! We know that at the moment it is difficult to find water to put in your red and blue buckets. I would like to ask you a question! You can answer by pressing 1 or 2 on your phone. Your answer is free. What can we do to have treated water for drinking and to wash our hands with ash and soap when water is so hard to find? If you think we should no longer wash our hands with soap or ash or drink clean water because water has become hard to find, press 1. If you think we should use our extra jerrycan provided to store water when water is available, press 2.<br><br>**If 1 was pressed:** Thanks for trying! Even if water is hard to find, we should strive to put water in both of our buckets to reassure ourselves that the family is washing their hands with soap or ash and drinking treated water. Please use the jerrycan we provided to store extra water when it's available. We should use a small amount of water when we wash our hands with ash or soap by turning off the tap when we are not using it. And for drinking water, we can boil water using the embers left over from food preparation if we do not have enough water to fill our bucket. If it is difficult to do this, we can take a small amount of boiled water used for preparing foufou to give to small children because they are the ones most affected by diarrhea. Share this message with your family! I wish you well!<br><br>**If 2 was pressed:** Great job. Correct! Even if water is hard to find, we should strive to put water in both of our buckets to reassure ourselves that the family is washing their hands with soap or ash and drinking treated water. Please use the jerrycan we provided to store extra water when it's available. We should use a small amount of water when we wash our hands with ash or soap by turning off the tap when we are not using it. And for drinking water, we can boil water using the embers left over from food preparation if we do not have enough water to fill our bucket. If it is difficult to do this, we can take a small amount of boiled water used for preparing foufou to give to small children because they are the ones most affected by diarrhea. Share this message with your family! I wish you well!<br><br>**If no button was pressed:** Until now we have not received your answer! Even if water is hard to find, we should strive to put water in both of our buckets to reassure ourselves that the family is washing their hands with soap or ash and drinking treated water. Please use the jerrycan we provided to store extra water when it's available. We should use a small amount of water when we wash our hands with ash or soap by turning off the tap when we are not using it. And for drinking water, we can boil water using the embers left over from food preparation if we do not have enough water to fill our bucket. If it is difficult to do this, we can take a small amount of boiled water used for preparing foufou to give to small children because they are the ones most affected by diarrhea. Share this message with your family! I wish you well! | Even if water has become hard to find, we should strive to put water in both of our buckets to reassure ourselves that the family is washing their hands with soap or ash and drinking treated water. We should use a small amount of water when we wash our hands with ash or soap by turning off the tap when we are not using it. And for drinking water, we can boil water using the embers left over from food preparation if we do not have enough water to fill our bucket. If it is difficult to do this, we can take a small amount of boiled water used for preparing foufou to give to small children because they are the ones most affected by diarrhea. Please also use the jerry can we provided to store extra water when it's available. Share this message with your family! -Dr. Picha |

| | | | | | |
|---|---|---|---|---|---|
| **70** | **Week 33** | **Voice** | Message sharing and water treatment | Hello again! This is Dr. Picha from the Provincial General Referral Hospital in Bukavu. My message today is for my brothers. We know you are working hard to support your children. We applaud you. If my calls come while you are outside your home, please remember to share this information with your wives, children, and other household members. My messages are important to keep you and your family healthy and happy. Please always treat your water with chlorine tablets. You can buy these from the local market when you're coming home from work. Chlorine tablets cost 200 CF per tablet and will keep you and your children safe from severe diarrhea. Share the message! Have a good afternoon! | We know you are working hard to support your children. We applaud you. If my calls come while you are outside your home, please remember to share this information with your wives, children, and other household members. My messages are important to keep you and your family healthy and happy. Please always treat your water with chlorine tablets - you can buy these from the local market when you're coming home from work. Chlorine tablets cost 200 CF per tablet and will keep you and your children healthy from severe diarrhea. Share the message! Have a good afternoon! -Dr. Picha |
| **71** | **Week 33** | **IVR** | Clean your handwashing station and drinking water vessel at once per week | Hello again! This is Dr. Picha from the Provincial General Referral Hospital in Bukavu. We hope you and your family are all well. Today again I have one question to ask you. You can answer by pressing 1 or 2 on your phone. After pressing 1 or 2, you will receive the correct answer. Your answer will not cost you anything. How often should you wash your handwashing station and drinking water vessel we provided with soap and water? If you think every day, press 1. If you think weekly, press 2.<br><br>**If 1 was pressed:** Thanks for trying! It is very important to clean the bucket used for hand washing and the one used for storing drinking water weekly with soap and water so that they are no longer an entry point for germs in your family. If they are dirty, they will not only look unpleasent in your household, but can also cause germs to build up. Your family will stay healthy by keeping these materials clean - so clean them weekly with soap and water. Stay well! Share the message.<br><br>**If 2 was pressed:** Congratulations! It is very important to clean the bucket used for hand washing and the one used for storing drinking water weekly with soap and water so that they are no longer an entry point for germs in your family. If they are dirty, they will not only look unpleasent in your household, but can also cause germs to build up. Your family will stay healthy by keeping these materials clean - so clean them weekly with soap and water. Stay well! Share the message.<br><br>**If no button was pressed:** By bad luck, we haven't received your answer! The correct answer is: it is very important to clean the bucket used for hand washing and the one used for storing drinking water weekly with soap and water so that they are no longer an entry point for germs in your family. If they are dirty, they will not only look unpleasent in your household, but can also cause germs to build up. Your family will stay healthy by keeping these materials clean - so clean them weekly with soap and water.Stay well! Share the message. | It is very important to clean the bucket used for hand washing and the one used for storing drinking water weekly with soap and water so that they are no longer an entry point for germs in your family. And when they are dirty they will look nice in your household and could cause germs to build up. Your family will be healthy every time by keeping these materials clean by cleaning them weekly with soap and water. Stay well! Share the message. -Dr. Picha |

| | | | | | |
|---|---|---|---|---|---|
| **72** | **Week 34** | **Voice** | Locations of pharmacies with chlorine tablets in your area | Hello! This is Dr. Picha from the Provincial General Referral Hospital in Bukavu. Today we are here with Mwanza, the mother of the little child who came to the hospital for treatment of severe diarrhea. She says she had some questions about the chlorine tablets for treating her drinking water.<br>**Mwanza:** Dr. Picha! Since the chlorine tablets that you gave me for water treatment were finished, we have not treated our drinking water. What can we do to get these tablets?<br>**Dr. Picha:** Maman Mwanza! I am sorry to hear that nowadays, you and your family are drinking untreated water because you cannot find where to buy the chlorine tablets. Chlorine tablets for water treatment are available in pharmacies in Bukavu. You can find them in PANZI, ESSENCE, KADUTU, NYAWERA, NGUBA, PLACE, BRASSERIE and BAGIRA. It is up to you now to see which place is close by. It costs 200 CF per tablet that treats 20 liters of water. You will remember that it takes money to get these tablets but it will take a lot more money to pay for the care for severe diarrhea. Be vigilant! Share the message to keep your family healthy and happy. | Chlorine tablets for water treatment are available in pharmacies in Bukavu. You can find them in PANZI, ESSENCE, KADUTU, NYAWERA, NGUBA, PLACE, BRASSERIE and BAGIRA. It is up to you now to see which place is close by. It costs 200 CF per tablet that will treat 20 liters of water. You will remember that it takes money to get these tablets, but it will take a lot of money to pay for the care for severe diarrhea. Be vigilant! Share the message to keep your family healthy and happy. -Dr. Picha |
| **73** | **Week 34** | **IVR** | How who to construct a new drinking water vessel with tap | Hello again! This is Dr. Picha from the Provincial General Referral Hospital in Bukavu. Today we are here with Mwanza, the mother of a little child who came to the hospital for treatment of severe diarrhea. She has a question!<br>**Mwanza:** Dr. Picha! Our blue bucket was broken and I wanted to buy another bucket but I don't know how to add a tap. Can you help me?<br>For those who are listening! Can you help Mwanza? You can press 1 or 2 on your phone to answer the question. Your answer will be free of charge. If you think that the tap you need is not avaliable in Bukavu, press 1. If you think the tap you need is avaliable in the local market, press 2.<br>**If 1 was pressed:** Thanks for trying! Everyone in your family can put the tap on a new bucket. Please refer to the card we gave you previously with this information. Buy a plastic tap at your local market for about 6000 CF. You should install this tap at the bottom of your bucket. Almost at the end of the bucket, use a nail to trace the size of the tap on your bucket. You will then cut out this marked circle using a knife so the tap fits inside this hole. Take off the o-ring from the tap and insert the tap in the outside of your bucket, then use some tape on the part of the tap inside of the bucket like the photo we gave you to seal this off from leaking. Finally, screw on the o-ring we gave you on the part of the tap inside the bucket, and tighten the tap by twisting around as hard as you can. Please make a new bucket with tap for handwashing or drinking water to keep your family healthy. Share this message with your family!<br>**If 2 was pressed:** Thank you! Everyone in your family can put the tap on a new bucket. Please refer to the card we gave you previously with this information. Buy a plastic tap at your local market for about 6000 CF. You should install this tap at the bottom of your bucket. Almost at the end of the bucket, use a nail to trace the size of the tap on your bucket. You will then cut out this marked circle using a knife so the tap fits inside this hole. Take off the o-ring from the tap and insert the tap in the outside of your bucket, then use some tape on the part of the tap inside of the bucket like the photo we gave you to seal this off from leaking. Finally screw on the o-ring we gave you on the part of the tap inside the bucket, and tighten the tap by twisting around as hard as you can. Please make a new bucket with tap for handwashing or drinking water to keep your family healthy. Share this message with your family!<br>**If no button was pressed:** We haven.t received your answer! Everyone in your family can put the tap on a new bucket. Please refer to the card we gave you previously with this information. Buy a plastic tap at your local market for about 6000 CF. You should install this tap at the bottom of your bucket. Almost at the end of the bucket, use a nail to trace the size of the tap on your bucket. You will then cut out this marked circle using a knife so the tap fits inside this hole. Take off the o-ring from the tap and insert the tap in the outside of your bucket, then use some tape on the part of the tap inside of the bucket like the photo we gave you to seal this off from leaking. Finally screw on the o-ring we gave you on the part of the tap inside the bucket, and tighten the tap by twisting around as hard as you can. Please make a new bucket with tap for handwashing or drinking water to keep your family healthy. Share this message with your family! | Anyone in your family can put the tap on a new bucket. Please refer to the card we gave you before with this information. Buy a plastic tap at your local market for about 6000 CF. You install this tap at the bottom of your bucket. Near the end of the bucket, use a nail to trace the size of the tap on your bucket then you will cut out this marked circle using a knife so the tap fits inside this hole. Take off the o-ring from the tap and insert the tap outside of your bucket, then use some tape on the part of the tap inside of the bucket like the photo we gave you to seal this off from leaking. Screw the o-ring we gave you on the part of the tap inside the bucket, and tighten the tap by twisting hard. Please make a new bucket with a tap for hand washing or drinking water to keep your family safe. Share the message! -Dr. Picha |

| | | | | | |
|---|---|---|---|---|---|
| **74** | **Week 35** | **Voice** | Children drinking chlorinated water and washing their hands with soap grow better | Hello again! This is Dr. Picha from the Provincial General Referral Hospital in Bukavu. I have an important message for you today. We recently conducted a study in Walungu which found that children who drank chlorine treated water and washed their hands with soap or ash to reduce their contact with germs grew taller and healthier than children who were exposed to lots of germs from not washing their hands with soap or ash. Children are the ones who are most often affected by diarrheal diseases so keep them healthy and happy by making sure they always wash their hands with ash or soap before eating and after using the toilet. Make sure that you always have chlorine treated water to give them. Share the message to stay healthy and happy! | We recently conducted a study in Walungu which found that children who drank chlorine treated water and washed their hands with soap or ash to reduce their contact with germs grew taller and healthier than children exposed to lots of germs from not practicing handwashing or water treatment. Children are the ones who are most often affected by diarrheal diseases so keep them healthy and happy by making sure they always wash their hands with ash or soap before eating and after using the toilet and make sure you always have chlorine treated water to give them. Share the message to stay healthy and happy! -Dr. Picha |
| **75** | **Week 35** | **IVR** | Handwashing with soap after cleaning a child's anus | Hello! This is Dr. Picha from the Provincial General Referral Hospital in Bukavu. We have been talking to each other for a long time and we are very happy to know that your family is now used to using soap to wash their hands at key moments. We have a question for you - you can answer it by pressing 1 or 2 on your phone. There's no charge for answering!<br><br>If you clean your child's anus with soap and water, do you still need to wash your hands with soap afterward? If you think washing your child's anus with soap and water is sufficient and further handwashing with soap is not needed, press 1. If you think after washing your child's anus with soap and water you must still wash your hands with soap, press 2.<br><br>**If 1 was pressed:** Thank you very much for trying! It is important to wash your hands with soap after cleaning a child's anus even if you wash your child's anus with soap and water. This is because germs from stool can still remain on your hands unless you wash your hands with soap thoroughly. Please always wash your hands with soap after cleaning a child's anus. Keep your family healthy and happy. We will talk again! Share the message.<br><br>**If 2 was pressed:** Good answer! It is important to wash your hands with soap after cleaning a child's anus even if you wash your child's anus with soap and water. This is because germs from stool can still remain on your hands unless you wash your hands with soap thoroughly. Please always wash your hands with soap after cleaning a child's anus. Keep your family healthy and happy. We will talk again! Share the message.<br><br>**If no button was pressed:** By bad luck, we haven't received your answer! It is important to wash your hands with soap after cleaning a child's anus even if you wash your child's anus with soap and water. This is because germs from stool can still remain on your hands unless you wash your hands with soap thoroughly. Please always wash your hands with soap after cleaning a child's anus. Keep your family healthy and happy. We will talk again! Share the message. | It is important to wash your hands with soap after cleaning a child's anus even if you wash your child's anus with soap and water. This is because germs from stool can still remain on your hands unless you wash your hands with soap throughly. Please always wash your hands with soap after cleaning a child's anus. Keep your family health and happy. We will talk again! Share the message. -Dr. Picha |

| | | | | | |
|---|---|---|---|---|---|
| **76** | **Week 36** | **Voice** | Please do not keep your handwashing station and soap and treated drinking water locked away from children when you are not home | Hello again, this is Dr. Picha from the Provincial General Referral Hospital in Bukavu. Some families like yours keep their hand washing station and the bucket of treated water in a locked bedroom when they are away from home so their children do not play with it. This means your children can't wash their hands with ash or soap or drink this treated water using these materials when you are not home. We all know the importance of using these materials. Therefore, please keep the hand washing station and the treated water bucket in a place that is accessible to everyone for their use. I will call you again in the next few days. Share the message to stay healthy and happy. | Some families like yours keep their hand washing station and the bucket of treated water in a locked bedroom when they are away from home so their children do not play with it. This means your children can't wash their hands with ash or soap or drink this treated water using these materials when you are not home. We all know the importance of using these materials. Therefore, please keep the hand washing station and the treated water bucket in a place that is accessible to everyone for their use. Share the message to stay healthy and happy. -Dr. Picha |
| **77** | **Week 36** | **IVR** | Attach your bottle with soap or ash to the handwashing station by a string | Hello! This is Dr. Picha from the Provincial General Referral Hospital in Bukavu. I hope you and your family are well. I want to ask you a question! You can answer by pressing 1 or 2 on your phone. Answering this question will not cost you anything. I have a new little one myself and as we know children can get into everything! They can even move the bottle of ash or soapy water away from the handwashing station. What can we do about this? If you think there is nothing we can do about this, press 1. If you think you can tie a string to the plastic bottle we gave you with soapy water and ash and tie this to your handwashing station, press 2.<br><br>**If 1 was pressed:** Thanks for trying! Children will always be children and play with everything. To keep them from misplacing your bottle with soap or ash, please attach a string to the hole in the cap of these bottles and tie them to your handwashing station. This has been done by many households like yours and has worked to keep the ash and soapy water next to the handwashing station. Please make sure you and your household members always wash your hands with soap before eating, before feeding your child, after toileting, and after touching child feces to keep your family healthy and happy. Share the message!<br><br>**If 2 was pressed:** Thank you very much! Good answer! Children will always be children and play with everything. To keep them from misplacing your bottle with soap or ash, please attach a string to the hole in the cap of these bottles and tie them to your handwashing station. This has been done by many households like yours and has worked to keep the ash and soapy water next to the handwashing station. Please make sure you and your household members always wash your hands with soap before eating, before feeding your child, after toileting, and after touching child feces to keep your family healthy and happy. Share the message!<br><br>**If no button was pressed:** Until now, we have not received your response! The correct answer is: children will always be children and play with everything. To keep them from misplacing your bottle with soap or ash, please attach a string to the hole in the cap of these bottles and tie them to your handwashing station. This has been done by many households like yours and has worked to keep the ash and soapy water next to the handwashing station. Please make sure you and your household members always wash your hands with soap before eating, before feeding your child, after toileting, and after touching child feces to keep your family healthy and happy. Share the message! | Children will always be children and play with everything. To keep them from misplacing your bottle with soap or ash, please attach a string to the hole in the cap of these bottles and tie them to your handwashing station. This has been done by many households like yours and has worked to keep the ash and soapy water next to the handwashing station. Please make sure you and your household members always wash your hands with soap before eating and feeding your child and after toileting and touching child feces to keep your family healthy and happy. Share the message! -Dr. Picha |

| | | | | | |
|---|---|---|---|---|---|
| **78** | **Week 37** | **Voice** | Everyone in the household should help with water treatment | Hello again, this is Dr. Picha from the Provincial General Referral Hospital in Bukavu. If you are traveling and away from your home, you should leave the eldest child in your household responsible for treating your household drinking water using chlorine tablets. A good parent must protect their children and other family members by giving them safe water to drink even when they are not at home. Remember that when a child or other household member gets sick, the family needs to mobilize a lot of money for care. Keep your children healthy by always giving them chlorine treated water. Don't forget to share this message with your family members. | If you are traveling and away from your home, you should leave the eldest child in your household responsible for treating your household drinking water using chlorine tablets. A good parent must protect their children and other family members by giving them safe water to drink even when they are not at home. Share the message. -Dr. Picha |
| **79** | **Week 37** | **IVR** | Handwashing with ash | Hello again! This is Dr. Picha from the Provincial General Referral Hospital in Bukavu. I have the pleasure of speaking with you again today. Today we are here with Mwanza, the mother of a little child who came to the hospital for treatment of severe diarrhea. She has a question! You can answer by pressing 1 or 2 on your phone and your answer is free.<br>**Mwanza:** Hello! Today I am here to talk about ash. My husband lost his job, so our money is very tight and we have 5 kids. We don't have money for soap for handwashing. To ensure my children and I stay healthy we wash our hands with ash. My question for you today: does ash really remove germs? If you think ash can really remove germs, press 1. If you think that the ash was only used by people of the old generation and that it cannot remove germs, press 2.<br><br>**If 1 was pressed:** Congratulations! Like people of the past generations, people of the present generations can also use the ash to wash their hands. There are those who neglect it and consider it dirt, but it plays the same role as soap in eliminating microbes. It can be used when soap is not available and it is a good practice to teach children, because they can go to relatives who do not have soap to wash their hands and they won't worry about washing their hands because they will use ash. Protect your health and the health of your family! Share the message.<br><br>**If 2 was pressed:** Thank you so much for trying! Like people of the past generations, people of the present generations can also use the ash to wash their hands. There are those who neglect it and consider it dirt, but it plays the same role as soap in eliminating microbes. It can be used when soap is not available and it is a good practice to teach children, because they can go to relatives who do not have soap to wash their hands and they won't worry about washing their hands because they will use ash. Protect your health and the health of your family! Share the message.<br><br>**If no button was pressed:** By bad luck, we haven't received your answer yet! Like people of the past generations, people of the present generations can also use the ash to wash their hands. There are those who neglect it and consider it dirt, but it plays the same role as soap in eliminating microbes. It can be used when soap is not available and it is a good practice to teach children, because they can go to relatives who do not have soap to wash their hands and they won't worry about washing their hands because they will use ash. Protect your health and the health of your family! Share the message. | Like people of the past generations, people of the present generations can also use ash to wash their hands. There are those who neglect it and consider it as dirt, but it plays the same role as soap in eliminating microbes. It can be used when soap is not available and it is a good practice to teach children, because they can go to relatives who have soap to wash their hands and they won't worry about washing their hands because they will use ash. Protect your health and that of your family! Share the message. -Dr. Picha |
| **80** | **Week 38** | **Voice** | How to prepare soapy water using small leftover pieces of bar soap | Hello! This is Dr. Picha from the Provincial General Referral Hospital in Bukavu. We are here with Mwanza. For today, we wanted to talk again about making soapy water for hand washing. Mama Mwanza, we have been learning together for a long time and I wish you could tell others what you do to make soapy water when you don't have detergent.<br>**Mwanza:** Thank you very much Dr. Picha! To make soapy water when I don't have detergent, I use the leftover soap from when I wash clothes. I put it in an empty bottle, then I add water, shake it to mix the water and soap, and then I use it to wash my hands.<br>**Dr. Picha:** For those who are listening, do what Mama Mwanza does to keep your family healthy. | To make soapy water when you don't have detergent, take the soap left over from washing clothes, put it in an empty bottle, add water to the bottle, shake to mix the water and soap, and then start using it to wash your hands. Do this every time you need to make soapy water to keep your family healthy! -Dr. Picha |

| | | | | | |
|---|---|---|---|---|---|
| **81** | **Week 38** | **IVR** | How to prepare soapy water using small leftover pieces of bar soap | Hello! This is Dr. Picha from the Provincial General Referral Hospital in Bukavu. We wanted to talk you again about making soapy water to wash our hands. But I have a question! You can answer by pressing 1 or 2 on your phone and your answer will not be charged. What can you do to make soapy water when you don't have detergent? If you think there is no way to make soapy water without detergent, press 1. If you think that bar soap can also be used to make soapy water, press 2.<br><br>**If 1 was pressed:** Thank you very much for trying! To make soapy water when you don't have detergent, take the little bar soap left over from washing clothes, cut it up and put it in an empty bottle, add water to the bottle, shake to mix the soap with the water and then start using it to wash your hands. Do this to keep your family healthy.<br><br>**If 2 was pressed:** Thank you! Good answer! To make soapy water when you don't have detergent, take the little bar soap left over from washing clothes, cut it up and put it in an empty bottle, add water to the bottle, shake to mix the soap with the water and then start using it to wash your hands. Do this to keep your family healthy.<br><br>**If no button was pressed:** By bad luck we didn't get your answer! The good answer is: To make soapy water when you don't have detergent, take the little bar soap left over from washing clothes, cut it up and put it in an empty bottle, add water to the bottle, shake to mix the soap with the water and then start using it to wash your hands. Do this to keep your family healthy. | To make soapy water when you don't have detergent, take the little soap left over from washing clothes, cut it up and put it in an empty bottle, add water to the bottle, shake to mix the soap with the water and then start using it to wash your hands. Do this to keep your family healthy. -Dr. Picha |
| **82** | **Week 39** | **Voice** | Water treatment and safe water storage | Hello again! This is Dr. Picha from the Provincial General Referral Hospital in Bukavu. Many households in your area are reporting that it is difficult to find tap water to fill the blue bucket with drinking water. We remind you once again that any water whether it is rain water, deep well water, or even lake water can be used for drinking water if we add a chlorine tablet. If you don't have drinking water, you can take the water left in your thermos after boiling the water used for tea and use it as drinking water. You can pour it into the blue bucket to be used by your family for drinking water. Always store your drinking water in your blue bucket with the lid tightly secured on the top to keep your drinking water safe from contamination. Please make sure to stop children from putting their hands or objects inside your blue bucket. Treat and safely store your drinking water to protect your children and your family from severe diarrhea. Be careful! | Any water, whether it is rain water, deep well water, or even lake water can be used for drinking water if we add a chlorine tablet. And if you don't have drinking water, you can take the water left your thermos after boiling the water used for tea and use it as drinking water. You can pour it into the blue bucket to be used by your family for drinking water. Always store your drinking water in your blue bucket with the lid tightly on the top to keep your drinking water safe from contamination. Please make sure to stop children from putting their hands or objects inside your blue bucket. Treat and safely store your drinking water to protect your children and your family from severe diarrhea. Be careful! -Dr. Picha |

| | | | | | |
|---|---|---|---|---|---|
| 83 | **Week 39** | **IVR** | Water treatment | Hello again! This is Dr. Picha from the Provincial General Referral Hospital in Bukavu. Today we are here with Mwanza, the mother of a small child who came to the hospital because of severe diarrhea. She has a question!<br>**Mwanza:** Dr. Picha! Currently, my family has had difficulty finding money to buy water for drinking. I don't know what we can do to find drinking water?<br>What else can Mwanza do to find drinking water? You can help Mwanza by pressing 1 or 2 on your phone. Your answer will not be charged! If you think there is nothing you can do to find drinking water when you have no money, press 1. If you think that any water like rain water, well water, or lake water can be used as drinking water if it is treated, press 2.<br><br>**If 1 was pressed:** Thank you very much for trying! When it is difficult to find tap water, rain water, well water, and even lake water can be used for drinking - but only if you treat it. You can treat it by adding a chlorine tablet in your blue bucket and wait 30 minutes. You can also heat this water until large bubbles form to boil. If you don't have drinking water to fill your blue bucket, you can also use leftover boiled water from the thermos after making tea. Always keep your drinking water in your blue bucket with the lid on, and only dispense water using the tap. Never put hands or fingers inside your blue bucket. Keep your family safe from severe diarrhea by drinking treated water and safely storing your water! Bye!<br><br>**If 2 was pressed:** Good answer! When it is difficult to find tap water, rain water, well water, and even lake water can be used for drinking - but only if you treat it. You can treat it by adding a chlorine tablet in your blue bucket and wait 30 minutes. You can also heat this water until large bubbles form to boil. If you don't have drinking water to fill your blue bucket, you can also use leftover boiled water from the thermos after making tea. Always keep your drinking water in your blue bucket with the lid on, and only dispense water using the tap. Never put hands or fingers inside your blue bucket. Keep your family safe from severe diarrhea by drinking treated water and safely storing your water! Bye!<br><br>**If no button was pressed:** By bad luck we had not yet received your answer! When it is difficult to find tap water, rain water, well water, and even lake water can be used for drinking - but only if you treat it. You can treat it by adding a chlorine tablet in your blue bucket and wait 30 minutes. You can also heat this water until large bubbles form to boil. If you don't have drinking water to fill your blue bucket, you can also use leftover boiled water from the thermos after making tea. Always keep your drinking water in your blue bucket with the lid on, and only dispense water using the tap. Never put hands or fingers inside your blue bucket. Keep your family safe from severe diarrhea by drinking treated water and safely storing your water! Bye! | When it is difficult to find tap water, rain water, well water and even lake water can be used for drinking only if you had one chlorine tablet to your blue bucket and wait 30 minutes. You can also heat this water until large bubbles form to boil. Additionally, if you don't have drinking water to fill your blue bucket, you can take the water left in the thermos after boiling the tea water and use it as drinking water. Always keep your drinking water in your blue bucket with the lid on, and only dispense water using the tap. Never put hands or fingers inside your blue bucket. Keep your family safe from severe diarrhea by drinking treated water and safely storing your water! Bye! -Dr. Picha |

| | | | | | |
|---|---|---|---|---|---|
| **84** | **Week 40** | **Voice** | Importance of always keeping water in your handwashing station | Hello again! This is Dr. Picha from the Provincial General Referral Hospital in Bukavu. I hope you and your family are all doing well! Are you having trouble finding water to fill your handwashing station? Many households like yours are facing that same issue, but they have been able to overcome this challenge by using lake, rain, or well water to fill their handwashing stations. There has been a rise in diarrhea patients in your area, and handwashing with soap or ash will ensure your family stays healthy and happy. Please make sure you always have water in your hand washing station, and encourage children and all household members to always wash their hands with soap before eating and after using the toilet. Remember that the health of your family is in your hands! I will call you again soon! | Many households like you are faced that same issue, but they have been able to overcome this challenge by using lake, rain, or well water to fill their handwashing stations. There has been a rise in diarrhea patients in your area, and handwashing with soap or ash will ensure your family will stay healthy and happy. Please make sure you always have water in your hand washing station, and encourage children and all household members to always wash their hands with soap before eating and after using the toilet. Remember that the health of your family is in your hands! I will call you again soon! -Dr. Picha |

| | | | | | |
|---|---|---|---|---|---|
| **85** | **Week 40** | **IVR** | Rainwater and lake water can be used to fill your handwashing stations | Hello again! This is Dr. Picha from the Provincial General Referral Hospital in Bukavu. There have been many diarrhea patients in your area coming to your local health facility in the past week. I hope you and your family are all well and without severe diarrhea! I am here with Mwanza. She has made several trips to the health facility recently because her children have had severe diarrhea. Mwanza, tell us, what is happening?<br>**Mwanza:** I am having trouble finding water to fill my hand washing station. The tap in my area often does not have water. What should I do?<br>Can you help Mwanza? To answer, you can press 1 or 2 on your phone! There is no charge for answering the question. How can Mwanza find water to fill her handwashing station when there is no water coming from the taps in her area? If you think that rain water, well water, or lake water can be used to fill your handwashing station and wash your hands with soap or ash, press 1. If you think Mwanza has to wait until water comes to her local taps to fill her handwashing station, press 2.<br><br>**If 1 was pressed:** Thank you very much. Good answer! Many households told us they sometimes do not have water from their local taps to fill their handwashing station, but they overcome this challenge by using rain water, well water, or even lake water to fill their hand washing station. Rainwater, well water, or even lake water can be used to wash your hands with soap or ash to keep your family healthy. Make sure there is always water in your handwashing station. Make sure that you and your children always wash your hands with soap or ash before eating and preparing food and after toileting events. Stay healthy and happy!<br><br>**If 2 was pressed:** Thank you very much for trying! Many households told us they sometimes do not have water from their local taps to fill their handwashing station, but they overcome this challenge by using rain water, well water, or even lake water to fill their hand washing station. Rainwater, well water, or even lake water can be used to wash your hands with soap or ash to keep your family healthy. Make sure there is always water in your handwashing station. Make sure that you and your children always wash your hands with soap or ash before eating and preparing food and after toileting events. Stay healthy and happy!<br><br>**If no button was pressed:** Until now, we have not received your answer! Many households told us they sometimes do not have water from their local taps to fill their handwashing station, but they overcome this challenge by using rain water, well water, or even lake water to fill their hand washing station. Rainwater, well water, or even lake water can be used to wash your hands with soap or ash to keep your family healthy. Make sure there is always water in your handwashing station. Make sure that you and your children always wash your hands with soap or ash before eating and preparing food and after toileting events. Stay healthy and happy! | Many households told us they sometimes do not have water from their local taps to fill their handwashing station. owever, they have overcome this challenge by using rain water, well water or even lake water to fill their hand washing station. Rain water, well water or even lake water can be used to wash your hands with soap or ash to keep your family healthy. Make sure there is always water in your handwashing station. Make sure that you and your children always wash your hands with soap or ash before eating and preparing food and after toileting events. Stay healthy and happy! -Dr. Picha |

| | | | | | |
|---|---|---|---|---|---|
| **86** | **Week 41** | **Voice** | Please do let children put hands or objects in stored drinking water | Hello! This is Dr. Picha from the Provincial General Referral Hospital in Bukavu. Be careful because we currently have many diarrhea patients coming to the health facility in your neighborhood! Mwanza's youngest child recently got sick with severe diarrhea and had to go back to the health facility. Mwanza, can you tell us what happened?<br>**Mwanza:** My baby got sick with severe diarrhea and we had to go back to the hospital! Both my husband and I had to take time off from work to care for our sweet little boy. We lost money from taking time off from work.<br>**Dr. Picha:** I'm so sorry to hear this. What happened?<br>**Mwanza:** My children are always playing and they take the lid off of our blue drinking water vessel, and they contaminated all of our precious drinking water with germs by putting their hands and dipping their cups in the bucket.<br>**Dr. Picha:** Please tell your children and stop them from taking the lid off the blue bucket and putting their hands and objects inside, because this releases the chlorine that is inside your water and adds germs from fingers and cups to your precious drinking water. Always treat your water with chlorine tablets or by heating water until large bubbles come to a boil then safely store your water in your blue bucket with the lid. Children and all household members should always use the tap to dispense your water and never put fingers or cups or objects in your blue bucket as this will contaminate your water. Keep your children and family healthy and happy! Be careful! | Please tell your children and stop them from taking the lid off the blue bucket and putting their hands and objects inside because this releases the chlorine that is inside your water and add germs from fingers and cups to your precious drinking water. Always treat your water with chlorine tablets or by heating water until large bubbles form to boil then safely store your water in your blue bucket with the lid tightly secured. Children and all household members should always use the tap to dispense water and never put fingers or cups or objects in your blue bucket as this will contaminate the water. Keep your children and family healthy and happy! Be careful! -Dr. Picha |

| | | | | | |
|---|---|---|---|---|---|
| **87** | **Week 41** | **IVR** | Please do let children put hands or objects in stored drinking water | Hello again! This is Dr. Picha from the Provincial General Referral Hospital in Bukavu. Today we are here with Mwanza, the mother of a small child who came to the hospital because of severe diarrhea. She says she has a question for us!<br>**Mwanza:** Dr. Picha! My children keep coming to the health facility with severe diarrhea. Sometimes I see my children removing the lid on the drinking water bucket and putting their hands and cups inside. Could this be contaminating our precious drinking water?<br>Can you help Mwanza? You can answer by pressing 1 or 2 on your phone. There is no charge for your reply. If you think it is okay to remove the lid on the drinking water bucket and put hands and cups inside, press 1. If you think that removing the lid on the drinking water bucket and putting hands and cups inside can contaminate water with germs, press 2.<br><br>**If 1 was pressed:** Thank you very much for trying! Please tell your children and stop them from taking the lid off the blue bucket and putting their hands and objects inside. This releases the chlorine that is inside your water and adds germs from fingers and cups to your precious drinking water. Always treat your water with chlorine tablets or by heating water until large bubbles form to boil then safely store your water in your blue bucket with the lid. Children and all household members should always use the tap to dispense your water and never put fingers or cups or objects in your blue bucket as this will contaminate your water. Keep your children and family healthy and happy! Be careful!<br><br>**If 2 was pressed:** Congratulations! Good answer really! Please tell your children and stop them from taking the lid off the blue bucket and putting their hands and objects inside. This releases the chlorine that is inside your water and adds germs from fingers and cups to your precious drinking water. Always treat your water with chlorine tablets or by heating water until large bubbles form to boil then safely store your water in your blue bucket with the lid. Children and all household members should always use the tap to dispense your water and never put fingers or cups or objects in your blue bucket as this will contaminate your water. Keep your children and family healthy and happy! Be careful!<br><br>**If no button was pressed:** Until now we have not received your answer! Please tell your children and stop them from taking the lid off the blue bucket and putting their hands and objects inside. This releases the chlorine that is inside your water and adds germs from fingers and cups to your precious drinking water. Always treat your water with chlorine tablets or by heating water until large bubbles form to boil then safely store your water in your blue bucket with the lid. Children and all household members should always use the tap to dispense your water and never put fingers or cups or objects in your blue bucket as this will contaminate your water. Keep your children and family healthy and happy! Be careful! | Please tell your children and stop them from taking the lid off the blue bucket and putting their hands and objects inside, because this releases the chlorine that is inside your water and add germs from fingers and cups to your precious drinking water. Always treat your water with chlorine tablets or by heating water until large bubbles form to boil then safely store your water in your blue bucket with the lid. Children and all household members should always use the tap to dispense your water and never put fingers or cups or objects in your blue bucket as this will contaminate your water. Keep your children and family healthy and happy! Be careful! -Dr. Picha |
| **88** | **Week 42** | **Voice** | Diarrhea patients are increasing in your area this week | Hello again. This is Dr. Picha from the Provincial General Referral Hospital in Bukavu. Diarrhea patients are increasing in your area this week. Our intervention team is finding that households from our program that are returning to the health facility because they are not treating their water with chlorine tablets or heating water until large bubbles form to boil. Many of these households have water in their blue bucket but have not added a chlorine tablet. For the health of your family, after putting water in your blue drinking water bucket, make sure you add a chlorine tablet immediately in the bucket before proceeding with other activities. The most important thing in all this is to find a family member who will be in charge of treating the drinking water. This will prevent us from forgetting to treat our water and will protect us from severe diarrheal diseases. Be careful! Share the message! | Diarrhea patients are increasing in your area this week. For the health of your family, after you finish putting water in your blue drinking water bucket, make sure you add a chlorine tablet immediately in the bucket before proceeding with other activities. This will prevent us from forgetting to treat our water and will protect us from severe diarrhea diseases. Be careful! Share the message! -Dr. Picha |

| | | | | | |
|---|---|---|---|---|---|
| **89** | **Week 42** | **IVR** | Water treatment and safe water storage | Hello again. This is Dr. Picha from the Provincial General Referral Hospital in Bukavu. Diarrhea patients are still increasing in your area this week. Alarmingly we are finding that intervention households like yours are returning to the health facility because they have stopped adding chlorine tablets to their water. We know that it may happen that you or your family members may add water to your blue bucket and then forget to use a chlorine tablet. Let's talk about this.<br>I have a question to ask to you: You can answer by pressing 1 or 2 on your phone. Your answer is free. What can we do to remember to put a chlorine tablet in our blue bucket after adding water to it? If you think there is nothing that can be done about this forgetfulness to put a chlorine tablet in your drinking water, press 1. If you think you should add a chlorine tablet as soon as you put water in your bucket to help you remember to treat your water, press 2.<br><br>**If 1 was pressed:** Thank you for trying. You should add a chlorine tablet as soon as you put water in your blue bucket to help you remember to treat your water. Always keep your lid on tightly. It is important to find a family member who will be in charge of treating your drinking water using chlorine tablets. You can also heat your water until large bubbles form to boil, and store this water in your blue bucket with the lid on. Always treat your water, this will protect you and your children from severe diarrhea diseases. Be careful! Share the message.<br><br>**If 2 was pressed:** Congratulations you are right! You should add a chlorine tablet as soon as you put water in your blue bucket to help you remember to treat your water. Always keep your lid on tightly. It is important to find a family member who will be in charge of treating your drinking water using chlorine tablets. You can also heat your water until large bubbles form to boil, and store this water in your blue bucket with the lid on. Always treat your water, this will protect you and your children from severe diarrhea diseases. Be careful! Share the message.<br><br>**If no button was pressed:** Unfortunately, we didn't get your answer. You should add a chlorine tablet as soon as you put water in your blue bucket to help you remember to treat your water. Always keep your lid on tightly. It is important to find a family member who will be in charge of treating your drinking water using chlorine tablets. You can also heat your water until large bubbles form to boil, and store this water in your blue bucket with the lid on. Always treat your water, this will protect you and your children from severe diarrhea diseases. Be careful! Share the message. | You should add a chlorine tablet as soon as you put water in your blue bucket to help you remember to treat your water, always keep your lid tightly on. It is important to find a family member who will be in charge of treating your drinking water using chlorine tablets. You can also heat your water until large bubbles form to boil, and store this water in your blue bucket with the lid on. Always treat your water - this will protect you and your children from from severe diarrheal diseases. Be careful! Share the message. -Dr. Picha |
| **90** | **Week 43** | **Voice** | Handwashing with ash | Hello, this is Dr. Picha from the Provincial General Referral Hospital in Bukavu. We hope that you and your family are doing well. My team and I have noticed that severe diarrhea is still on the rise in your area and that many people in your area wash their hands without soap or ash. Ash is not dirt and using ash to wash your hands when you don't have soap can help to reduce germs that cause diarrhea. That is why we keep reminding you that like soap, ash also removes germs from your hands and it is important to use it when you wash your hands so that you don't end up in the hospital again because of severe diarrheal illness. Protect yourself and your family from severe diarrhea! Share the message. | Ash is not dirt - using ash to wash your hands when you don't have soap can help to reduce germs that cause diarrhea. That is why we keep reminding you that like soap, ash also removes germs from your hands and it is important to use it when you wash your hands so that you don't end up in the hospital again because of severe diarrhea illness. Protect yourself and your family from severe diarrhea! Share the message. -Dr. Picha |

| | | | | | |
|---|---|---|---|---|---|
| **91** | **Week 43** | **IVR** | Handwashing with ash | Hello again. This is Dr. Picha from the Provincial General Referral Hospital in Bukavu. I hope that you and your family are healthy and without severe diarrhea. I have one question to ask you: You can answer by pressing 1 or 2 on your phone. Your answer is free. What can we do if we do not have soap for handwashing? If you think we must use water only, press 1. If you think that we should use ash, press 2.<br><br>**If 1 was pressed:** Thanks for trying. You can use ash to wash your hands if you have no soap available in your home. Ash is free and removes germs from your hands. Always put a bottle of ash next to your handwashing station. Protect your family against cholera and severe diarrhea. Share this message.<br><br>**If 2 was pressed:** Congratulations you are correct. You can use ash to wash your hands if you have no soap available in your home. Ash is free and removes germs from your hands. Always put a bottle of ash next to your handwashing station. Protect your family against cholera and severe diarrhea. Share this message.<br><br>**If no button was pressed:** Unfortunately, we didn't receive your answer. You can use ash to wash your hands if you have no soap available in your home. Ash is free and removes germs from your hands. Always put a bottle of ash next to your handwashing station. Protect your family against cholera and severe diarrhea. Share this message. | You can use ash for washing your hands if you have no soap available in your home. Ash is free and removes germs on your hands. Always put a bottle of ash next to your handwashing station and tie this with a string so children will not move this. Protect your family against cholera and severe diarrhea. Share this message. -Dr. Picha |
| **92** | **Week 44** | **Voice** | Keep a bottle of ash tied to your handwashing station | Hello again. This is Dr. Picha on the phone from the Provincial General Referral Hospital in Bukavu. Today, we are here with Mwanza, the mother of a young child who came to the hospital with severe diarrhea. Mama Mwanza, please tell us what has happened.<br>**Mwanza:** Dr. Picha, every day I put ash in a bottle and place it next to my hand washing station in the morning before I go to work. But, after a short while, I notice that my children have gone to play with this bottle as a toy. I still don't know what to do!<br>**Dr. Picha:** Mama Mwanza, you are not the only one having this difficulty. To prevent children from removing the bottle containing ash near the hand washing station, you can tie this bottle with a wire to the handwashing station. This way the ash can be used for hand washing. Protect your family from severe diarrhea. Share the message! I will call you again later! | To prevent children from removing the bottle containing ash near the hand washing station, you can tie this bottle with a wire to the handwashing station. This way the ash can be used for hand washing. Protect your family from severe diarrhea. Share the message! I will call you again later! -Dr. Picha |

| | | | | | |
|---|---|---|---|---|---|
| **93** | **Week 44** | **IVR** | To prevent children from playing with your ash or soapy water bottle tie the bottle to handwashing station | Hello again. This is Dr. Picha from the Provincial General Referral Hospital in Bukavu. Today, I'm happy to speak with you and with Mama Mwanza. Mama Mwanza wants to share her experience with you.<br>**Mwanza:** One of my children keeps moving the ash bottle I always place next to the handwashing station to go play with it. This has been a problem for me.<br><br>How can you help Mwanza? You can answer by pressing 1 or 2 on your phone. Your answer is free. How can Mwanza stop her children from removing the ash bottle from next to her handwashing station? If you think that she should keep the ash bottle in the bedroom and only take it out for meal times, press 1. If you think that she should tie the ash bottle on a string attached to the handwashing station, press 2.<br><br>**If 1 was pressed:** Thank you for trying. To prevent children from removing the ash bottle next to the hand washing station, you can tie the bottle with a string to the handwashing station. This way the ash can be used for hand washing, and children can't remove it. Protect your family from severe diarrhea. I will call you again later! Share the message.<br><br>**If 2 was pressed:** Congratulations, your answer is correct. To prevent children from removing the ash bottle next to the hand washing station, you can tie the bottle with a string to the handwashing station. This way the ash can be used for hand washing, and children can't remove it. Protect your family from severe diarrhea. I will call you again later! Share the message.<br><br>**If no button was pressed:** Unfortunately, we didn't receive your answer. To prevent children from removing the ash bottle next to the hand washing station, you can tie the bottle with a string to the handwashing station. This way the ash can be used for hand washing, and children can't remove it. Protect your family from severe diarrhea. I will call you again later! Share the message. | To prevent children from removing the ash bottle next to the hand washing station you can tie this bottle with a string to the handwashing station. This way the ash can be used for hand washing, and children can't remove it. Protect your family from severe diarrhea. I will call you again later! Share the message. -Dr. Picha |
| **94** | **Week 45** | **Voice** | Use a clean cloth to dry your hands after handwashing with soap | Hello again. This is Dr. Picha from the Provincial General Referral Hospital in Bukavu. It is important that you always wash your hands with soap or ash before preparing food, feeding a child, or eating. After you wash your hands with ash or soap, it is important you dry your hands on a clean cloth. Do not simply wipe your hands on your clothing as this can lead to recontamination with germs. If you don't have a clean cloth, you can use a piece clothing that has just been washed that hasn't been worn yet like a t-shirt. You can put this clothing or piece of clothing on a nail next to the handwashing station. Always wash your hands with soap or ash and dry your hands on a clean cloth or piece of laundry to stay healthy. Be well. Share the message! | After handwashing with soap or ash, do not simply wipe your hands on your clothing as this can led to recontamination with germs. If you don't have a clean cloth, you can use a piece clothing that has just been washed that hasn't been worn yet like a t-shirt. You can put this clothing or piece of clothing on a nail next to the handwashing station. Be well. Share the message! -Dr. Picha |

| | | | | | |
|---|---|---|---|---|---|
| **95** | **Week 45** | **IVR** | Use a clean cloth to dry your hands after handwashing with soap | Hello again. This is Dr. Picha from the Provincial General Referral Hospital in Bukavu. Today I'm here again with Mwanza, she has a question.<br>**Mwanza:** Dr. Picha! After I wash my hands with soap or ash what is the best way to dry them?<br><br>Can you help Mwanza? You can press 1 or 2 on your phone to answer. Your answer is free. If you think the best way to dry your hands after washing them with soap or ash is to dry them on your clothing, please press 1. If you think the best way to dry your hands after washing them with soap or ash is to dry them on a clean cloth next to your handwashing station, press 2.<br><br>**If 1 was pressed:** Thank you for trying. It is very important to place a clean cloth next to your handwashing station to use for drying your hands after you wash your hands with soap or ash. Drying your hands on your clothing can put dirt and germs back on your hands. If you don't have a clean cloth, you can use a piece of clothing you just washed in the laundry, like a wrap or shirt. Follow my guidance to keep your family healthy and happy. Share the message!<br><br>**If 2 was pressed:** Congratulations, you're correct. Great job. It is very important to place a clean cloth next to your handwashing station to use for drying your hands after you wash your hands with soap or ash. Drying your hands on your clothing can put dirt and germs back on your hands. If you don't have a clean cloth, you can use a piece of clothing you just washed in the laundry, like a wrap or shirt. Follow my guidance to keep your family healthy and happy. Share the message!<br><br>**If no button was pressed:** Unfortunately, we didn't receive your answer. Thank you for trying. It is very important to place a clean cloth next to your handwashing station to use for drying your hands after you wash your hands with soap or ash. Drying your hands on your clothing can put dirt and germs back on your hands. If you don't have a clean cloth, you can use a piece of clothing you just washed in the laundry, like a wrap or shirt. Follow my guidance to keep your family healthy and happy. Share the message! | Thank you for trying. It is very important to place a clean cloth next to your handwashing station to use for drying your hands after you wash your hands with soap or ash. Drying your hands on your clothing can put dirt and germs back on your hands. If you don't have a clean cloth, you can use a piece of clothing you just washed in the laundry like a wrap or shirt. Follow my guidance to keep your family healthy and happy. Share the message! -Dr. Picha |
| **96** | **Week 46** | **Voice** | Encourage your children to prepare and refill the ash in the bottle for handwashing | Hello again! This is Dr. Picha from the Provincial General Referral Hospital in Bukavu. Some children have the habit of playing with soap or soapy water when it is near the hand washing station. This causes some parents to keep soap locked in the bedroom. If you also keep soap in the bedroom to prevent children from playing with it, then make ash for children to use for handwashing. Put the ash in a bottle, and tie the bottle to the handwashing station with a string. Ash does not cost money but it eliminates the germs on the hands. Encourage your children to prepare and refill the ash in the bottle. Remember, your family's health is in your hands. Share the message. | Have your young children use ash for handwashing before eating and after toileting when you are away at work. Ash does not cost money, but it can eliminate germs on hands. Encourage your children to prepare and refill the ash in the bottle. Remember, your family's health is in your hands. Share the message. -Dr. Picha |

| | | | | | |
|---|---|---|---|---|---|
| 97 | **Week 46** | **IVR** | To prevent children from playing with your ash or soapy water bottle tie the bottle to handwashing station | Hello again! This is Dr. Picha from the Provincial General Referral Hospital in Bukavu. I hope that you and your family are doing well. I have a question for you. You can answer by pressing 1 or 2 on your phone. Your answer is free.<br><br>What should you do if your children always play with soapy water or bar soap? If you think you should have children use ash for handwashing instead of soap or soapy water, press 1. If you think that you should lock the soapy water or bar soap in your bedroom and bring this out at meal times only, press 2.<br><br>**If 1 was pressed:** Congratulations, you are correct. If your children always play with soap or bar soap, you can have them use ash in a bottle tied to the handwashing station. Wash your hands with ash before preparing food, before feeding a child, before eating, and after toileting or cleaning a child's feces to stay free from diarrhea. Share my word to stay healthy and happy!<br><br>**If 2 was pressed:** Thank you for trying to answer. If your children always play with soap or bar soap, you can have them use ash in a bottle tied to the handwashing station. Wash your hands with ash before preparing food, before feeding a child, before eating, and after toileting or cleaning a child's feces to stay free from diarrhea. Share my word to stay healthy and happy!<br><br>**If no button was pressed:** Unfortunately, we didn't receive your answer. If your children always play with soap or bar soap, you can have them use ash in a bottle tied to the handwashing station. Wash your hands with ash before preparing food, before feeding a child, before eating, and after toileting or cleaning a child's feces to stay free from diarrhea. Share my word to stay healthy and happy! | If your children always play with soap or bar soap you can have them use ash instead in a bottle tied to the handwashing station. Wash your hands with ash before preparing food, feeding a child, before eating, and after toileting or cleaning a child's feces to stay free from diarrhea. Share my word to stay healthy and happy! -Dr. Picha |
| 98 | **Week 47** | **Voice** | Diarrhea is expensive if you have to take off from work or pay for treatment | Hello again. This is Dr. Picha from the Provincial General Referral Hospital in Bukavu. We hope that you and your family are in good health. Many mothers and fathers leave their households early each day in the morning and return at night due to work and going to the market. Some of these mothers and fathers are neglecting to fill their drinking water vessel and treat their water with chlorine, and do not fill their handwashing station before they go to work. These households are often the ones that are returning to their local health facilities because their children have severe diarrhea. When a member of the household gets sick, it can cost a lot of money to care for them and money is lost from time that must be taken off from work. For this reason, every morning before you leave your home make sure your handwashing station has water and ash or soap, and that you treat your household drinking water using a chlorine tablet. I will call you back later. Stay safe! Share the message. | When a member of the household gets sick, it can cost a lot of money to care for them and money is lost from the time that must be taken off from work. For this reason, every morning before you leave your home make sure your handwashing station has water and ash or soap, and that you treat your household drinking water using a chlorine tablet. I will call you back later. Share the message. Stay safe! -Dr. Picha |

| | | | | | |
|---|---|---|---|---|---|
| **99** | **Week 47** | **IVR** | Refilling your handwashing station and drinking storage container | Hello again! This is Dr. Picha from the Provincial General Referral Hospital in Bukavu. Today I'm here again with Mwanza, she has a question.<br>**Mwanza:** Hello! Both myself and my husband are outside of our household for most of the day at work. We leave early in the morning and return late in the evening. Our children are at home alone all day. How do we make sure there is always water in our handwashing station and chlorine treated water in our blue bucket?<br><br>Please help Mwanza. You can answer by pressing 1 or 2 on your phone. Your answer is free. How can we make sure there is always water in our handwashing station and chlorine treated water in our blue bucket if we are outside our home all day? If you think before you go to work in the morning you should always fill your handwashing station with water, and fill your blue bucket and add a chlorine tablet, press 1. If you think we are helpless here and can do nothing, press 2.<br><br>**If 1 was pressed:** Congratulations, you are correct! Before you go to work in the morning, you should always fill your handwashing station with water and fill your blue bucket and add a chlorine tablet. Remember to tie a bottle of ash or soapy water to your handwashing station. Follow my words to keep your family healthy and happy. Share my words!<br><br>**If 2 was pressed:** Thanks for trying. Before you go to work in the morning, you should always fill your handwashing station with water and fill your blue bucket and add a chlorine tablet. Remember to tie a bottle of ash or soapy water to your handwashing station. Follow my words to keep your family healthy and happy. Share my words!<br><br>**If no button was pressed:** Unfortunately, we didn't receive your answer. Before you go to work in the morning, you should always fill your handwashing station with water and fill your blue bucket and add a chlorine tablet. Remember to tie a bottle of ash or soapy water to your handwashing station. Follow my words to keep your family healthy and happy. Share my words! | Before you go to work in the morning, you should always fill your handwashing station with water and fill your blue bucket and add a chlorine tablet. Remember to tie a bottle of ash or soapy water to your handwashing station. Follow my words to keep your family healthy and happy. Share my words! -Dr. Picha |
| **100** | **Week 48** | **Voice** | Always use your handwashing station for handwashing with soap or ash | Hello again, this is Dr. Picha from the Provincial General Referral Hospital in Bukavu. We are here with Mwanza, another mother of a diarrhea patient.<br>**Mwanza:** Dr. Picha! I went to my neighbor's house yesterday and she had put the intervention materials under her bed in her bedroom because her children were playing with them. I told her about the household in our area that recently had a child die because of this type of behavior. That household stopped handwashing with soap and ash and treating their water with chlorine 6 months into the program, and their child got diarrhea and died.<br>**Dr. Picha:** I am appalled to hear that! It is very bad to keep the hand washing station and the bucket for storing drinking water in the bedroom under the bed. These interventions materials should be used to keep you, your family, and your children healthy from diarrhea. Always treat your drinking water with chlorine and wash your hands with soap or ash to protect your family members from severe diarrhea. Tie your bottle of ash or soapy water to your handwashing station to keep children from playing with it. If a family member gets sick, it is a serious problem because it will require a lot of money for care. Stay vigilant. Share the message! | There has been a diarrhea death in your area. Make sure you are using your handwashing station to wash your hands with soap or ash, and always treat your water using a chlorine tablet or heating it until large bubbles form for boiling. Always keep water in your buckets. Be safe! Share the message. -Dr. Picha |

| | | | | | |
|---|---|---|---|---|---|
| **101** | **Week 48** | **IVR** | There is an alarming increase in cholera cases in your area this week | Hello again. This is Dr. Picha from the Provincial General Referral Hospital in Bukavu. As I mentioned earlier this week, there is an alarming increase in cholera cases in your area this week. We hope that you and your family are doing well. A household nearby yours recently had a child that died from cholera. I have an important question for you today. You can answer by pressing 1 or 2 on your phone. Your answer is free.<br>If your family is healthy, do you still need to use your handwashing station and chlorine tablets and blue drinking water vessel? If you think that if your family is healthy, you do not need to use your handwashing station and chlorine tablets and blue drinking water vessel, press 1. If you think that your household must always use your handwashing station and chlorine tablets and blue drinking water vessel, even when no one is sick in your household, press 2.<br><br>**If 1 was pressed:** Thank you for trying. Your household must always use your handwashing station, chlorine tablets, and blue drinking water vessel even when no one is sick with diarrhea in your household. Cholera sets in quickly and you must be prepared. You can go from healthy to very sick with cholera in the hospital rapidly. Therefore, you must protect your family at all times by keeping water in your handwashing station with a bottle of ash or soapy water tied next to it, and by always keeping your blue bucket full of water and adding a chlorine tablet to treat your water. If a family member gets sick, it is a serious problem because it will require a lot of money for care and time loss from work. Stay vigilant. Remember cholera is in your area. Share my message!<br>**If 2 was pressed:** Congratulations, your answer is correct. Your household must always use your handwashing station, chlorine tablets, and blue drinking water vessel even when no one is sick with diarrhea in your household. Cholera sets in quickly and you must be prepared. You can go from healthy to very sick with cholera in the hospital rapidly. Therefore, you must protect your family at all times by keeping water in your handwashing station with a bottle of ash or soapy water tied next to it, and by always keeping your blue bucket full of water and adding a chlorine tablet to treat your water. If a family member gets sick, it is a serious problem because it will require a lot of money for care and time loss from work. Stay vigilant. Remember cholera is in your area. Share my message!<br>**If no button was pressed:** Unfortunately, we didn't receive your answer. Your household must always use your handwashing station, chlorine tablets, and blue drinking water vessel even when no one is sick with diarrhea in your household. Cholera sets in quickly and you must be prepared. You can go from healthy to very sick with cholera in the hospital rapidly. Therefore, you must protect your family at all times by keeping water in your handwashing station with a bottle of ash or soapy water tied next to it, and by always keeping your blue bucket full of water and adding a chlorine tablet to treat your water. If a family member gets sick, it is a serious problem because it will require a lot of money for care and time loss from work. Stay vigilant. Remember cholera is in your area. Share my message! | There is an alarming increase in cholera cases in your area this week. A household nearby to you recently had a child that died from cholera. Your household must always keep water in your handwashing station and tie a bottle of soap or ash to it, and you must have water in your blue drinking water vessel and add a chorine tablet. Cholera sets in quickly and you must be prepared. If a family member gets sick, it is a serious problem because it will require a lot of money for care and time loss from work. Share my message! -Dr. Picha |
| **102** | **Week 49** | **Voice** | Many households remove the small basin below the handwashing station because they say that children play with wastewater | Hello, this is Dr. Picha from the Provincial General Referral Hospital in Bukavu. I hope you and your family members are in good health. We know that some children play with water left in the small red basin under your handwashing station. This water can have germs so it is important when you finish washing your hands with soap or ash, you pour the water from your basin into the toilet and put the small basin on top of the handwashing station bucket to keep it away from your children playing with it. Return the small basin under the handwashing station before using it for handwashing with soap or ash. Always wash your hands with soap or ash before preparing food, before feeding a child, before eating, and after using the toilet or cleaning a child's feces. Help young children to do the same. By doing this, you will be protecting your children as well as other members of your household from severe diarrhea. The responsibility to protect yourself and your children from severe diarrhea involves everyone in the household. Stay healthy! Share the message! | We know that some children play with water left in the small red basin under your handwashing station. This water can have germs, so it is important when you finish washing your hands with soap or ash, you pour the water from your basin in the toilet and put the small basin on top of handwashing station bucket to keep it away from your children playing with it. Share the message. Stay healthy! -Dr. Picha |

| | | | | | |
|---|---|---|---|---|---|
| **103** | **Week 49** | **IVR** | Do not keep the handwashing station locked in your bedroom | Hello, this is Dr. Picha from the Provincial General Referral Hospital in Bukavu. We know that many children play with water left in the small basin after washing hands with soap or ash. I have a question to ask you. You can answer by pressing by 1 or by 2 on your phone. Your answer is free.<br><br>What should we do to keep our children from playing with the dirty water from handwashing in the red basin under the handwashing station? If you think we should move the handwashing station in the bedroom, press 1. If you think you can empty the red basin after handwashing each time, and put the basin on top of the handwashing station when it is not being used, press 2.<br><br>**If 1 was pressed:** Thank you for trying to answer. Do not keep the handwashing station locked in your bedroom. Please! We know that some children play with water left in the small red basin under your handwashing station so it is important when you finish washing your hands with soap or ash, you should pour the water from your basin in the toilet and put the small basin on top of handwashing station bucket to prevent your children from playing with it. Put the small basin under the handwashing station before using it for handwashing with soap and ash. Share my message to stay healthy and happy!<br><br>**If 2 was pressed:** Congratulations you are correct. Do not keep the handwashing station locked in your bedroom. Please! We know that some children play with water left in the small red basin under your handwashing station so it is important when you finish washing your hands with soap or ash, you should pour the water from your basin in the toilet and put the small basin on top of handwashing station bucket to prevent your children from playing with it. Put the small basin under the handwashing station before using it for handwashing with soap and ash. Share my message to stay healthy and happy!<br><br>**If no button was pressed:** Unfortunately, we didn't receive your answer. Do not keep the handwashing station locked in your bedroom. Please! We know that some children play with water left in the small red basin under your handwashing station so it is important when you finish washing your hands with soap or ash, you should pour the water from your basin in the toilet and put the small basin on top of handwashing station bucket to prevent your children from playing with it. Put the small basin under the handwashing station before using it for handwashing with soap and ash. Share my message to stay healthy and happy! | Do not keep the handwashing station locked in your bedroom. We know that some children play with water left in the small red basin under your handwashing station. Therefore, it is important that when you finish washing your hands with soap or ash, you pour the water from your basin in the toilet and put the small basin on top of handwashing station bucket to prevent your children from playing with it. Put the small basin under the handwashing station before using it for handwashing with soap or ash. Share my message to stay healthy and happy! - Dr. Picha |
| **104** | **Week 50** | **Voice** | Handwashing with soap or ash for 20 seconds | Hello again, this is Dr. Picha from the Provincial General Referral Hospital in Bukavu. You are now experts at handwashing with soap and ash! Congratulations. Just as we are experts, we must train our children to be experts also. One challenge that I have noticed among children is that they often wash their hands with soap or ash too quickly. Everyone must wash their hands with soap or ash for 20 seconds to remove germs and make sure their hands are clean. To remind them, sing two verses of the handwashing song with them. I have Mwanza here with me to tell you about this song [MWANZA SINGS THE HANDWASHING SONG]. | You are now experts at handwashing with soap and ash! Congratulations. Just as we are experts, we must train our children to be experts also. One challenge that I have noticed among children is that they often wash their hands with soap or ash too quickly. Everyone must wash their hands with soap or ash for 20 seconds to remove germs and make sure their hands are clean. To remind them, sing two verses of the handwashing song with them. Share the message! Be careful. -Dr. Picha |

| | | | | | |
|---|---|---|---|---|---|
| **105** | **Week 50** | **IVR** | Both children and adults need to wash their hands with ash or soap for at least 20 seconds to make them free from germs | Hello again, this is Dr. Picha from the Provincial General Referral Hospital in Bukavu. Today I'm here again with Mwanza, she has a question.<br>**Mwanza:** How long do we need to wash our hands with soap or ash?<br><br>You can answer by pressing 1 or 2. Your answer is free. Tell Mwanza how long she needs to wash her hands with soap or ash. If you think she needs to wash her hands with soap or ash for at least 20 seconds, press 1. If you think she needs to wash her hands with soap or ash for at least 10 seconds, press 2.<br><br>**If 1 was pressed:** Congratulations, your answer is correct. Both children and adults need to wash their hands with ash or soap for at least 20 seconds to make them free from germs. To encourage your young children to wash their hands with ash or soap for 20 seconds, remember the handwashing song we taught you and teach your children to sing at least two verses of this song to wash their hands with soap or ash for 20 seconds. Remember my words to keep your family healthy and happy. Share my words! Be well!<br><br>**If 2 was pressed:** Thank you for trying. Both children and adults need to wash their hands with ash or soap for at least 20 seconds to make them free from germs. To encourage your young children to wash their hands with ash or soap for 20 seconds, remember the handwashing song we taught you and teach your children to sing at least two verses of this song to wash their hands with soap or ash for 20 seconds. Remember my words to keep your family healthy and happy. Share my words! Be well!<br><br>**If no button was pressed:** Unfortunately, we didn’t receive your answer. Both children and adults need to wash their hands with ash or soap for at least 20 seconds to make them free from germs. To encourage your young children to wash their hands with ash or soap for 20 seconds, remember the handwashing song we taught you and teach your children to sing at least two verses of this song to wash their hands with soap or ash for 20 seconds. Remember my words to keep your family healthy and happy. Share my words! Be well! | Both children and adults need to wash their hands with ash or soap for at least 20 seconds to make them free from germs. To encourage your young children to wash their hands with ash or soap for 20 seconds, remember the handwashing song we taught you and teach your children to sing at least two verses of this song to wash their hands with soap or ash for 20 seconds. Remember my words to keep your family healthy and happy. Share my words! Be well! -Dr. Picha |

| | | | | | |
|---|---|---|---|---|---|
| **107** | **Week 51** | **IVR** | Water treatment with chlorine tablets | Hello again! This is Dr. Picha from the Provincial Gneral Referral Hospital of Bukavu. Some households in our program have told us that they cannot treat their water because they did not have money to buy 20 liters of water for drinking. I have a question to ask you about this. You can answer by pressing 1 or 2 on your phone. Your answer is free.<br>Which costs more the price of a jerry can of drinking water or the cost of care for diarrheal diseases?<br>If you think buying 20 liters of drinking water cost more than hospital fees for severe diarrhea, press 1. If you think that hospital fees for diarrhea cost more than buying 20 liters of drinking water press 2.<br><br>**If 1 was pressed:** Thank you very much for trying! Our health is priceless and we can't compare our health with the 100 CF cost of a jerrycan of water costs. Hospital fees for severe diarrhea can be very costly as you know! In addition you can lose income if you miss work due to being in the hospital or taking care of a sick family member. Don't take this risk! Pay the money to buy 20 liters of water so you can treat this water with a chlorine tablet. Put the health of your family first. If someone in your family got sick, it would cost more than a jerry can of water. Also remember that you can treat rain or river water with a chlorine tablet to make it safe to drink. Keep your family healthy and happy. Share the message!<br><br>**If 2 was pressed:** Good answer! Our health is priceless and we can't compare our health with the 100 CF cost of a jerrycan of water costs. Hospital fees for severe diarrhea can be very costly as you know! In addition you can lose income if you miss work due to being in the hospital or taking care of a sick family member. Don't take this risk! Pay the money to buy 20 liters of water so you can treat this water with a chlorine tablet. Put the health of your family first. If someone in your family got sick, it would cost more than a jerry can of water. Also remember that you can treat rain or river water with a chlorine tablet to make it safe to drink. Keep your family healthy and happy. Share the message!<br><br>**If no button was pressed:** We had not yet received your answer! Our health is priceless and we can't compare our health with the 100 CF cost of a jerrycan of water costs. Hospital fees for severe diarrhea can be very costly as you know! In addition you can lose income if you miss work due to being in the hospital or taking care of a sick family member. Don't take this risk! Pay the money to buy 20 liters of water so you can treat this water with a chlorine tablet. Put the health of your family first. If someone in your family got sick, it would cost more than a jerry can of water. Also remember that you can treat rain or river water with a chlorine tablet to make it safe to drink. Keep your family healthy and happy. Share the message! | Our health is priceless and we can't compare our health with the 100 CF a jerry can of water costs. Hospital fees for severe diarrhea can be very costly as you know! In addition you can lose income if you miss work due to being in the hospital or taking care of a sick family member. Don't take this risk! Pay the money to buy 20 liters of water so you can treat this water with a chlorine tablet. Put the health of your family first. If someone in your family got sick, it would cost more than a jerry can of water. Also remember that you can treat rain or river water with a chlorine tablet to make it safe to drink. Keep your family healthy and happy. Share the message! Share the message! -Dr. Picha |

| | | | | | |
|---|---|---|---|---|---|
| **109** | **Week 52** | **IVR** | Handwashing with soap and ash | Hello again! This is Dr. Picha from the Provincial General Referral Hospital in Bukavu. I hope you and your family are well. I have a question for you again today. You can answer by pressing 1 or 2 on your phone. Your answer will be free of charge. Do you still remember that ash can help with hand washing? If you think you could never use ash to wash your hands, press 1. If you think you can use ash to wash your hands when soap or soapy water is not available, press 2.<br><br>**If 1 was pressed:** Thank you very much for trying! Some people don't want to use ash to wash their hands, but like soap, ash helps to remove germs from the hands and can be used when soap is not available. Also, you don't have to use ash to wash your hands. But it is a practice we should teach children and other family members, because one day they may find themselves in a situation where they don't have soap and will use ash to wash their hands. I hope that you will not accept that another member of your family could get sick with severe diarrhea. Protect your family! I will call you again.<br><br>**If 2 was pressed:** Congratulations! Good answer! Some people don't want to use ash to wash their hands, but like soap, ash helps to remove germs from the hands and can be used when soap is not available. Also, you don't have to use ash to wash your hands. But it is a practice we should teach children and other family members, because one day they may find themselves in a situation where they don't have soap and will use ash to wash their hands. I hope that you will not accept that another member of your family could get sick with severe diarrhea. Protect your family! I will call you again.<br><br>**If no button was pressed:** Up to now we have not yet received your answer! Some people don't want to use ash to wash their hands, but like soap, ash helps to remove germs from the hands and can be used when soap is not available. Also, you don't have to use ash to wash your hands. But it is a practice we should teach children and other family members, because one day they may find themselves in a situation where they don't have soap and will use ash to wash their hands. I hope that you will not accept that another member of your family could get sick with severe diarrhea. Protect your family! I will call you again. | Some people don't want to use ash to wash their hands, but like soap, ash helps to remove germs from the hands and can be used when soap is not available. Also, you don't have to use ash to wash your hands. But it is a practice we should teach children and other family members, because one day they may find themselves in a situation where they don't have soap and will use ash to wash their hands. I hope that you will not accept that another member of your family could get sick with severe diarrhea. Protect your family! -Dr. Picha |
| **110** | **Study End Message** | **Voice** | Congratulations for completing 12 months of the program | Hello! This is Dr. Picha from Bukavu Provincial Hospital. Congratulations on completing 12 months with our Program. You have become an expert in water treatment and hand washing with soap. So you should be a teacher for other families on how to treat their drinking water with chlorine tablets, as well as how to wash their hands with soap at key moments. Keep your family healthy and free from severe diarrhea by washing your hands with soap after every key event, especially before eating and after using the toilet or after cleaning the child's anus, and by drinking only water treated with chlorine tablets. Stay safe! | Congratulations! You have become an expert in water treatment and hand washing with soap. So you should show other families about how to treat their drinking water with chlorine tablets, as well as how to wash their hands with soap at key moments. Keep your family healthy and free from severe diarrhea by washing your hands with soap and by drinking treated water. -Dr. Picha |
