## Supplemental File 2 for "Process Evaluation for the Delivery of a Water, Sanitation and Hygiene Mobile Health Program: Randomized Controlled Trial of the PICHA7 Mobile Health Program"

**Supplemental File 2.** Summary of PICHA7 pilot mHealth program interactive voice response quiz questions

| IVR Quiz question | Correct answer | N | Received | | Replied | | Correct | | Invalid | |
|---|---|---|---|---|---|---|---|---|---|---|
| | | | % | n | % | n | % | n | % | n |
| Will using materials such as soapy water bottle, hand washing station, drinking water storage bucket, chlorine tablets and stools we provided help to prevent cholera and severe diarrhea? | Yes | 139 | 99% | 138 | 84% | 117 | 91% | 107 | 6% | 7 |
| How long after adding a chlorine tablet to water should you wait to drink? | 30 minutes | 163 | 95% | 155 | 53% | 86 | 81% | 70 | 9% | 8 |
| How many key times are there for washing hands with soap? | 5 key moments | 162 | 96% | 155 | 57% | 93 | 75% | 70 | 15% | 14 |
| What is the correct way to boil your drinking | Boil water until large l | 162 | 96% | 156 | 50% | 81 | 72% | 58 | 10% | 8 |
| Do you need to treat drinking water collected | Yes | 162 | 96% | 155 | 53% | 86 | 59% | 51 | 15% | 13 |
| Can Mwanza buy chlorine tablets for her drinking water in the market areas in Bukavu? | Yes | 162 | 94% | 153 | 44% | 71 | 70% | 50 | 11% | 8 |
| Do we wash our hands only when they look | No | 162 | 90% | 146 | 49% | 79 | 70% | 55 | 18% | 14 |
| How do you make soapy water for hand | Put 7 caps full of deter | 162 | 92% | 149 | 53% | 86 | 73% | 63 | 14% | 12 |
| Can ash be used if you do not have money to buy detergent powder or soap? | Yes | 162 | 94% | 152 | 54% | 88 | 56% | 49 | 16% | 14 |
| Do people who are outside the home during the day need to wash their hands with soap? | Yes | 162 | 90% | 146 | 43% | 69 | 29% | 20 | 17% | 12 |
| What should you do if one of your children has diarrhea while they are in the bed with your other children? | When a child has diarrhea, he or she should not sleep in the same bed with siblings. | 162 | 88% | 142 | 49% | 80 | 61% | 49 | 16% | 13 |
| Can Mwanza buy materials in the market to fix her handwashing station? | Yes | 161 | 88% | 142 | 42% | 67 | 60% | 40 | 16% | 11 |

| | | | | | | | | | | |
|---|---|---|---|---|---|---|---|---|---|---|
| How do we avoid severe diarrhea like cholera during the 7 day high risk period after someone in your household has diarrhea? | Wash hands with soap and treat drinking water by boiling or using chlorine tablets. | 161 | 91% | 146 | 48% | 77 | 73% | 56 | 16% | 12 |
| Can you remind me how to correctly boil water? | Boil water until large l | 159 | 87% | 138 | 48% | 76 | 64% | 49 | 22% | 17 |
| Can you remind me how often I need to wash the blue bucket for my drinking water? | Every time you get water for drinking. | 159 | 85% | 135 | 45% | 71 | 61% | 43 | 21% | 15 |
| How much water do you need in your blue drinking water vessel before you add an chlorine tablet? | At least 20 Liters | 158 | 91% | 143 | 42% | 67 | 63% | 42 | 24% | 16 |
| Is there another way to wash our hands when you do not have soap or detergent? | Yes, ash can be used. | 158 | 90% | 142 | 41% | 65 | 18% | 12 | 18% | 12 |
| Can you help Mama Mwanza by explaining to her the importance of these photos that my team gave her? | The photos remind you and your family to treat drinking water with chlorine tablets, the key moments of hand washing with soap, and how to put a tap on the blue bucket and the red bucket. | 157 | 87% | 136 | 35% | 55 | 56% | 31 | 31% | 17 |
| Not treating drinking water water with chlorine tablets or not boiling water can put you and your children at risk of returning to the health facility with diarrhea. True or false? | True | 155 | 85% | 131 | 40% | 62 | 55% | 34 | 37% | 23 |
| How do I use my handwashing station stool? | Only use stool to hold the handwashing station and water vessel, it is not meant for sitting. | 154 | 88% | 136 | 38% | 59 | 71% | 42 | 20% | 12 |

| | | | | | | | | | | |
|---|---|---|---|---|---|---|---|---|---|---|
| Did you keep using your handwashing station with basin? | Always keep your basin under or on top of your handwashing station, and always keep water inside for handwashing and soap or ash. Keep your children from coming back to the hospital with diarrhea. Always wash your hands with soap or ash. | 151 | 89% | 135 | 38% | 58 | 59% | 34 | 26% | 15 |
| We no longer buy chlorine tablets to treat our drinking water. What should we do? | Boil water | 150 | 89% | 133 | 37% | 56 | 55% | 31 | 27% | 15 |
| Do you currently have water in your red bucket? | Yes | 149 | 85% | 126 | 32% | 48 | 60% | 29 | 23% | 11 |
| What can we do to avoid going back to the health facility because our children contract severe diarrhea ? | Protect yourself and your children by making sure everyone in your households is washing their hands with soap or ash before eating and after toileting, as well as treating your water using chlorine tablets or boiling. | 149 | 89% | 133 | 34% | 50 | 56% | 28 | 20% | 10 |
| Do you think it is still important for your children to wash their hands with soap or ash before eating a donut to prevent severe diarrhea? | Yes | 147 | 90% | 132 | 41% | 61 | 62% | 38 | 31% | 19 |

| | | | | | | | | | | |
|---|---|---|---|---|---|---|---|---|---|---|
| After leaving the hospital, we can get sick with severe diarrhea again if we don't wash our hands with soap and if we don't treat our drinking water with chlorine tablets or boil it. True or false. | True | 147 | 89% | 131 | 43% | 63 | 60% | 38 | 19% | 12 |
| How should you boil your water? | Boil water until large l | 143 | 84% | 120 | 37% | 53 | 58% | 31 | 26% | 14 |
| Is handwashing with soap or ash before eating and after toileting as important for fathers and brother as for our mothers and sisters? | Yes | 148 | 86% | 128 | 45% | 67 | 54% | 36 | 25% | 17 |
| Who in your household should wash their hands with soap or ash before preparing food? | Everyone | 147 | 88% | 130 | 39% | 58 | 64% | 37 | 16% | 9 |
| Does the stool of young children contain microbes? | Yes | 146 | 92% | 134 | 40% | 59 | 54% | 32 | 36% | 21 |
| Is it okay to wash my hands with water only after cleaning my child's anus? | No | 146 | 89% | 130 | 44% | 64 | 58% | 37 | 27% | 17 |
| Are your children washing their hands with soap after coming from the toilet? | Yes | 143 | 90% | 129 | 43% | 61 | 51% | 31 | 31% | 19 |
| Do I need to wash my hands with ash or soap if I only urinate? | Yes | 141 | 89% | 126 | 45% | 63 | 56% | 35 | 19% | 12 |
| Do you think Mwanza should share the chlorine tablets with other family members or neighbors? | No | 140 | 91% | 127 | 30% | 42 | 45% | 19 | 38% | 16 |
| What can we do to have treated water for drinking and to wash our hands with ash and soap when water is so hard to find? | Try to put water in both of our buckets make sure we are handwashing with soap and treating water. | 143 | 90% | 129 | 47% | 67 | 64% | 43 | 25% | 17 |
| How often should you wash your handwashing station and drinking water vessel we provided with soap and water? | Weekly | 142 | 91% | 129 | 39% | 56 | 39% | 22 | 29% | 16 |

| | | | | | | | | | | |
|---|---|---|---|---|---|---|---|---|---|---|
| Our blue bucket was broken and I wanted to buy another bucket but I don't know how to add a tap. Can you help me? | Please refer to the card we gave you previously with this information. Buy a plastic tap at your local market. | 141 | 94% | 133 | 44% | 62 | 61% | 38 | 29% | 18 |
| If you clean your child's anus with soap and water, do you still need to wash your hands with soap after? | Yes | 139 | 94% | 130 | 41% | 57 | 65% | 37 | 21% | 12 |
| Children can move the bottle of ash or soapy water away from the handwashing station. What can we do about this? | Attach a string to the hole in the cap of these bottles and tie them to the handwashing station. | 139 | 94% | 131 | 43% | 60 | 63% | 38 | 27% | 16 |
| Does ash remove germs? | Yes | 139 | 93% | 129 | 45% | 62 | 66% | 41 | 26% | 16 |
| What can you do to make soapy water when you don't have detergent? | Take the little bar soap left over from washing clothes, cut it up and put it in an empty bottle, add water to the bottle, shake to mix the soap with the water. | 141 | 96% | 136 | 65% | 92 | 63% | 58 | 22% | 20 |
| Currently, my family has had difficulty finding money to buy water for drinking. I don't know what we can do to find drinking water. | When it is difficult to find tap water, rain water, well water and even lake water can be used for drinking only if you had one chlorine tablet to your blue bucket and wait 30 minutes. | 134 | 90% | 121 | 43% | 57 | 58% | 33 | 32% | 18 |

| | | | | | | | | | | |
|---|---|---|---|---|---|---|---|---|---|---|
| How can Mwanza find water to fill her handwashing station when there is no water coming from the taps in her area? | When it is difficult to find tap water, rain water, well water and even lake water can be used for drinking only if you had one chlorine tablet to your blue bucket and wait 30 minutes. | 132 | 89% | 118 | 43% | 57 | 42% | 24 | 30% | 17 |
| Sometimes I see my children removing the lid on the drinking water bucket and putting their hands and cups inside. Could this be contaminating our precious drinking water? | Yes | 132 | 89% | 117 | 44% | 58 | 59% | 34 | 26% | 15 |
| What can we do to not forget to put a chlorine tablet in our blue bucket after adding water to it? | Add a chlorine tablet as soon as you put water in your blue bucket to help you remember to treatment your water, always keep your lid tightly on. | 134 | 92% | 123 | 40% | 53 | 53% | 28 | 32% | 17 |
| What can we do if we do not have soap for handwashing ? | Use ash | 134 | 93% | 125 | 45% | 60 | 65% | 39 | 22% | 13 |
| How can Adolphine stop her children from removing the ash bottle from next to her handwashing station? | Attach a string to the hole in the cap of these bottles and tie them to the handwashing station. | 132 | 92% | 121 | 42% | 55 | 65% | 36 | 20% | 11 |
| After I wash my hands with soap or ash what is the best way to dry them? | Place a clean cloth next to you are handwashing station. | 131 | 89% | 117 | 45% | 59 | 64% | 38 | 24% | 14 |

| | | | | | | | | | | |
|---|---|---|---|---|---|---|---|---|---|---|
| What should you do if your children always play with soapy water or bar soap? | Attach a string to the hole in the cap of these bottles and tie them to the handwashing station. | 129 | 91% | 117 | 42% | 54 | 54% | 29 | 28% | 15 |
| How do we make sure there is always water in our handwashing station and chlorine treated water in our blue bucket? | Before you go to work in the morning, you should always fill your handwashing station with water and fill your blue bucket and add a chlorine tablet. | 128 | 92% | 118 | 44% | 56 | 38% | 21 | 39% | 22 |
| If your family is healthy do you still need to use your handwashing station and chlorine tablets and blue drinking water vessel? | Yes | 128 | 91% | 117 | 40% | 51 | 71% | 36 | 20% | 10 |
| What should we do to keep our children from playing with the dirty water from handwashing in the red basin under the handwashing station? | When you finish washing your hands with soap or ash, pour the water from your basin in the toilet and put the small basin on top of handwashing station bucket to keep it away from your children playing with it. | 128 | 87% | 111 | 34% | 43 | 70% | 30 | 28% | 12 |
| How long do we need to wash our hands with soap or ash | At least 20 second | 127 | 82% | 104 | 39% | 50 | 34% | 17 | 34% | 17 |
| Can we continue to use hand washing station with taps which contain rust? | Yes | 127 | 91% | 116 | 40% | 51 | 41% | 21 | 25% | 13 |
| Do you still remember that ash can help with hand washing? | Yes | 123 | 89% | 110 | 33% | 40 | 70% | 28 | 18% | 7 |

| | | | | | | | | | | |
|---|---|---|---|---|---|---|---|---|---|---|
| Which costs more: the price of a jerry can of drinking water or the cost of care for diarrheal diseases? | Care for diarrheal diseases | 123 | 89% | 109 | 33% | 40 | 50% | 20 | 23% | 9 |
| If you don't have an empty bottle of liquid soap to keep your soapy water that you made for hand washing, what else can you do to keep your soapy water safely? | You can use any plastic bottle to make soapy water and add a hole on the top to make it easier to squeeze out the soapy water. | 124 | 90% | 112 | 31% | 38 | 37% | 14 | 16% | 6 |
| What can you do to find more Chlorine tablets if the ones we gave you run out? | You can buy Chlorine tablets in local pharmacies or in town and they are not expensive. If you don't have chlorine tablets, you can boil water. | 125 | 87% | 109 | 34% | 42 | 67% | 28 | 26% | 11 |
| What can you do to make sure you do not forget to treat your family's drinking water once you put the water in the bucket? | Add the Chlorine tablet right after you put water in your blue bucket. | 125 | 87% | 109 | 39% | 49 | 59% | 29 | 33% | 16 |
| My children are always playing with tap on the handwashing station. What should I do? | Turn your bucket over on your stool so the tap faces towards the wall so young children can't play with it. | 126 | 89% | 112 | 37% | 46 | 65% | 30 | 26% | 12 |

| | | | | | | | | | | |
|---|---|---|---|---|---|---|---|---|---|---|
| What can you do to get soapy water to use for hand washing at key times when you don't have detergent? | You can make soapy water by cutting up small leftover pieces of soap and adding this to a plastic bottle with water. When there is no soap or soapy water, ash can be used. | 126 | 90% | 114 | 41% | 52 | 21% | 11 | 29% | 15 |
| Do you think that when your family leaves your current address to go to another place, you can no longer have diarrhea? | No | 127 | 91% | 115 | 45% | 57 | 68% | 39 | 14% | 8 |
| What should you do if you do not have enough water to fill the 20 liter bucket to add a chlorine tablet? | Even if you don't have 20 liters of water to use the chlorine tablets we gave you, you can boil water. | 129 | 89% | 115 | 32% | 41 | 27% | 11 | 32% | 13 |
| What will Mwanza do to treat drinking water for her family? | You can buy another bucket or use a clean 20 liter jerry can with a lid. | 131 | 91% | 119 | 30% | 39 | 62% | 24 | 18% | 7 |
| Should you give your neighbor or other family members a bucket or chlorine tablets if they ask for them? | No | 130 | 91% | 118 | 33% | 43 | 65% | 28 | 14% | 6 |

N and n values based on number of Households.

Invalid indicates either an invalid button was pressed (not 1 or 2) or no response was given
